# Dissecting the contribution of common variants to risk of rare neurodevelopmental conditions

**DOI:** 10.1101/2024.03.05.24303772

**Authors:** Qin Qin Huang, Emilie M Wigdor, Patrick Campbell, Daniel S Malawsky, Kaitlin E Samocha, V Kartik Chundru, Petr Danecek, Sarah Lindsay, Thomas Marchant, Mahmoud Koko Musa, Sana Amanat, Davide Bonifanti, Eamonn Sheridan, Elizabeth J Radford, Jeffrey C Barrett, Caroline F Wright, Helen V Firth, Varun Warrier, Alexander Strudwick Young, Matthew E Hurles, Hilary C Martin

## Abstract

Although rare neurodevelopmental conditions have a large Mendelian component, common genetic variants also contribute to risk. However, little is known about how this polygenic risk is distributed among patients with these conditions and their parents, its interplay with rare variants, and whether parents’ polygenic background contributes to their children’s risk beyond the direct effect of variants transmitted to the child (i.e. via indirect genetic effects potentially mediated through the prenatal environment or ‘genetic nurture’). Here, we addressed these questions using genetic data from 11,573 patients with rare neurodevelopmental conditions, 9,128 of their parents and 26,869 controls. Common variants explained ∼10% of variance in overall risk. Patients with a monogenic diagnosis had significantly less polygenic risk than those without, supporting a liability threshold model, while both genetically undiagnosed patients and diagnosed patients with affected parents had significantly more risk than controls. In a trio-based model, using a polygenic score for neurodevelopmental conditions, the transmitted but not the non-transmitted parental alleles were associated with risk, indicating a direct genetic effect. In contrast, we observed no direct genetic effect of polygenic scores for educational attainment and cognitive performance, but saw a significant correlation between the child’s risk and non-transmitted alleles in the parents, potentially due to indirect genetic effects and/or parental assortment for these traits. Indeed, as expected under parental assortment, we show that common variant predisposition for neurodevelopmental conditions is correlated with the rare variant component of risk. Our findings thus suggest that future studies should investigate the possible role and nature of indirect genetic effects on rare neurodevelopmental conditions, and consider the contribution of common and rare variants simultaneously when studying cognition-related phenotypes.

## Main

Rare conditions affect 3.5%-6% of the global population^1^ and of these, the majority involve the central nervous system^2^. While genomic sequencing has revolutionized the diagnosis of rare neurodevelopmental conditions, which typically include intellectual disability and/or developmental delay, a monogenic diagnosis is only identified for about 30-40% of patients^3–5^. Common variants also contribute to risk for rare neurodevelopmental conditions^6,7^. In particular, this common variant contribution overlaps with polygenic risk for schizophrenia and for predisposition to reduced educational attainment and cognitive performance^6^. Accordingly, rare damaging variants in constrained genes, which play a major role in risk of rare neurodevelopmental conditions, are also associated with reduced educational attainment and cognitive performance and increased risk of mental health conditions in UK Biobank ^8–12^. In this work, we seek to better understand the nature of common variant risk for rare neurodevelopmental conditions, its interplay with rare variants and its distribution amongst different patients and their parents.

We begin by leveraging new, larger genome-wide association studies (GWASs) than were previously available^6^ to explore the extent to which common variant effects on rare neurodevelopmental conditions are correlated with their effects on a broad range of mental health conditions. This is motivated by findings that some psychiatric conditions have a partial neurodevelopmental origin ^13–15^, and that people with rare neurodevelopmental conditions^16^, as well as their relatives ^17–19^, are more likely to have psychiatric conditions. Furthermore, some of this overlap appears to be driven by certain rare copy number variants with variable expressivity^20–22^, suggesting some shared etiology between psychiatric and rare neurodevelopmental conditions. Here we explore whether shared common variant effects may also contribute, and whether this is independent of the genetic overlap between these conditions and cognitive traits.

Little is known about the interplay between rare and common variants in the context of rare neurodevelopmental conditions, and dissecting this will be key to fully understanding their genetic architecture and improving genetic diagnosis and risk prediction. Here we address two hypotheses in this space, testing the liability threshold model and whether common variants modify the penetrance of rare variants. The liability threshold model predicts that an individual will develop a condition once the sum of independent genetic and environmental risk factors exceeds some threshold^23–26^. Under this model, one might expect that patients with neurodevelopmental conditions who have a highly penetrant damaging variant would require, on average, less polygenic load to cross a diagnostic threshold than those without such variants (<u>**Extended Data Figure 1**</u>). We previously saw no significant difference in polygenic scores between patients with *versus* without a monogenic diagnosis^6^, but in this work, we anticipated that increased sample size and improved diagnostic rate^5,27^ might improve power. Since rare variants associated with neurodevelopmental conditions appear to act additively with polygenic scores in affecting cognitive ability in UK Biobank^9,10^, we hypothesized that polygenic background would modify the penetrance of these inherited rare variants in families with neurodevelopmental conditions, as it does, for example, in the context of *BRCA1/2* variants predisposing to breast cancer^28^.

**Figure 1.**
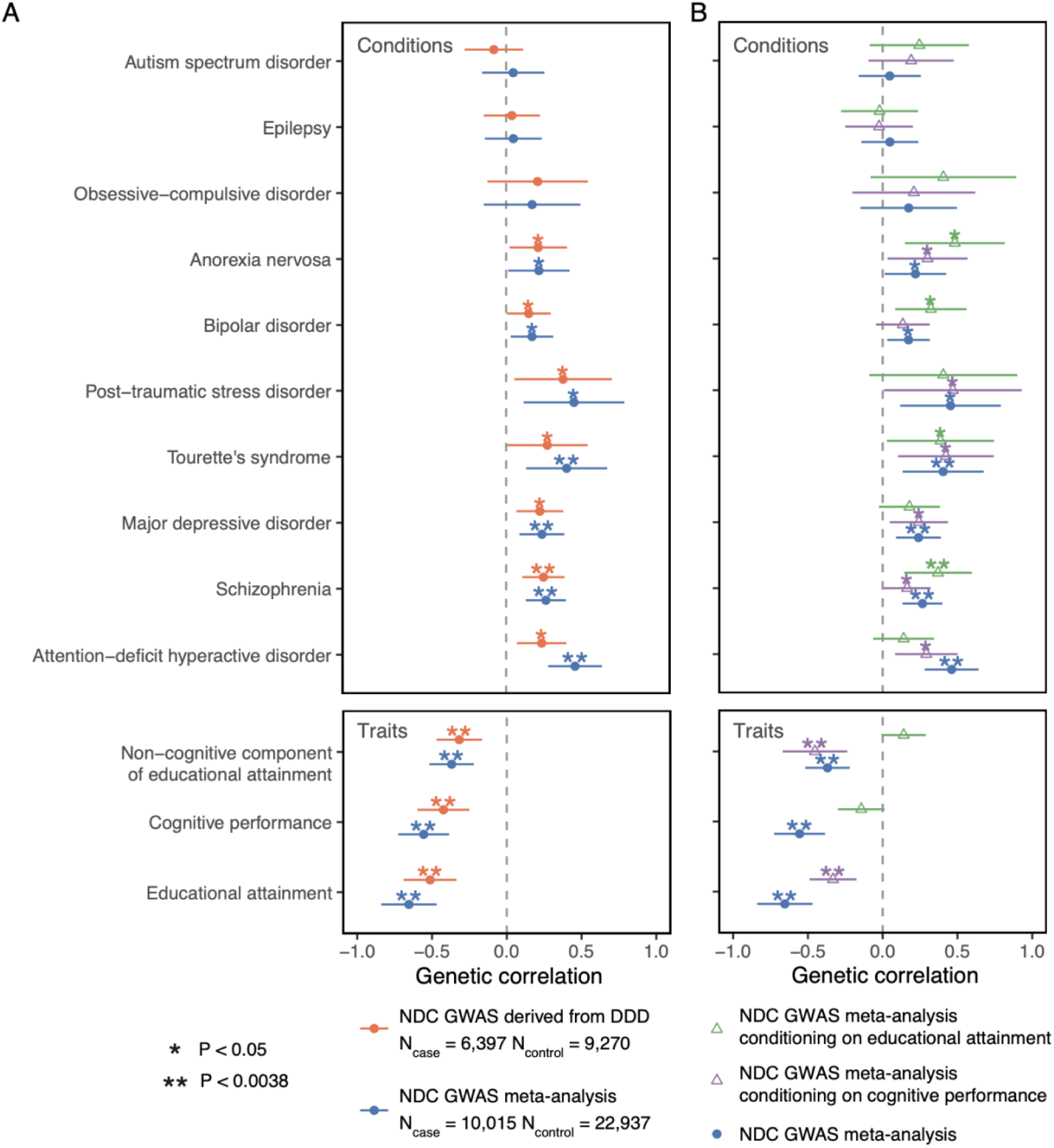
Genetic correlations between neurodevelopmental conditions (NDCs) and other brain-related traits and conditions. **A)** shows the estimates from Linkage Disequilibrium Score Regression for the DDD GWAS (orange) and the meta-analysis of neurodevelopmental conditions between DDD and GEL (blue). **B)** shows the estimates for the meta-analysis after conditioning on the GWAS summary statistics for educational attainment (green) or cognitive performance (purple) using GenomicSEM. Error bars show 95% confidence intervals.

Finally, we explore the extent to which common variants predisposing to rare neurodevelopmental conditions act directly on the affected individuals carrying them (“direct genetic effects”). Many studies have shown that genetic associations between common genetic variants and educational and cognitive phenotypes shrink when estimated within families ^29–33^. One possible explanation for this is that variants associated with these traits have indirect genetic effects, i.e. they have some effect on the parents, and this then affects the offspring through the family environment ^29,33–35^. These indirect genetic effects are under-explored in the context of rare diseases, but we hypothesized that they may play a role in rare neurodevelopmental conditions given the genetic overlap with educational attainment.

We address these questions using two large UK-based cohorts of individuals with rare neurodevelopmental conditions, the Deciphering Developmental Disorders study (DDD; N=7,955 patients with genotype array and exome sequence data) and the Genomics England 100,000 Genomes project (GEL; N=3,618 patients with genome sequence data), combined with several control cohorts (<u>**Supplementary Table 1**</u>). We have included a “Frequently Asked Questions” document in less technical language to explain the study, and to address some possible misunderstandings.

## Results

### GWAS meta-analysis for neurodevelopmental conditions reveals novel genetic correlations with other brain-related traits and conditions

We first sought to replicate the key findings from our previous GWAS for neurodevelopmental conditions^6^ in a large independent cohort. We identified a subset of GEL rare disease families with neurodevelopmental conditions and removed families overlapping with the DDD study (**<u>Methods</u>**). Almost all probands with neurodevelopmental conditions in GEL (97%) had intellectual disability or global developmental delay, versus 88% of those in DDD. The cohorts were broadly phenotypically similar (<u>**Extended Data Figure 2**</u>; **<u>Supplementary Note 1</u>**). To avoid spurious results due to population stratification, all genetic analyses were conducted in a genetically homogeneous subset of individuals with genetic similarity to British individuals from the 1000 Genomes Project ^36^, henceforth referred to as having GBR ancestry.

**Figure 2.**
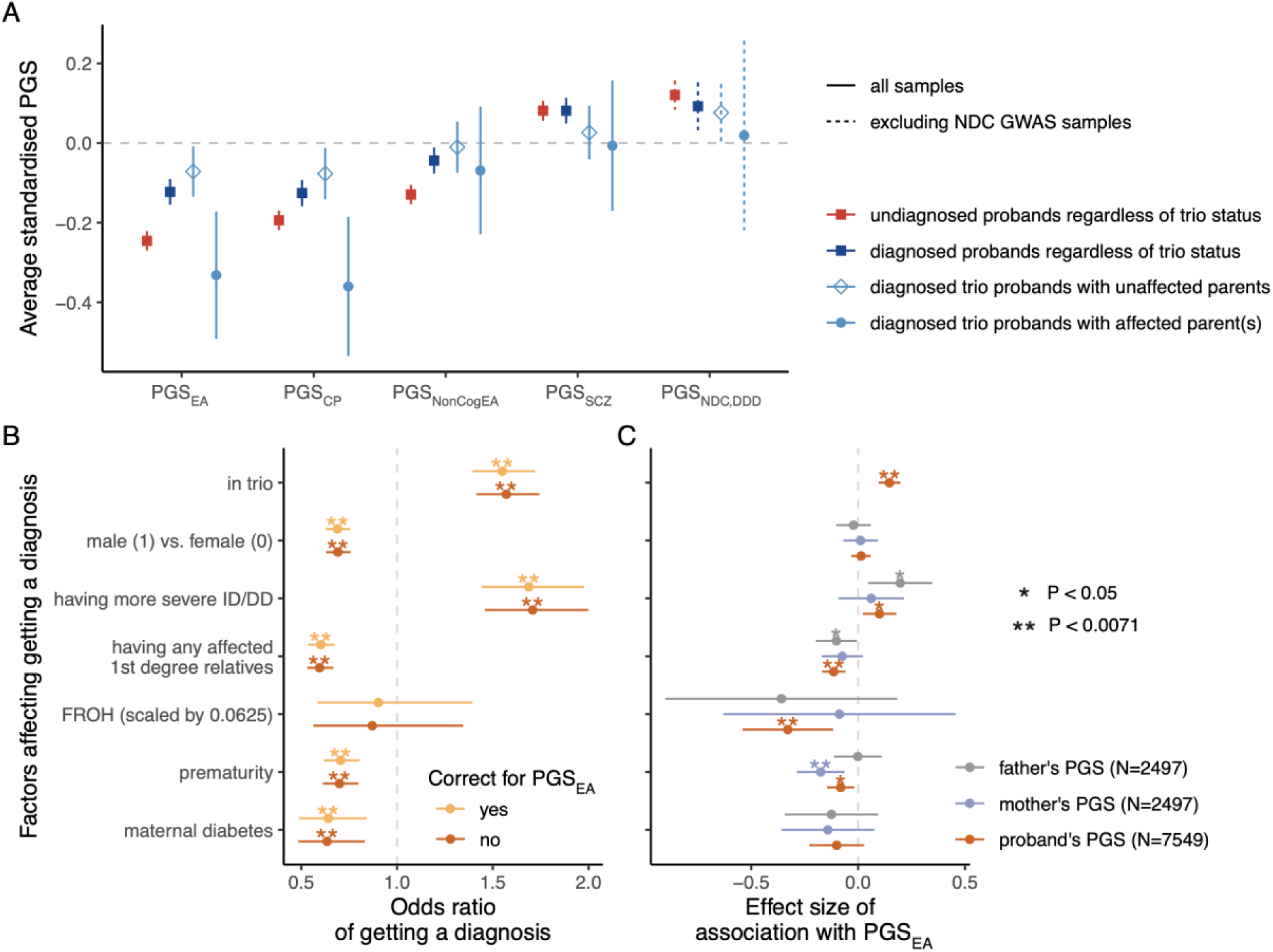
Disentangling polygenic score associations with diagnostic status. **A**) Average polygenic scores in probands with (“diagnosed”; N=3,821; dark blue) *versus* without (“undiagnosed; N=6,345; red) a monogenic diagnosis, from DDD and GEL combined. Subsets of diagnosed probands from trios are in light blue. The polygenic scores have been standardized such that the controls (UK Household Longitudinal Study+GEL for all polygenic scores except PGS_NDD,DDD_, for which only GEL controls were used) have mean 0 and variance 1. See **<u>Supplementary Table 5</u>** for results of statistical tests comparing the various groups. See also <u>**Extended Data Figure 4. B**</u>) Associations between various factors and diagnostic status within the full DDD cohort^5^, with or without correcting for the proband’s PGS_EA_, calculated within GBR-ancestry probands with neurodevelopmental conditions from DDD using logistic regression (see **<u>Supplementary Table 8</u>**). **C**) Associations between these factors and DDD probands’, mothers’ or fathers’ PGS_EA_, assessed via linear regression. Two asterisks indicate that the association passed Bonferroni correction for seven factors. Error bars show 95% confidence intervals. FROH: the fraction of the genome in runs of homozygosity; the expected value is 0.0625 for individuals whose parents are first cousins. EA: educational attainment; CP: cognitive performance; NonCogEA: the non-cognitive component of EA^42^; SCZ: schizophrenia; NDC,DDD: neurodevelopmental conditions, with the GWAS conducted in DDD *versus* the UK Household Longitudinal Study, and the polygenic score tested only in samples excluded from the GWAS (GEL and DDD Omni chip).

When comparing 3,618 unrelated patients with neurodevelopmental conditions to 13,667 unrelated controls within GEL, polygenic scores (PGSs) for educational attainment^37^, cognitive performance^37^, and schizophrenia^38^ each explained a significant but small amount of variance on the liability scale (<1%; logistic regression p<3.9x10^-4^). This was similar to that observed when comparing 6,397 unrelated patients from DDD with 9,270 independent unrelated controls (<u>**Supplementary Table 2**</u>). The polygenic score for neurodevelopmental conditions derived from our GWAS in DDD^6^ was also associated with neurodevelopmental conditions within GEL (p=1.1x10^-6^, R^2^=0.11% on the liability scale; <u>**Supplementary Table 2**</u>).

These results suggested that the polygenic contribution to rare neurodevelopmental conditions was similar between these two cohorts. Thus, to increase power to study common variant effects on these conditions, we conducted a GWAS in GEL, then meta-analyzed the results with the DDD GWAS (<u>**Extended Data Figure 3**</u>; **<u>Supplementary Data 1, 2 and 3</u>**). No single nucleotide polymorphism (SNP) passed genome-wide significance (p<5x10^-8^) in either DDD or GEL alone, but in the meta-analysis, six SNPs were significant in two independent loci on chromosomes 15 and 22, respectively (lead SNPs: rs113446150, p=4.0x10^-8^; rs2284084, p=1.7x10^-8^; **<u>Supplementary Note 2</u>**). Variants at one of these loci are associated with cognitive traits^39,40^. The fraction of phenotypic variance explained by genome-wide common variants - the SNP heritability - was estimated at between 3.7% (95% CI: 1.7–5.7%) and 11.2% (8.5–13.8%), depending on the method used (<u>**Supplementary Table 3**</u>).

**Figure 3.**
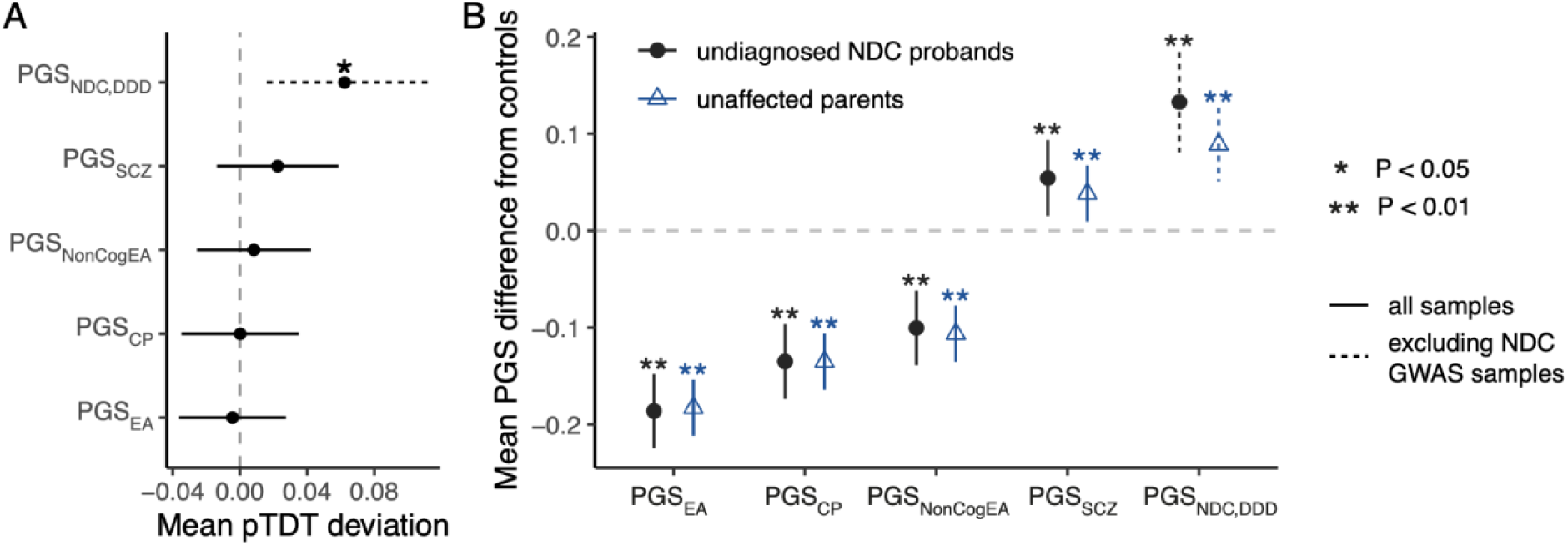
Polygenic background in parents of patients with neurodevelopmental conditions. **A)** Polygenic transmission disequilibrium test (pTDT) in undiagnosed probands with unaffected parents. We tested if probands’ polygenic score deviated from mean parental polygenic score in trios from GEL (N=1,343) and DDD (N=1,523, or N=224 for testing PGS_NDC,DDD_). Plotted is the mean pTDT deviation (difference between the child’s polygenic score and the mean parental polygenic score, in units of the SD of the latter), with error bars showing 95% confidence intervals. **B)** Mean polygenic score difference from control samples (GEL+UK Household Longitudinal Study, or GEL alone when testing PGS_NDC,DDD_). This includes only the samples in the trios used in the pTDT analysis. Error bars indicate 95% confidence intervals estimated from two-sided *t*-tests. See also <u>**Extended Data Figure 4**</u> and **<u>Supplementary Table 7</u>**.

To test for possible shared genetic contributors to rare neurodevelopmental conditions and other brain-related traits and conditions, we calculated genetic correlations (*r_g_*) between them using our own and published GWAS meta-analyses. We observed the expected negative genetic correlations between neurodevelopmental conditions and educational attainment ^37^ (EA; *r_g_*=- 0.65 [-0.84, -0.47], p=4.9x10^-12^) and cognitive performance ^37^ (CP; *r_g_*=-0.56 [-0.73, -0.39], p=1.6x10^-10^), stronger in magnitude than those observed with the DDD GWAS alone, and a positive genetic correlation with schizophrenia^38^ (SCZ; *r_g_*=0.27 [0.13, 0.40], p=9.7x10^-5^) (<u>**Figure 1A**</u>; <u>**Supplementary Table 4**</u>). Additionally, we detected significant genetic correlations (p<0.0038=0.05/13 traits) with several other mental health conditions including Attention-Deficit Hyperactive Disorder (ADHD)^41^ (*r_g_*=0.46 [0.28, 0.64], p=5.2x10^-7^), and with the non-cognitive component of educational attainment derived from GWAS-by-subtraction (NonCogEA) ^42^ (*r_g_*=- 0.37 [-0.52, -0.22], p=1.2x10^-6^) (<u>**Figure 1A**</u>). We hypothesized that the genetic correlations with mental health conditions could be explained at least in part by their relationship with educational attainment^42,43^, given the strong negative genetic correlation between that and neurodevelopmental conditions. To explore this, we used Genomic Structural Equation Modelling ^44^ (GenomicSEM) to re-estimate the genetic correlations while conditioning on the educational attainment GWAS summary statistics (<u>**Figure 1B**</u>). This significantly attenuated the genetic correlation with ADHD (*r_g_*=0.14 [-0.06, 0.34], p=0.18; two-sided z-test p=0.021 compared to unconditional *r_g_*), but the genetic correlations with the other conditions did not significantly change.

These results confirmed that common variants collectively associate with rare neurodevelopmental conditions in two independent cohorts, and that these common variant effects are shared with other brain-related conditions and cognitive traits. To further explore the contribution of polygenic background, below we used polygenic scores for neurodevelopmental conditions from the DDD-derived GWAS^6^ (PGS_NDC,DDD_) and for the most significantly genetically correlated traits (PGS_EA_, PGS_CP_, PGS_NonCogEA_, PGS_SCZ_) for which much larger GWASs and thus more powerful polygenic scores are available. All polygenic scores were corrected for principal components and standardized such that the controls from GEL and the UK Household Longitudinal Study have mean 0 and variance 1, except PGS_NDC,DDD_ which was standardized in the GEL controls alone. We note that several of these polygenic scores are significantly correlated with each other (<u>**Supplementary Figure 1**</u>), so, in the analyses below, our correction for multiples of five tests is conservative.

### Probands with monogenic diagnoses have less polygenic risk

Since 36% of patients in these cohorts have a molecular monogenic diagnosis (including *de novo*, recessive, X-linked or inherited dominant diagnoses), we next tested whether these diagnosed patients differed from undiagnosed patients in terms of their polygenic risk. Consistent with the liability threshold model (<u>**Extended Data Figure 1**</u>), we observed significantly higher PGS_EA_ (DDD and GEL combined; average difference Δ=0.12 SD, two-sided *t*-test p=3.0x10^-9^), PGS_CP_ (Δ=0.068 SD, p=1.2x10^-3^), and PGS_NonCogEA_ (Δ=0.085 SD, p=3.7x10^-5^) in probands with *versus* without a monogenic diagnosis (all passing Bonferroni significance i.e. p<0.05/5; <u>**Figure 2A**</u>). Despite this, we observed that for all polygenic scores except for PGS_NonCogEA_, the diagnosed probands still had significantly more polygenic risk than the controls (p<0.05/5; <u>**Figure 2A**</u>; <u>**Supplementary Table 5**</u>). Sensitivity analyses suggest that this observation is not driven by ascertainment bias in the controls, although the effect size is sensitive to the choice of control cohort, particularly for PGS_EA_ (**<u>Supplementary Note 3</u>**, <u>**Extended Data Figure 4**</u>, <u>**>Extended Data Figure 5, Supplementary Table 6**</u>). The difference between the diagnosed probands and controls is driven by those with affected parents (i.e. those reported by clinicians to show a similar phenotype to their child), who had significantly more polygenic risk for several traits than those with unaffected parents (e.g. PGS_EA_ Δ=0.26 SD, p=3.4x10^-3^) (<u>**Extended Data Figure 4; Supplementary Table 5**</u>). However, amongst undiagnosed probands, both those with affected parents and with unaffected parents showed significantly more polygenic risk than controls (<u>**Extended Data Figure 4**</u>, <u>**Supplementary Table 7**</u>).

**Figure 4.**
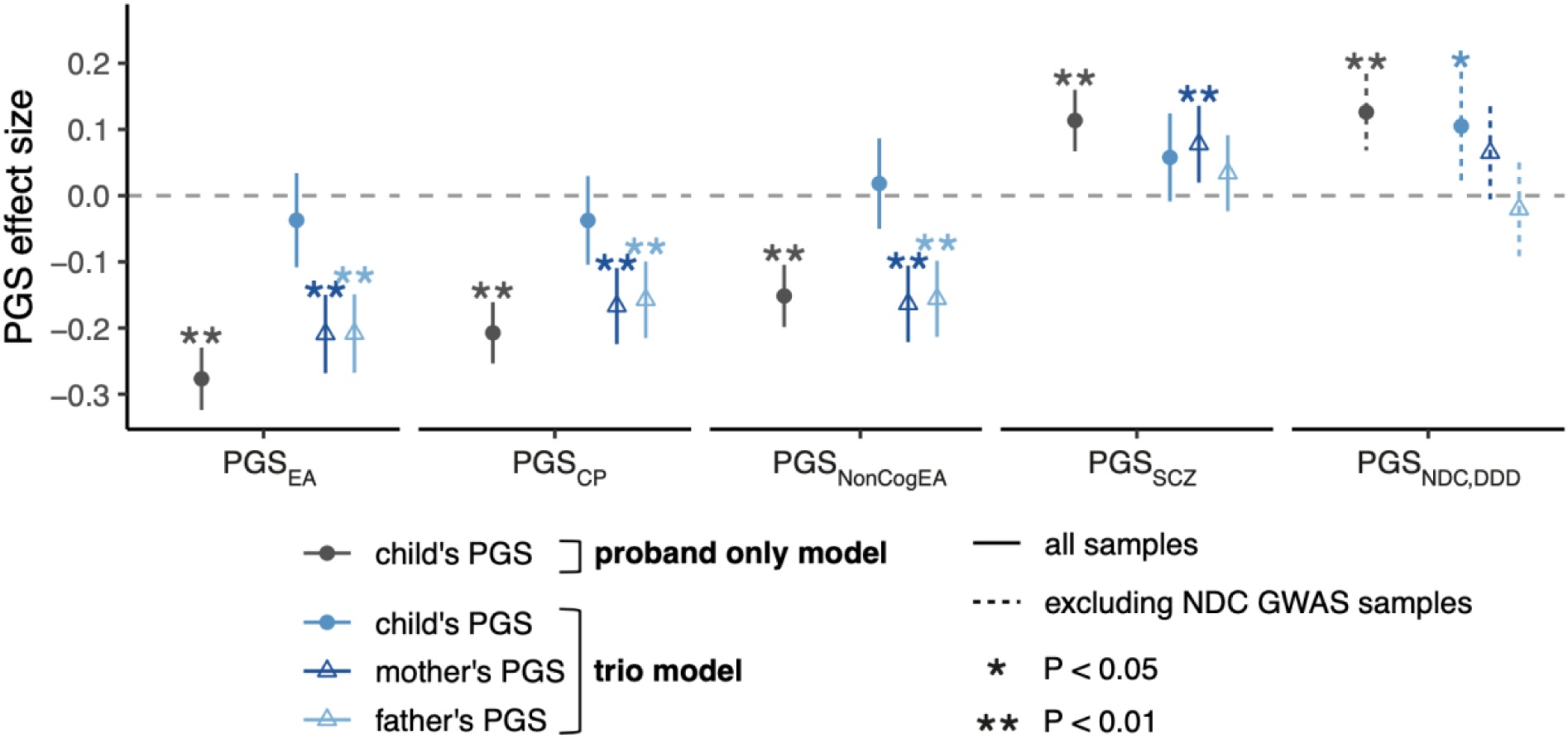
Regressions comparing undiagnosed probands with neurodevelopmental conditions to controls, with and without controlling for parental PGSs. The plot shows effect sizes of PGSs on case/control status, testing either the child’s PGS alone (“proband only”) amongst trio probands, or while additionally controlling for the parents’ PGSs (“trio model”). These were obtained from a logistic regression comparing undiagnosed proband with neurodevelopmental conditions from 2,866 trios in which parents are unaffected with 4,804 control trios from GEL (N=872), the Avon Longitudinal Study of Children and Parents (N=1,434) and the Millennium Cohort Study (N=2,498). Error bars indicate 95% confidence intervals.

**Figure 5.**
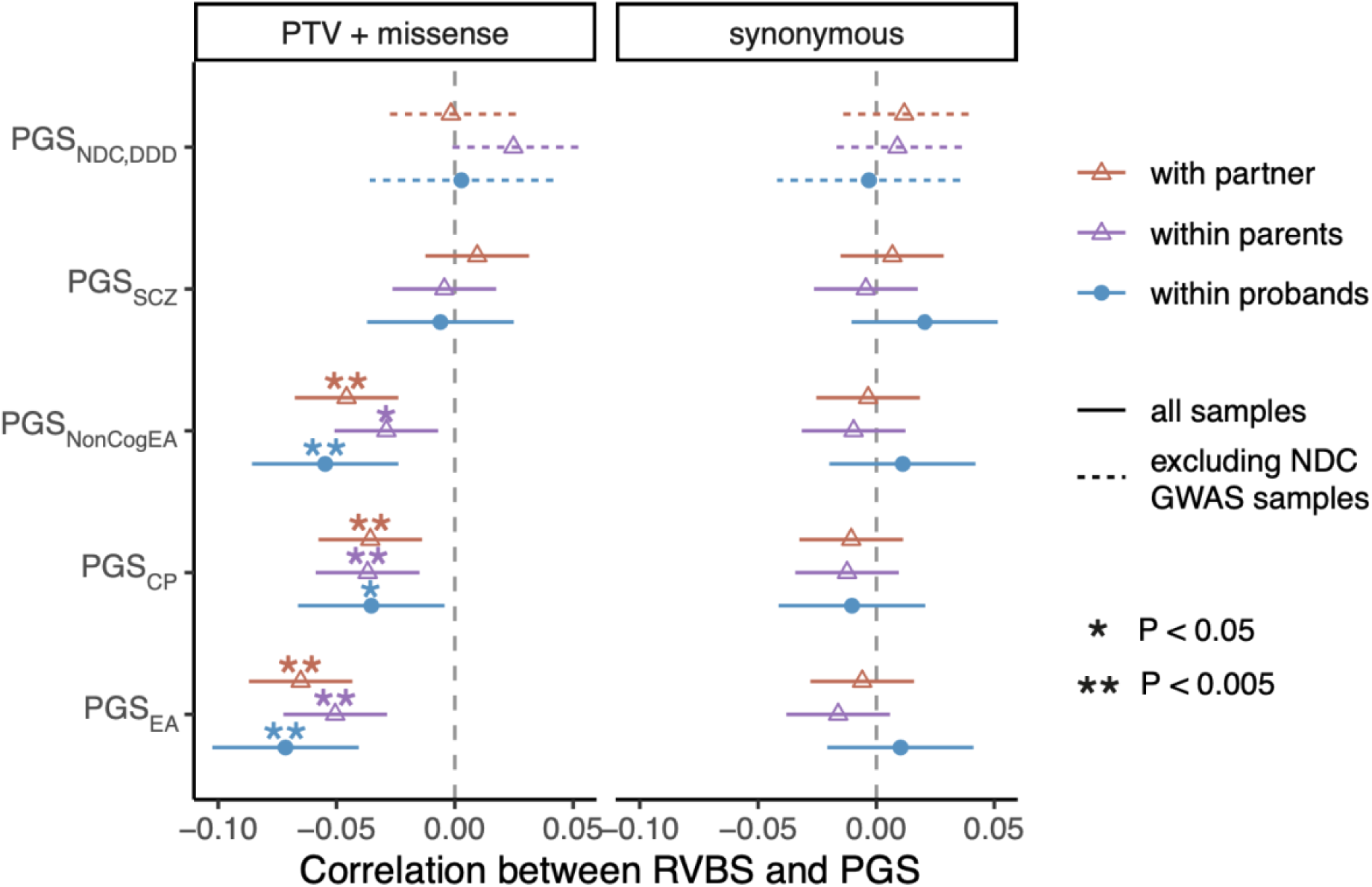
Correlation between rare variant burden scores and polygenic scores in patients with neurodevelopmental conditions and their parents. Correlation coefficients between the number of rare inherited rare damaging coding (left) or synonymous variants (right; negative control) in constrained genes and polygenic scores within/between different sets of individuals. In blue are the correlations within probands with neurodevelopmental conditions whose parents are unaffected (i.e. the child’s rare variant burden score, RVBS, with their own polygenic score, PGS), and in purple are the correlations within their parents. In orange is the cross-parental correlation i.e. one parent’s RVBS correlated with the other parent’s PGS. We calculated the correlations in a combined sample of trios with neurodevelopmental conditions from DDD and GEL (N=3,999 or 2,553 for PGS_NDC,DDD_ excluding samples from the original GWAS^6^). Note that both the RVBSs and PGSs have been corrected for 20 genetic principal components. Error bars represent 95% confidence intervals.

We next explored whether the difference in polygenic risk between diagnosed and undiagnosed probands was related to various technical, clinical and prenatal factors that are associated with receiving a monogenic diagnosis in DDD^5^ (<u>**Figure 2B**</u>). For example, diagnosed probands were more likely to be in a trio (probably due to the ability to distinguish *de novo* from inherited variants) and to have severe intellectual disability, and less likely to have been born prematurely (a known epidemiological risk factor for neurodevelopmental conditions^45–47)^ (<u>**Supplementary Table 8**</u>). We hypothesized that some of these associations might be confounding, or be confounded by the association between PGS_EA_ and diagnostic status, since, for example, single-parent households and premature birth are associated with higher levels of deprivation/lower parental educational attainment^48,49^. Indeed, we observed that the probands’ PGS_EA_ was significantly associated with several of these factors (<u>**Figure 2C**</u>): a higher chance of being in a trio and having more severe intellectual disability, and a lower chance of being born prematurely and having any affected first-degree relatives (**<u>Extended Data Figure 6</u>**). However, it was not associated with sex (**<u>Supplementary Note 4</u>**; **<u>Extended Data Figure 7</u>**) or maternal diabetes (<u>**Figure 2C**</u>; <u>**Supplementary Table 8**</u>). Controlling for PGS_EA_ minimally altered the association between these factors and diagnostic status (<u>**Figure 2B**</u>). Only a small part of the association between PGS_EA_ and diagnostic status was mediated by the effects of trio status (11%, 95% CI: 6.2–20.8%) and prematurity (3.1%, 95% CI: 0.4–7.3%). Thus, the observation that diagnosed patients tend to have lower polygenic risk than undiagnosed probably largely reflects the liability threshold model under which both common and rare variants contribute to risk (<u>**Extended Data Figure 1**</u>).

### Limited evidence for over-transmission of polygenic risk from unaffected parents to probands

Common variants are inherited from parents, and most of the parents in our sample are reported by clinicians to be clinically unaffected (89.2% in DDD and 95.4% in GEL, although the clinical annotation of parental affected status may be imperfect). Given this, and results in autism^50^, we hypothesized that probands without monogenic diagnoses inherit higher common variant risk for neurodevelopmental conditions from unaffected parents than one would expect given their parents’ mean risk. Applying the polygenic transmission disequilibrium test (pTDT)^50^ to undiagnosed trios with unaffected parents (<u>**Figure 3A**</u>) (1,343 in GEL plus 1,523 in DDD), we saw nominally significant over-transmission of PGS_NDC,DDD_ in 1,567 families not included in the original GWAS (pTDT deviation = 0.062; paired *t*-test p=0.014). This over-transmission was significant in females (pTDT deviation = 0.10, p=0.0078 in 589 trios) but not in males (pTDT deviation = 0.036, p=0.27 in 978 trios) (<u>**Extended Data Figure 7C**</u>; **<u>Supplementary Note 4</u>**). However, we saw no significant transmission disequilibrium for the other polygenic scores (paired *t*-test p>0.05), in either sex or in both sexes combined. Given the known over-transmission of PGS_EA_ to autistic individuals^50^, we excluded autistic individuals from our sample and repeated the pTDT, but conclusions were unchanged (<u>**Supplementary Figure 2**</u>). When focusing on probands with a monogenic genetic diagnosis, we saw no significant transmission distortion for any polygenic score tested (<u>**Supplementary Figure 3**</u>).

To put the pTDT results in context, we compared average polygenic scores between unaffected parents of undiagnosed patients and controls (<u>**Figure 3B**</u>; <u>**Supplementary Table 7**</u>). For all five polygenic scores tested, the parents had more polygenic risk than controls (two-sided *t*-test p<5.2x10^-6^ for all polygenic scores except PGS_SCZ_ which had p=0.0093). Given this observation and the results from the pTDT, we conclude that risk for neurodevelopmental conditions is affected both by familial polygenic background, or factors correlated with it, and by polygenic risk (specifically, PGS_NDC,DDD_) that is over-transmitted from unaffected parents to affected children.

### Non-transmitted common alleles in unaffected parents are associated with their children’s risk

We next explored one way in which familial polygenic background might affect children’s risk of neurodevelopmental conditions, namely indirect genetic effects, i.e. effects of alleles in parents on parental phenotypes that affect their offspring’s risk through the family environment. Indirect genetic effects have been argued to explain around ∼30-45% of the association between polygenic predictors of educational attainment and school grades^33,51^ and educational attainment^29,40,52^, although these inferences have been contested as confounded by parental assortment and population stratification^51,53^. To investigate the possible role of indirect genetic effects in risk of neurodevelopmental conditions, we compared 2,866 affected trio probands whose parents are unaffected with 4,804 control trios from two UK birth cohorts (N=3,932 trios) and from GEL (N=872 trios without neurodevelopmental conditions). Using logistic regression, we tested whether the children’s polygenic scores for traits related to neurodevelopmental conditions were significantly associated with case status (“proband only” model), and whether this held after conditioning on the parents’ polygenic scores (i.e. including all three trio members’ polygenic scores as covariates in the “trio model”)^54^ (<u>**Figure 4**</u>). The idea of this model is to isolate the environmentally-mediated portion of polygenic risk in the parents from the direct effects of alleles transmitted to their children. Following Young *et al*.^31^, we refer to the coefficients on the parental polygenic scores in the trio model as the “non-transmitted coefficients”, since they are mathematically equivalent to the coefficients on the polygenic score constructed from the non-transmitted alleles in a joint regression with the proband polygenic score (**<u>Methods</u>**).

For PGS_EA_, PGS_CP_, and PGS_NonCogEA_, we found that undiagnosed probands’ polygenic scores were no longer significantly associated with having a neurodevelopmental condition after conditioning on their parents’ polygenic scores in the trio model (implying limited or no direct genetic effects), whereas the non-transmitted coefficients were highly significant (<u>**Figure 4**</u>). This result held for PGS_EA_ and PGS_NonCogEA_ when analyzing trios with *versus* without neurodevelopmental conditions from GEL alone, and when using different combinations of control cohorts (<u>**Supplementary Figure 4**</u>); for PGS_CP_, results from these sensitivity analyses were more equivocal, but no evidence of direct genetic effects was seen. This finding could imply that there are aspects of the environment — including the prenatal environment — that are correlated with these non-transmitted alleles and that affect risk of neurodevelopmental conditions, including genetically-influenced parental phenotypes. However, our observations could also be due to the effects of parental assortment (i.e. phenotypic correlation between partners), which we discuss further below.

For PGS_NDC,DDD_, we found that the probands’ polygenic scores were still nominally significantly associated with having a neurodevelopmental condition after controlling for their parents’ polygenic scores in the trio model (<u>**Figure 4**</u>). This implies that there is a direct genetic effect of PGS_NDC,DDD_ on the probands’ risk of neurodevelopmental conditions, consistent with the over-transmission observed in <u>**Figure 3A**</u>. For schizophrenia, we saw no significant effect of the probands’ PGS_SCZ_ (p=0.089) in the trio model, whereas the mothers’ PGS_SCZ_ was significant (p=8.6x10^-3^). Thus, in summary, there is evidence for direct genetic effects of the polygenic score for rare neurodevelopmental conditions, but not for polygenic scores for related traits.

### Exploring the role of prenatal factors

We explored whether prenatal factors might mediate the effects of non-transmitted parental alleles on risk of neurodevelopmental conditions (**<u>Supplementary Note 5</u>**). Preterm delivery (i.e. giving birth prematurely)^55^, which is a risk factor for neurodevelopmental conditions in the offspring^45–47^, showed significant genetic correlations with lower educational attainment (r_g_=- 0.30 [-0.39, -0.21], p=2.3x10^-10^), mirroring the epidemiological association^56^, and with neurodevelopmental conditions (r_g_=0.58 [0.18, 0.97], p=0.004) (<u>**Extended Data Figure 8A**</u>, <u>**Supplementary Table 9**</u>). Premature birth was also associated with lower PGS_EA_ in DDD (<u>**Extended Data Figure 8B**</u>). In theory, the genetic correlation between educational attainment and premature delivery could reflect a causal effect of lower educational attainment on premature birth, and/or a causal effect of premature birth on lower educational attainment. Using Mendelian randomization, we found some evidence that lower educational attainment causally increases the risk of giving birth prematurely and of neurodevelopmental conditions (p=1.5x10^-5^ and p=6.5x10^-19^ respectively, using the inverse variance-weighted method; <u>**Extended Data Figure 9**</u>; **<u>Supplementary Note 5</u>**). The fact that the neurodevelopmental conditions of the kind studied in this paper are, by definition, childhood-onset, implies that individuals’ own educational attainment is unlikely to causally influence their risk of developing a condition; instead, our finding of a causal effect of educational attainment on these conditions is more likely to reflect a causal effect of parents’ educational attainment on their children’s risk, consistent with the presence of indirect genetic effects. However, we did not find significant evidence that prematurity explained the association between neurodevelopmental conditions and non-transmitted common variants in the parents that are associated with educational attainment (**<u>Supplementary Note 5</u>**; <u>**Supplementary Figure 5**</u>).

### Parental assortment obscures the true nature of common variant effects

Another factor that may contribute to the significant correlation between non-transmitted alleles in parents and neurodevelopmental conditions in their children is parental assortment, the phenomenon whereby people are more likely to choose partners with similar traits to themselves. Parental assortment is known to be particularly strong for educational attainment and cognitive ability, with estimates of phenotypic correlation between spouses ranging from 0.25 to 0.6^57–64^. It is also observed for psychiatric conditions^63,65–67^, including in parents of autistic individuals and of individuals with neurodevelopmental conditions due to the 16p12.1 deletion ^68^. One consequence of parental assortment is that it induces a correlation between alleles that act in the same direction on a trait, both between parents and, in their descendents, within and between loci^57^. Thus, parental assortment on cognitive ability or correlated traits (e.g. educational attainment) would be expected to lead to individuals with inherited rare variants associated with reduced cognitive ability^8,9,12,69^ also having a polygenic background of common variants associated with reduced cognitive ability^57,68^. In the context of our polygenic score analyses in <u>**Figure 4**</u>, in the proband-only model, the proband’s polygenic score would statistically capture (‘tag’) the correlated effects of these rare variants (which causally impact neurodevelopmental conditions^69,70^). However, in the trio model, the proband’s polygenic score would no longer be correlated with the rare variant component after conditioning on the parents’ polygenic scores, because the rare and common variant components segregate approximately independently within-family; instead, this correlation with the rare variant component would be reflected by the non-transmitted coefficients on the parents’ polygenic scores^53^.

To explore this potential genetic consequence of parental assortment in our cohorts, we tested whether the common and rare variant components contributing risk of neurodevelopmental conditions are indeed correlated. From the sequencing data in DDD and GEL, we extracted rare (MAF<1x10^-4^) protein-truncating variants (PTVs) and damaging missense variants in genes intolerant of loss-of-function (LoF) variation (“constrained genes”), which are associated with reduced cognitive ability^9^ and risk of neurodevelopmental conditions^69,70^. Consistent with the effects of parental assortment, amongst unaffected parents of probands with neurodevelopmental conditions, we observed that the number of rare damaging coding variants in constrained genes (the “rare variant burden score”, RVBS) in one parent was significantly negatively correlated with the other parent’s PGS_EA_ (*r*=-0.065, p=5.5x10^-9^), PGS_CP_ (*r*=-0.036, p=1.4x10^-3^) and PGS_NonCogEA_ (*r*=-0.046, p=4.3x10^-5^) (<u>**Figure 5**</u>), after correcting for genetic principal components. As expected, a similar correlation was seen within the probands themselves (<u>**Figure 5**</u>), regardless of whether including all probands, undiagnosed probands, or probands with *de novo* diagnoses, and if restricting RVBS to haploinsufficient genes associated with developmental disorders (<u>**Supplementary Figure 6**</u>). We also saw a similar result amongst control children from the Millennium Cohort Study, including after applying weights to adjust for non-random sampling and attrition in that cohort, indicating that this correlation is not only observed in patients with neurodevelopmental conditions (<u>**Supplementary Figure 7**</u>). We saw no significant correlation between any of the polygenic scores and the burden of rare synonymous variants in constrained genes or dominant genes associated with developmental conditions (<u>**Figure 5**</u>, <u>**Supplementary Figure 6**</u>), confirming that the result observed for deleterious variants is unlikely to be due to population structure artifacts. The correlations between polygenic scores and rare damaging variants may explain why we saw very limited evidence that these polygenic scores modify the penetrance of such variants in families with neurodevelopmental conditions (**<u>Supplementary Note 6</u>**, <u>**Supplementary Figure 8**</u>).

To explore whether the correlation between common and rare variants associated with neurodevelopmental conditions could be driving the association between non-transmitted common alleles and children’s risk shown in <u>**Figure 4**</u>, we extended the trio model to control for the probands’, mothers’ and fathers’ rare variant burden scores as well as polygenic scores (<u>**Extended Data Figure 10**</u>). This did not change our original conclusion from the trio regression, namely that the risk of neurodevelopmental conditions is correlated with non-transmitted common alleles in the parents that are associated with cognitive performance, educational attainment and the non-cognitive component thereof, but not with the transmitted common alleles. However, we cannot rule out that this association with non-transmitted common alleles is primarily driven by the assortment-induced correlation between common and rare variants, since the rare variant burden score we have used likely only captures a small proportion of the total rare variant component (just as the polygenic score only captures a small fraction of SNP heritability).

In summary, we find that parents’ non-transmitted alleles at common variants ascertained for their association with educational attainment and cognitive performance are correlated with their children’s risk of neurodevelopmental conditions, but we do not see evidence for direct genetic effects from transmitted alleles. Further work is needed to confirm whether this association with the non-transmitted alleles is due to true indirect genetic effects and/or parental assortment.

## Discussion

Here we combined two large cohorts of patients with rare neurodevelopmental conditions to explore the contribution of common variants to risk. After first demonstrating that polygenic scores for neurodevelopmental conditions and several related traits were significantly associated with case/control status within both DDD and GEL (<u>**Supplementary Table 2**</u>), we conducted a GWAS meta-analysis of patients with neurodevelopmental conditions from the two cohorts and revealed significant genetic correlations with several psychiatric conditions which had not been previously reported^6^ (**Figure 1A**). Conditional genetic correlations (<u>**Figure 1B**</u>) show that several of these (e.g. schizophrenia, Tourette’s) are not simply driven by the component of polygenic risk for neurodevelopmental conditions that is shared with educational attainment. This suggests that these mental health conditions share underlying biology with neurodevelopmental conditions that is independent of that captured by effects of common variants on educational attainment, although we acknowledge that estimates of genetic correlations can be biased by cross-trait parental assortment and other confounding factors^71^.

We showed that polygenic scores for several traits that are genetically correlated with neurodevelopmental conditions were significantly associated with having a monogenic diagnosis, with the strongest effect observed for educational attainment (<u>**Figure 2A**</u>). Our previous work had found no such difference in polygenic background between diagnosed and undiagnosed probands in DDD ^6^, and it is likely that power has been improved here by our larger sample size and better definition of which probands truly have a monogenic diagnosis ^5,27^. Our result is consistent with a liability threshold model for rare neurodevelopmental conditions; children without a large-effect monogenic variant may require higher polygenic load (or a major environmental contribution such as a teratogenic infection e.g. Zika virus) to move their phenotype over the threshold required to be clinically diagnosed with a neurodevelopmental condition (<u>**Extended Data Figure 1**</u>). Perhaps important for consideration in clinical settings, we find probands with more affected first-degree relatives had both a lower PGS_EA_ and a lower chance of getting a monogenic diagnosis in DDD (<u>**Extended Data Figure 6**</u>), emphasizing that if there are multiple first-degree relatives with neurodevelopmental conditions in a family, this may not necessarily be due to a monogenic cause. Our observation that diagnosed patients with affected parents (most of whom have inherited dominant diagnoses), and their parents, have lower average PGS_EA_ than those with unaffected parents (<u>**Extended Data Figure 4**</u>) is consistent with the effects of parental assortment (<u>**Figure 5**</u>).

Since most parents of the patients we studied are annotated as clinically unaffected, we hypothesized that they might be over-transmitting polygenic risk to their affected offspring. We saw nominally significant over-transmission of PGS_NDC,DDD_ from unaffected parents to undiagnosed probands, but saw no significant transmission distortion for PGS_EA_ or PGS_CP_ (<u>**Figure 3A**</u>), despite these polygenic scores explaining much more variance in risk than PGS_NDC,DDD_ (<u>**Supplementary Table 2**</u>). Consistent with this, in a two-generation model (<u>**Figure 4**</u>), we found evidence for a direct genetic effect of PGS_NDC,DDD_ on risk of neurodevelopmental conditions, but no evidence for direct genetic effects of the other polygenic scores tested. Instead, we observed that the parents’ PGS_EA_, PGS_CP_ and PGS_NonCogEA_ were significantly associated with their children’s risk even after controlling for the children’s PGS, indicating a correlation between non-transmitted alleles and the children’s phenotype. This may be due to indirect genetic effects and/or the consequences of parental assortment.

Previous papers have shown that non-transmitted alleles in the parents are associated with children’s educational attainment and school grades, explaining a third to a half of the overall association between educational outcomes and PGS_EA_ that is seen in population-based samples^29,33,34,51^. However, the interpretation of this finding is still a matter of debate, since most of these papers use models that can give spurious or inflated indirect genetic effect estimates due to population stratification and/or parental assortment^51,53,72^. Parental assortment induces a correlation between the polygenic score associated with the trait under assortment and the remaining genetic component of the phenotype with which the polygenic score would be uncorrelated under random assortment. This includes the component due to rare variants, which could have a much stronger effect on risk of neurodevelopmental conditions than the common variant component. We demonstrated (to our knowledge, for the first time) a correlation between the rare and common variant components affecting cognitive and educational outcomes, both between parents and within both offspring and parents (<u>**Figure 5**</u> and Supplementary Figure 7). This supports the hypothesis that the association of PGS_EA_ with lower risk of neurodevelopmental conditions is at least partly due to the assortment-induced correlation of PGS_EA_ with rare variants affecting both neurodevelopmental conditions and educational attainment. Although these observed correlations are small in magnitude (|*r*|<0.1), it is likely that the correlation between the total common and rare variant components of educational outcomes and neurodevelopmental conditions is substantially higher than this^53^, since only small fractions of these components are likely to be captured by the polygenic scores and our rare variant burden score, respectively. Very large whole-genome sequenced datasets will be required to better characterize the total rare variant component of these traits and estimate this correlation more accurately.

With the current study design, we were unable to demonstrate the presence of indirect genetic effects on risk of neurodevelopmental conditions unambiguously, and nor could we test whether, if present, these are mediated by parenting behaviors. However, we did explore whether common genetic variants might influence risk by affecting prenatal risk factors (a form of indirect genetic effects). We found that educational attainment showed a significant negative genetic correlation with preterm delivery, whereas neurodevelopmental conditions showed a significant positive genetic correlation with it even after conditioning on educational attainment with GenomicSEM (r_g_=0.47) (<u>**Extended Data Figure 8A**</u>). This is consistent with epidemiological studies that found an association between prematurity and poorer neurocognitive outcomes even after controlling for socioeconomic confounders^45,73–79^. We found some evidence from Mendelian randomization that lower educational attainment is causally associated with preterm delivery (<u>**Extended Data Figure 9B**</u>; **<u>Supplementary Note 5</u>**); this may be because lower educational attainment is associated with several factors that increase the risk of preterm delivery in the mother (such as a short inter-pregnancy interval^80^, exposure to tobacco smoke during pregnancy^81,82^, and pre-eclampsia^56^). We acknowledge that causal estimates from Mendelian Randomization analyses may be biased when using population-based GWASs, as we have done, so these findings should be considered tentative until confirmed using sufficiently well-powered within-family GWASs^32^. Although we did not find evidence for a causal effect of prematurity on neurodevelopmental conditions (**<u>Extended Data Figure 9C</u>**), several factors may have reduced the power of this analysis (**<u>Supplementary Note 5</u>**). We also saw no significant evidence that prematurity mediates indirect genetic effects of common alleles associated with educational attainment (**<u>Supplementary Note 5</u>**). However, it may be that our analysis was simply underpowered at this sample size, since we did see some attenuation (albeit not significant) of the non-transmitted coefficients for PGS_EA_ when removing premature probands (<u>**Supplementary Figure 5**</u>). Nonetheless, our results emphasize how genetics may confound epidemiological associations between risk factors and neurodevelopmental conditions^83,84^, and also suggest that studies seeking to characterize the nature of indirect genetic effects on educational outcomes should consider the contribution of prenatal factors.

Our study has several limitations. Firstly, the overall variance in risk of neurodevelopmental conditions explained by common variants is low (∼10%) and the polygenic scores tested here explain only a fraction of this. Having said that, these polygenic scores are statistically significant predictors of neurodevelopmental conditions (<u>**Supplementary Table 2**</u>) and are likely to explain more variance as GWAS sample sizes grow. Secondly, the reported significance of detected PGS effects does not simply reflect the strength of the real associations, but also the power of the original GWAS from which SNP effect sizes were derived. Thus, one must be cautious when comparing effects between polygenic scores for different traits. Thirdly, the phenotypic heterogeneity of the cohorts likely limits our power and may confound results. For example, missed diagnoses of autism amongst DDD and GEL participants with neurodevelopmental conditions (perhaps due to the young average age; **<u>Supplementary Note 1</u>**) could be confounding our result of there being no apparent under-transmission of PGS_EA_ (<u>**Figure 3A**</u>; <u>**Supplementary Figure 2**</u>), since PGS_EA_ may be over-transmitted to autistic individuals^26,50^ but under-transmitted to patients with intellectual disability who are not autistic. Fourthly, the fact that probands in trios tend to have higher polygenic scores for educational attainment than those not in trios (<u>**Extended Data Figure 5B**</u>) suggests that the trio probands are a non-random sample, which could potentially induce biases in trio-based analyses; for example, the undiagnosed trio probands may be enriched for monogenic causes in as-yet-undiscovered genes, which could reduce power when assessing over-transmission of polygenic risk. Additionally, many of our analyses are predicated on the assumption that the “unaffected parents” (i.e. those reported by the clinician not to have a similar phenotype to the proband) do not have phenotypes related to neurodevelopmental conditions. It may be that some fraction of them do have (or did have, earlier in life) relevant phenotypic features (e.g. learning difficulty, speech delay), but that these were not detected and recorded by the clinicians. The inclusion of these parents could be reducing power or confounding results in several analyses. Another caveat is that our estimates of effect size when comparing to controls are sensitive to the choice of control cohort, likely reflecting differences in educational-related ascertainment bias between them ^85^ (**<u>Supplementary Note 3</u>**; <u>**Extended Data Figure 5**</u>). Despite this, sensitivity analyses suggest that our main conclusions are robust and not driven by ascertainment bias of a particular control cohort (**<u>Supplementary Note 3</u>**; <u>**Extended Data Figure 4**</u>; <u>**Supplementary Figure 4**</u>; <u>**Supplementary Table 6**</u>). Finally, the correlation between the rare and common variant components of neurodevelopmental conditions (<u>**Figure 5**</u>), which is likely due to parental assortment, may have confounded several of these analyses.

In future, as GWAS discovery cohorts for both rare neurodevelopmental conditions and related traits increase in size, we will have more power to explore these common variant effects on risk, penetrance and phenotypic expressivity of these conditions. These studies should seek to confirm whether there really are no direct genetic effects of common variants influencing educational attainment and cognitive performance on risk of neurodevelopmental conditions, or whether these are just small. To disentangle the contribution of indirect genetic effects and parental assortment to common variant associations with neurodevelopmental conditions, future studies will need to use extended genealogies and/or more sophisticated modeling of the influence of parental assortment on common and rare variants than is currently possible ^51,53,72^. If these studies also had measures of epidemiological and prenatal risk factors such as prematurity, and of parental phenotypes and nurturing behaviors, one could explore how indirect genetic effects (if present) are mediated, which has potential implications for assessing the modifiability of risk. Finally, it will be important for future studies to explore the role of polygenic background in neurodevelopmental conditions in families with non-European genetic ancestries.

## Supporting information

FAQ and lay summaries

## Abbreviations

DDD: Deciphering Developmental Disorders study
GEL: Genomics England 100,000 Genomes Project
GWAS: genome-wide association study
GenomicSEM: Genomic Structural Equation Modelling
PGS: polygenic score
NDCs: neurodevelopmental conditions
EA: educational attainment
CP: cognitive performance
SCZ: schizophrenia
NonCogEA: the non-cognitive component of educational attainment
pTDT: polygenic transmission disequilibrium test
RBVS: rare variant burden score
UKHLS: United Kingdom Household Longitudinal Study
ALSPAC: Avon Longitudinal Study of Parents and Children
MCS: Millennium Cohort Study
QC: quality control
MAF: minor allele frequency
GSA: Global Screening Array from Illumina
SNP: single nucleotide polymorphism
HWE: Hardy-Weinberg Equilibrium.

## Acknowledgements

We are extremely grateful to families for their participation and engagement in the DDD study and 100,000 Genomes projects; without them, this research would not be possible. We also thank their clinicians and our colleagues (including the Sanger Human Genetics Informatics team, particularly Iaroslav Popov and Ruth Eberhardt) who assisted in the generation and processing of data. We are very grateful to Jillian Hastings-Ward, Hannah Podd and Hannah Humphrey from the Participant Panel for the 100,000 Genomes project, Ana Lisa Taylor Tavares from Genomics England, and Sarah Wynn from the patient organization Unique, for their assistance with writing the FAQ. We thank Athanasios Kousathanas and Loukas Moutsianas from Genomics England Bioinformatics Research Services for help with data quality control, Hilary Wong for useful discussions on prematurity, Michel Nivard for advice on use of GenomicSEM, and Angelica Ronald, Naomi Wray, and Nick Martin for helpful discussions.

DDD: The DDD study presents independent research commissioned by the Health Innovation Challenge Fund (grant no. HICF-1009-003). The full acknowledgements can be found at www.ddduk.org/access.html. This study makes use of DECIPHER, which is funded by the Wellcome Trust.

GEL: This research was made possible through access to data in the National Genomic Research Library, which is managed by Genomics England Limited (a wholly owned company of the Department of Health and Social Care). The National Genomic Research Library holds data provided by patients and collected by the NHS as part of their care and data collected as part of their participation in research. The National Genomic Research Library is funded by the National Institute for Health Research and NHS England. The Wellcome Trust, Cancer Research UK and the Medical Research Council have also funded research infrastructure.

UK Household Longitudinal Study: We used data from ‘Understanding Society: The UK Household Longitudinal Study’, which is led by the Institute for Social and Economic Research at the University of Essex and funded by the Economic and Social Research Council (grant number ES/M008592/1). The data were collected by NatCen and the genome-wide scan data were analysed by the Wellcome Trust Sanger Institute. Data governance was provided by the METADAC data access committee, funded by ESRC, Wellcome and MRC (grant number MR/N01104X/1).

ALSPAC: We are extremely grateful to all the families who took part in ALSPAC, the midwives for their help in recruiting them, and the whole ALSPAC team, which includes interviewers, computer and laboratory technicians, clerical workers, research scientists, volunteers, managers, receptionists and nurses. The UK Medical Research Council and Wellcome (Grant ref: 217065/Z/19/Z) and the University of Bristol provide core support for ALSPAC. This publication is the work of the authors and Hilary Martin will serve as a guarantor for the contents of this paper. Genome-wide genotyping data was generated by Sample Logistics and Genotyping Facilities at the Wellcome Sanger Institute and LabCorp (Laboratory Corporation of America) using support from 23andMe.

MCS: We are grateful to the Centre for Longitudinal Studies (CLS), UCL Social Research Institute, for the use of these data and to the UK Data Service for making them available. However, neither CLS nor the UK Data Service bear any responsibility for the analysis or interpretation of these data.

This research was funded in part by Wellcome (grant no. 220540/Z/20/A, “Wellcome Sanger Institute Quinquennial Review 2021–2026”). For the purpose of open access, the authors have applied a CC-BY public copyright license to any author accepted manuscript version arising from this submission. DB thanks the University of Cambridge Amgen Scholar Program for support.

## Author contributions

QQH and EMW conducted most of the analyses, with the remainder being conducted by PC and DSM. QQH and EMW carried out data preparation and quality control, with assistance from KES, VKC, PD, SL, TM, MKM, SA and DB. ES, CFW and HVF helped supervise the DDD study, together with MEH. EJR, VW, ASY and MEH provided key intellectual input. HCM supervised the analyses and directed the study. QQH, EMW, and HCM wrote the first draft of the manuscript, with input from PC, DSM, JCB, VW, ASY and MEH. All authors read and commented on the final manuscript.

## Online Methods

### Cohort Descriptions and phenotypes

#### Deciphering Developmental Disorders (DDD)

The aim of the DDD study is to find molecular diagnoses for families and patients affected by previously genetically undiagnosed, severe developmental conditions. Recruitment was conducted from 2011 to 2015 across twenty-four clinical genetics services in the United Kingdom (UK) and Ireland^86^. The DDD study has UK Research Ethics Committee approval (10/H0305/83, granted by the Cambridge South Research Ethics Committee and GEN/284/12, granted by the Republic of Ireland Research Ethics Committee). The clinical inclusion criteria included neurodevelopmental conditions, congenital, growth or behavioral abnormalities and dysmorphic features. Probands were systematically phenotyped via DECIPHER^87^ using Human Phenotype Ontology (HPO)^88^ terms and a bespoke online questionnaire that collected information on developmental milestones, growth measurements, number of affected relatives, prematurity, maternal diabetes, and other clinically-relevant parameters. The cohort has been described extensively^5,70,86,89^.

We focused on probands in the DDD cohort who had neurodevelopmental conditions (NDCs), which were defined previously by Niemi *et al*.^6^. Briefly, these were probands who had at least one of the following neurodevelopmental HPO terms or their descendant terms: abnormality of higher mental function (HP:0011446), neurodevelopmental abnormality (HP:0012759), abnormality of the nervous system morphology (HP:0012639), behavioural abnormality (HP:0000708), seizures (HP:0001250), encephalopathy (HP:001298), abnormal synaptic transmission (HP:0012535), or abnormal nervous system electrophysiology (HP:0001311).

#### Genomics England (GEL) 100,000 Genomes Project

The 100,000 Genomes project is an initiative by the UK Department of Health and Social Care to whole-genome sequence individuals with rare conditions and cancer in the National Health Service^90,91^. The 100,000 Genomes project was approved by the East of England—Cambridge Central Research Ethics Committee (REF 20/EE/0035). The rare disease branch of the project consists of sequencing data from ∼72,000 patients with rare conditions and their relatives, in ∼34,000 families with a variety of structures. There are over 190 rare conditions represented in the cohort, and about 23% of the patients have NDCs. The cohort was sequenced at around 35x coverage, and variant calling and quality control (QC) were performed by Genomics England^91,92^.

GEL NDC patients were defined as those recruited under the “Neurodevelopmental disorders” disease sub-category, or with more than one HPO term that was a descendent of “Neurodevelopmental Abnormality” (HP:0012759). We removed probands whose age of onset was >16 years or who had neurodegenerative conditions.

The set of unrelated GEL controls included cancer patients over 30 years old (N=10,469) and unaffected relatives (N=3,198) of probands with rare conditions who were not in the NDC set and did not have phenotypes similar to probands from DDD (“DDD-like”). The “DDD-like” probands were defined as those who:

1) were recruited into a disease model which was also used to recruit probands who had previously been recruited into DDD (see section below on identifying probands overlapping between the two cohorts), or
2) had one the top five HPO terms used in DDD and their descendants, namely HP:0000729 (autistic behaviour), HP:0001250 (seizure), HP:0000252 (microcephaly), HP:0000750 (delayed speech and language development) and HP:0001263 (global developmental delay).

Probands recruited into the neurodegenerative disorders subcategory or with an age of onset >16 years were removed from the DDD-like set, as were probands recruited into a disease subcategory for which the average age of probands was >16 years.

To define relatedness, we used a file generated by GEL consisting of a pairwise kinship matrix produced using the PLINK2^93,94^ implementation of the KING robust algorithm^95^ and a --king-cutoff of 0.0442 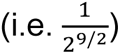.

#### Control cohorts

The UK Household Longitudinal Study (UKHLS) cohort consists of a continuation of the British Household Panel Survey (BHPS) of individuals living in the UK^96,97^. The Avon Longitudinal Study of Parents and Children (ALSPAC) is a birth cohort study of children born in Avon, England with expected dates of delivery between 1st April 1991 and 31st December 1992^98^. Eligible pregnant women (N=13,761) were recruited and their children have been phenotyped extensively over the last 30 years. Ethical approval for the study was obtained from the ALSPAC Ethics and Law Committee and the Local Research Ethics Committees. Please note that the study website (http://www.bristol.ac.uk/alspac/researchers/our-data/) contains details of all the data that is available through a fully searchable data dictionary and variable search tool. The Millennium Cohort Study (MCS) is a birth cohort study of children born across the UK during 2000 and 2001 from 18,552 families ^99,100^. Further information about recruitment of these cohorts is given in **<u>Supplementary Note 3</u>**.

### Preparation of genetic data

Individuals from DDD, UKHLS, ALSPAC, and MCS were genotyped on various arrays, whereas GEL individuals were whole-genome sequenced. The available data are summarized here briefly:

● A subset of the DDD cohort (all children and several thousand parents) was genotyped on three genotype array chips: the Illumina HumanCoreExome chip (CoreExome), the Illumina OmniChipExpress (OmniChip), and the Illumina Infinium Global Screening Array (GSA). Some probands were genotyped on more than one chip, as shown in <u>**Supplementary Figure 9**</u>. In downstream analysis, we used the CoreExome and OmniChip data for analyses of probands, and the GSA and OmniChip data for analyses of trios. QC of CoreExome (including DDD patients and 9,270 UKHLS controls genotyped on the same chip) and Omnichip data were performed by Niemi *et al*. ^6^ and we performed QC in the GSA data specifically for this paper (**<u>Supplementary Tables 10 and 11</u>**). The DDD cohort was also exome sequenced, and those data were used for the analyses involving rare variants. QC and processing of the exome data are described below in the section “Extracting and annotating rare variants”.
● GEL individuals were whole genome sequenced with 150bp paired-end reads using Illumina HiSeqX. Variant calling and QC were performed by Genomics England. We used 78,195 post-QC germline genomes from the Aggregated Variant Calls (aggV2) prepared by the GEL team. We kept variants that passed the QC filters shown in <u>**Supplementary Table 12**</u>.
● Data we received from ALSPAC were processed in two batches^97^. In the first batch, we received post-QC array data for G0 mothers (N=8,884) who were genotyped on the Illumina Human 660W chip and G1 children (N=8,932) genotyped on the HumanHap550 quad chip. In the second batch, we received another 2,198 parents (G0 mothers and G0 partners^101^) who were genotyped on the CoreExome array.
● We received data for 21,181 MCS samples who were genotyped using the GSA array chip^102^.

We applied standard QC filters in each dataset separately, described further in <u>**Supplementary Methods**</u>.

#### Genetically predicted ancestry

The **<u>Supplementary Methods</u>** provide detailed information on ancestry inference, but we summarize it briefly here. The identification of GBR-ancestry samples from the DDD CoreExome and OmniChip data was described previously^6^. To identify individuals of genetically inferred GBR ancestry in DDD GSA samples, we first projected post-QC samples onto 1,000 Genomes phase 3 individuals^36^ (<u>**Supplementary Figure 10**</u>). We then performed another principal component analysis (PCA) within the loosely defined European ancestry subset and identified a homogeneous subgroup (<u>**Supplementary Figure 11**</u>) using Uniform Manifold Approximation and Projection (UMAP)^103^. Since we merged parent-offspring trios genotyped on GSA and Omnichip array chips in downstream analysis, we kept GSA individuals who were similar to Omnichip individuals in terms of genetic ancestry in PCA space (<u>**Supplementary Figure 12**</u>). In GEL, we used individuals with genetically inferred European ancestry, which were identified by the GEL bioinformatics team. We further restricted to a homogeneous subset (N=56,249) that represents white British individuals (<u>**Supplementary Figure 13**</u>). Array data received from the ALSPAC all had genetically predicted European ancestry, so we did not perform any filtering based on genetic ancestry. We performed similar PCA and UMAP clustering to identify GBR-ancestry individuals in MCS (<u>**Supplementary Figure 14**</u>; <u>**Supplementary Figure 15**</u>), and further filtered to individuals who self-reported as being of White ethnicity.

#### Identifying and removing relatives within and across cohorts

Within each dataset, we identified up to third-degree relatives (kinship coefficient > 0.0442 by KING v2.2.4^95^) using post-QC genotyped array data or WGS data. We always used a subset of unrelated individuals (i.e. more distant than third-degree relatives) in downstream analysis. In analyses using trios, we made sure probands in trios were unrelated and parents were unrelated with parents from other families.

In analyses combining DDD and GEL, we removed from GEL any participants who were also recruited into DDD and or who were related to DDD participants, and also removed Scottish samples from DDD since we were unable to check whether GEL samples were related to them (**<u>Supplementary Methods</u>**). We removed individuals from the two birth cohorts who were related to each other or to DDD participants, which left 1,434 and 2,498 trios from ALSPAC and MCS, respectively (**<u>Supplementary Methods</u>**).

#### Imputation and post-imputation QC

Imputation of array data was performed in each genotyped cohort separately using the maximum number of variants available after QC. Prior to imputation, we removed palindromic SNPs, SNPs that were not in the imputation reference panel, and SNPs with mismatched alleles. DDD samples and UKHLS controls who were genotyped on the CoreExome array were imputed with the HRC r1.1 reference panel by Niemi *et al.*^6^. DDD GSA and Omnichip samples and ALSPAC samples were imputed to the TOPMed r2 reference panel using the TOPMed imputation server, and the MCS samples to the HRC r1.1 reference panel ^104–106^. We kept well imputed common variants with Minimac4 R^2^ >0.8 and minor allele frequency (MAF) >1%. For polygenic score analyses, we subsequently restricted to common variants that passed these QC filters in all genotyped cohorts and also passed QC in the GEL WGS data.

### Defining patients with *versus* without monogenic diagnoses

#### DDD

The DDD study identified clinically relevant rare variants from exome sequencing and chromosome microarray data using a filtering procedure described in Wright *et al.*^86^. The procedure focuses on identifying rare damaging variants that fit an appropriate inheritance mode in a set of genes that cause developmental disorders (DDG2P, https://www.deciphergenomics.org/ddd/ddgenes). Variants that pass clinical filtering are uploaded to DECIPHER^87^, where the patients’ clinicians are asked to classify them as definitely pathogenic, likely pathogenic, uncertain, likely benign or benign. We defined “diagnosed” probands as those with one or more variants either annotated as pathogenic/likely pathogenic in DECIPHER by their referring clinician, or predicted as pathogenic/likely pathogenic using autocoded ACMG diagnoses as described in ^5^. All remaining probands were classed as “undiagnosed”. Probands with a *de novo* diagnosis are those with a *de novo* mutation in a monoallelic or X-linked DDG2P gene that was either annotated or predicted as pathogenic/likely pathogenic.

#### GEL

The probands assigned diagnostic status were those included in the Genomic Medicine Service exit questionnaire, in which a clinician evaluated the pathogenicity of variants of interest identified through GEL’s custom pipeline. We defined “diagnosed” probands as those that had a pathogenic or likely pathogenic variant that is annotated as partially or fully explaining their phenotype in this exit questionnaire. Probands with a *de novo* diagnosis are those whose pathogenic/likely pathogenic variants from the exit questionnaire were annotated as *de novo* protein truncating or missense variants in DDG2P monoallelic or X-linked genes. We defined “undiagnosed” probands as those that were present in the exit questionnaire but not annotated as having a pathogenic or likely pathogenic variant and not annotated as “yes” or “partially” in the “case_solved_family” column. We further removed from this undiagnosed set any probands who have potential diagnoses in the Diagnostic Discovery data in GEL, which is a list of variants submitted by researchers that are thought likely to be pathogenic by the GEL clinical team.

### Extraction and quality control on rare variants

Quality control of DDD exome sequencing data and extraction of rare single nucleotide variants (SNVs), and insertion and deletions (indels) is summarized in <u>**Supplementary Table 13.**</u> Indels in the same gene and sample were removed (4% of indels with MAF < 1%), since these were often part of complex mutational events that would require haplotype-aware annotation.

For GEL, details of the QC of SNVs and indels in the WGS data are provided by the GEL team91,92 and variant QC is summarized in <u>**Supplementary Table 12**</u>. We use a custom python script to extract rare variants from GEL aggregated WGS variant call format files (aggV2). We filtered genotypes to those with genotype quality (GQ) ≥ 20 and read depth (DP) ≥ 10. We removed heterozygous genotypes that did not pass a binomial test of balanced REF and ALT alleles (p < 1x10^-3^) or for which ALT/(REF+ALT) (AB ratio) was not between 0.2 and 0.8. We further removed variants with missing high quality genotypes in more than 5% of all samples in aggV2. We removed indels in the same gene and sample for the same reason described above for DDD.

For MCS, details of the QC of exome sequencing data are in <u>**Supplementary Methods**</u>.

### Defining trio sample sets in DDD and GEL

The procedure used for filtering trios used in DDD and GEL is shown in <u>**Supplementary Figure 16**</u>. Briefly, in DDD, we combined data across GSA and OmniChip arrays and kept trios in which all three members had GBR ancestry and the proband had an NDC. We excluded trios recruited from Scottish centres and kept unrelated trios. We then split trios into those with both parents unaffected and those with one or both parents affected. These were then categorized as genetically diagnosed or undiagnosed. We applied similar filtering in GEL trios. See **<u>Supplementary Methods</u>**for more information.

### GWAS of neurodevelopmental conditions in GEL and meta-analysis with DDD

We used PLINK v1.9 to conduct a GWAS comparing individuals with NDCs (N=3,618) to controls (N=13,667) in GEL, controlling for 20 genetic principal components (PCs), age, and sex. Prior to running the GWAS, we removed variants with MAF < 1%, missingness > 2% or Hardy-Weinberg equilibrium (HWE) P-value < 1x10^-5^, and performed a differential missingness test between the NDC patients and controls and removed variants with p-value < 1x10^-5^. We repeated the GWAS comparing DDD patients with neurodevelopmental conditions on the CoreExome array (N=6,397) to UKHLS controls (N=9,270) using PLINK v1.9, after excluding DDD patients recruited from Scottish centres.

We used METAL^107^ to conduct an inverse variance-weighted GWAS meta-analysis between the DDD-UKHLS and GEL NDC GWASs. We removed palindromic SNPs with MAF > 0.4 since the strand could not be easily inferred using MAF. We also excluded SNPs with discordant AF (difference > 0.05) between the two cohorts. This left 5,451,801 overlapping SNPs in the meta-analysis.

### Heritability and genetic correlations

We used Linkage Disequilibrium Score Regression (LDSC)^108^ to estimate SNP heritability using summary statistics from the GWAS of NDCs in DDD, in GEL, and a meta-analysis of the two cohorts. We used ∼1 million common SNPs from HapMap3 with precomputed LD scores. SNP heritability on the liability scale was estimated assuming a cumulative population prevalence of 1% for rare neurodevelopmental conditions^6^. We used the effective sample size (4/(1/N_cases_ + 1/N_controls_)) or the sum of two effective sample sizes for the meta-analysis and a sample prevalence of 50% in LDSC, as recommended previously^109^. In addition, we also applied two methods to estimate SNP heritability using individual-level data in DDD and GEL separately. We performed GREML-LDMS^110^ stratified by LD (two bins of equal size) and MAF (three bins: 1%–5%, 5%–10%, >10%). We also ran phenotype correlation–genotype correlation (PCGC) regression^111^, using the LDAK-Thin Model to compute the kinship matrix using the direct method. We corrected for sex, and ten genetic principal components as covariates in both methods. We then meta-analyzed the SNP heritability estimates from DDD and GEL using an inverse-variance weighted method.

We used LDSC to estimate genetic correlations between the DDD NDC GWAS or the meta-analyzed NDC GWAS and various brain-related traits and conditions listed in <u>**Supplementary Table 14**</u>. We did not use the GEL NDC GWAS to calculate genetic correlations as the SNP heritability was not significantly different from zero according to LDSC.

### Estimating conditional genetic correlations with GenomicSEM

To estimate the genetic correlations between various traits/conditions (<u>**Supplementary Table 14**</u>) and NDCs independent of cognitive performance or educational attainment signals, we used genomic structural equation modelling (GenomicSEM)^42,44^. We estimated the genetic correlation between the target trait and a latent variable representing the non-cognitive component of NDCs, which was genetic influences on NDCs that were not explained by cognitive skills. We applied the GenomicSEM model without SNP effects. We also estimated genetic correlation with the “non-educational attainment” latent variable, which represented genetic influences on NDCs that were not accounted for by the educational attainment latent variable.

### Calculating polygenic scores

For calculating PGSs, we used the set of SNPs that were well-imputed in all array cohorts (Minimac4 R^2^ > 0.8), passed QC in GEL aggV2 samples, and had MAF >1% in all cohorts. We used LDPred^112^ to estimate weights for calculating PGSs and an LD reference panel composed of HapMap3^113^ common variants based on 5,000 unrelated individuals of white British genetically-inferred ancestry from the UK Biobank^114^ (**<u>Supplementary Methods</u>**). GWAS summary statistics for years of schooling (a measure for EA)^37^, the non-cognitive component of educational attainment (NonCogEA)^42^, cognitive performance (CP)^37^, schizophrenia (SCZ)^38^, and NDCs^6^ were matched with the list of overlapping SNPs (<u>**Supplementary Table 14**</u>). PGS_NDC,DDD_ was evaluated in the DDD Omnichip samples and the GEL samples which were not in the DDD GWAS. To make PGSs comparable across cohorts (DDD, GEL, UKHLS, MCS and ALSPAC), we performed a joint PCA across all cohorts and adjusted the raw scores for 20 PCs. For all analyses, residuals were scaled so that the combined set of unrelated control samples from GEL and UKHLS (or GEL controls only for PGS_NDC,DDD_) had mean = 0 and SD = 1.

### Analyses of polygenic scores

#### Evaluating variance explained by the PGSs

We evaluated how much variance in risk of NDCs was explained by the PGS on the liability scale^111,115,116^. We compared 6,397 NDC probands from DDD to 9,270 controls from UKHLS, and 3,618 NDC probands from GEL to 13,667 GEL controls defined as described above. We assumed the population prevalence of NDCs to be 1%^6^.

#### Comparison of PGS between different subsets of probands and parents

We used two-sided *t*-tests to compare PGSs between different groups of probands, parents and controls seen in <u>**Figure 2A**</u>, <u>**Figure 3B**</u>, <u>**Extended Data Figure 4**</u>, <u>**Extended Data Figure 5**</u>, and <u>**Supplementary Tables 5, 6 and 7**</u>. We report the mean difference in PC-corrected PGS between groups. Groups who were compared with each other include:

● Combined set of controls from GEL and UKHLS
● Control individuals from UK birth cohorts, ALSPAC and MCS
● Undiagnosed NDC probands regardless of trio status
● Diagnosed NDC probands regardless of trio status
● Undiagnosed NDC probands for whom both parents are unaffected
● Unaffected parents of undiagnosed NDC probands
● Undiagnosed NDC probands with one or both parents affected
● Affected parents of undiagnosed NDC probands
● Diagnosed NDC probands for whom both parents are unaffected
● NDC probands with *de novo* diagnoses for whom both parents are unaffected
● Unaffected parents of diagnosed NDC probands
● Diagnosed NDC probands with one or both parents affected
● Affected parents of diagnosed NDC probands

The sample size of each subset is listed in <u>**Supplementary Table 1**</u>. We excluded controls from UKHLS as well as DDD CoreExome and GSA probands when testing the DDD-derived NDC PGS (since these had been included in the original NDC GWAS, whereas the individuals genotyped on the Omnichip had not). All the *t*-tests involving NDC probands were performed in samples from DDD and GEL combined.

We also compared female probands *versus* male probands without a monogenic diagnosis regardless of trio status (2,427 and 1,574 male probands from DDD and GEL, and 1,426 and 918 female probands from DDD and GEL), and unaffected mothers versus unaffected fathers (1,523 trios from DDD and 1,343 trios from GEL) using two-sided *t*-tests (<u>**Extended Data Figure 7AB**</u>).

#### Associations between PGS and diagnostic status

We compared average PGSs in NDC probands with and without a monogenic diagnosis using two-sided *t*-tests, combining NDC probands from DDD and GEL regardless of whether they were in a trio or not. We compared NDC subgroups to the combined control set from UKHLS and GEL, as well as to unrelated children from the MCS cohort who were reweighted using available sociodemographic data to make them more representative of the general UK population (**<u>Supplementary Note 3</u>**).

Within DDD (N=7,549 without excluding Scottish samples or samples who were related to GEL participants), we tested whether the proband’s PGS_EA_ was associated with factors affecting getting a diagnosis in linear regression models:

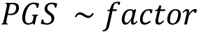

We investigated the following binary factors: trio status (N=5,507 with both parents exome sequenced but not necessarily genotyped), proband sex (N=4,421 male probands), whether the proband had any affected first-degree relatives (N=1,623), whether the proband was born preterm (N=1,098 with gestation <37 weeks), whether the mother had diabetes (N=242), and whether the proband had severe intellectual disability or developmental delay (ID/DD; N=941) *versus* mild or moderate ID/DD (N=1,887). We compared probands with the above mentioned characteristics to all other probands, except when comparing probands with severe *versus* mild or moderate ID/DD for which we excluded probands without ID/DD or with ID/DD of unknown severity. We also investigated a continuous factor, the degree of consanguinity, quantified by the fraction of the genome in runs of homozygosity (F_ROH_) divided by 0.0625, which is the expected fraction given a first-cousin marriage.

We also tested whether the mother’s or father’s PGS_EA_ was associated with the above factors, in a total of 2,497 samples; we did not test for association with trio status since parental genotype data were only available for full trios anyway.

See the <u>**Supplementary Methods**</u> for a description of estimation of the odds ratio of diagnosis for different configurations of affected relatives shown in <u>**Extended Data Figure 6**</u>, and of the mediation analysis to explore whether trio status and prematurity were mediating the association between PGS_EA_ and diagnostic status.

#### Evaluating over-transmission of PGS: the polygenic transmission disequilibrium test

We conducted polygenic transmission disequilibrium tests (pTDT) in undiagnosed and diagnosed probands from DDD (N=1,523 undiagnosed, 443 diagnosed) and GEL (N=1,343 undiagnosed, 507 diagnosed) combined. We also conducted pTDT in these trios excluding autistic probands.

The pTDT is a two-sided one-sample *t*-test of the probands’ PGS deviation from expectation, which is their parents’ mean PGS. The pTDT deviation is defined as:

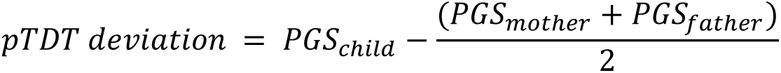

To evaluate whether the pTDT deviation is significantly different than 0, the pTDT test statistic (𝑡_𝑝𝑇𝐷𝑇)_ is defined as:

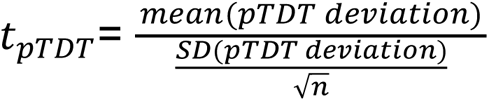

#### Analyses of non-transmitted coefficients

We evaluated direct genetic effects and effects of non-transmitted common alleles on NDC case status using logistic regression on PGSs:

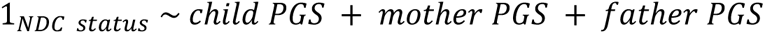

where 1_𝑁𝐷𝐶 𝑠𝑡𝑎𝑡𝑢𝑠_is an indicator variable for whether the individual is an NDC case (1) or control (0).

Since a child’s PGS is calculated using transmitted alleles and the difference between the sum of parents’ PGS and the child’s PGS is equivalent to a PGS derived from non-transmitted alleles, this model can be rewritten as:

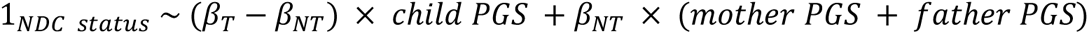

where 𝛽_𝑁𝑇_ indicates the non-transmitted coefficient and 𝛽_𝑇_ indicates the coefficient on transmitted alleles. The regression coefficient on child PGS in this trio model represents an unbiased estimate of direct genetic effect (difference between 𝛽_𝑇_ and 𝛽_𝑁𝑇_ ).

NDC probands were from DDD and GEL trios where the proband was undiagnosed and both parents were unaffected (N=2,866 trios). Control samples were trios from the two birth cohorts (ALSPAC and MCS, N=1,434 and N=2,498, respectively) as well as trios from GEL where the proband did not have DDD-like developmental disorders or NDCs (N=872).

We verified that the PGSs in the trio model did not exhibit excessive collinearity (see <u>**Supplementary Methods**</u>).

We performed various sensitivity analyses in the following subsets (<u>**Supplementary Figure 4**</u>): NDC probands *versus* controls from GEL trios only, and NDC patients from GEL and DDD *versus* each of the three control cohorts separately (GEL, MCS or ALSPAC). We also conducted the analysis while controlling for the rare variant burden score (RVBS) in GEL trios (<u>**Extended Data Figure 10**</u>; see the section below on “Analyses of PGSs and rare protein-coding variants”).

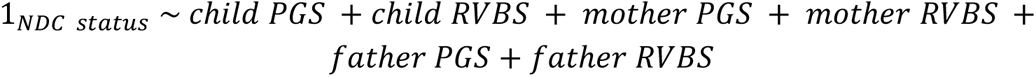

We restricted this latter analysis to GEL trios to minimize artifactual differences in rare variant calling and QC between cases and controls, which could otherwise create spurious associations.

See the <u>**Supplementary Methods**</u> for a description of how we modified the running of this trio model to investigate the hypothesis that the effects of non-transmitted alleles associated with educational attainment and cognition might be mediated by prematurity.

#### Analyses of PGSs and rare protein-coding variants

Sequence data from DDD, GEL, and MCS were annotated with the Variant Effect Predictor (VEP) ^117^. We kept the ‘worst consequence’ annotation across transcripts. From parents and probands, we extracted autosomal heterozygous protein-truncating variants (transcript ablation, frameshift, splice acceptor, splice donor and stop gained) annotated as high-confidence by LOFTEE^118^ (HC PTVs), as well as variants in the following classes which we grouped as “missense”: missense, stop lost, start lost, inframe insertion, inframe deletion, and loss-of-function variants annotated as low-confidence by LOFTEE^118^. We retained rare variants with MAF < 1 x 10^-5^ in each gnomAD super-population and MAF < 1 x 10^-4^ in the respective cohorts.

We considered four (non-mutually-exclusive) groups of damaging rare variants:

i) HC PTVs in constrained genes (pLI > 0.9^119^)
ii) HC PTVs and missense variants (MPC ≥ 2^120^) in constrained genes (pLI > 0.9)
iii) HC PTVs in monoallelic DDG2P genes with a loss-of-function mechanism (i.e. “absent gene product”)
iv) HC PTVs and missense variants (MPC ≥ 2) in monoallelic DDG2P genes with a loss-of-function mechanism.

We retained probands and parents who were heterozygous for these variants. We required the variants in the children to have been inherited from a parent.

To investigate whether parental assortment leads to correlated rare and common variant burden, we calculated rare variant burden scores (RVBSs) as the number of rare variants in the classes described above, then calculated the Pearson’s correlation coefficients between RVBSs and PGSs using the “cor” function in R. We used trios in which both parents were unaffected in this analysis. RVBSs were corrected for 20 genetic principal components using linear regression models. We then calculated the correlation coefficients between the PC-adjusted RVBSs in parents and the PC-adjusted PGSs in their partners. We also assessed the correlation within the same person amongst either children or parents. We repeated the analysis in subsets of trios where the proband was undiagnosed as well as in trios where the proband had a monogenic *de novo* diagnosis (<u>**Supplementary Figure 6**</u>). The main analysis in <u>**Figure 5**</u> and the sensitivity analysis in <u>**Extended Data Figure 10**</u> is based on group (ii) above, whereas **<u>Supplementary Figure 6</u>**, **<u>7</u>** and **<u>8</u>** show the results for all four groups of variants. To investigate whether the results were affected by uncorrected population structure, we also calculated RVBSs using rare synonymous variants in either monoallelic DDG2P genes with a loss-of-function mechanism or constrained genes, and assessed their correlation with PGS.

To assess whether PGS modify penetrance of rare inherited variants, we conducted one-sided paired *t*-tests comparing the PGS between unaffected parents transmitting a damaging variant to their affected offspring who inherited the variant (<u>**Supplementary Figure 8**</u>). We hypothesized that the unaffected parents would have a more protective polygenic background than their affected offspring (i.e. higher PGS_EA_, PGS_CP_, PGS_NonCogEA_ and lower PGS_SCZ_, PGS_NDC,DDD_). If more than one parent transmitted a variant to a proband, one parent-child pair was chosen at random from the trio. We used trios where the proband was undiagnosed and both parents were unaffected in this analysis.

### Construction and incorporation of weights for MCS

We were concerned that control cohorts might not be random samples of the population with respect to educational attainment, and that this might bias our effect sizes for the difference in PGSs between cases and controls (**<u>Supplementary Note 3</u>**). We decided to use MCS, for which extensive sociodemographic data are available, to calculate a mean PGS that would be representative of the general population, using inverse-probability weighting. MCS deliberately oversampled minority ethnic and disadvantaged individuals ^121^ (sampling bias), and they provide sampling weights to account for this. Additionally, missingness in each wave of data collection, including the collection of DNA for genotyping, was nonrandom (non-response bias). To correct for non-response bias, we produced non-response weights per individual using the inverse of the probability of being genotyped estimated from a logistic regression, considering covariates collected at the first study sweep, as previously described^121,122^ (**<u>Supplementary Methods</u>**). We fitted the model to predict who was within the sample of unrelated GBR-ancestry children with genotype data (N=5,884 of 6,036 children who had complete data for these covariates), and separately to predict who was within the subset of these that additionally had genotype data on both parents (N=2,445 of 2,498 trio children who had no missingness). To produce weights that account for both sampling bias and non-response bias, we multiplied the non-response weight from regression models by the sampling weights provided by MCS. These weights were then used to calculate adjusted PGSs shown in <u>**Extended Data Figure 4**</u> and <u>**Extended Data Figure 5C**</u> and adjusted correlation between PGS and RVBS shown in <u>**Supplementary Figure 7**</u>.

## Data availability

The raw and post-quality control genotype array data and exome sequence data from DDD are available through European Genome-phenome Archive, under EGAS00001000775.

Whole-genome sequence data and phenotypic data from the 100,000 Genomes project can be accessed by application to Genomics England (https://www.genomicsengland.co.uk/research/academic/join-gecip). GWAS summary statistics of neurodevelopmental conditions generated in this study are available in Supplementary Data. Researchers can apply to access genotype array data from ALSPAC (https://www.bristol.ac.uk/alspac/researchers/access/) and MCS (https://cls.ucl.ac.uk/data-access-training/data-access/). Publicly available GWAS summary statistics can be accessed at various resources: http://www.thessgac.org/data, https://pgc.unc.edu/for-researchers/download-results/, and https://egg-consortium.org/Gestational-duration-2023.html.

## Code availability

We used publicly available software: LDpred (https://github.com/bvilhjal/ldpred), LDSC (https://github.com/bulik/ldsc), GCTA-LDMS (https://yanglab.westlake.edu.cn/software/gcta/#GREMLinWGSorimputeddata), PCGC regression (https://dougspeed.com/pcgc-regression/), GenomicSEM (https://github.com/PerlineDemange/non-cognitive/blob/master/GenomicSEM/Genetic%20correlations/Without%20using%20SNP%20effects/function_rG_woSNP.R), and LHC-MR (https://github.com/LizaDarrous/lhcMR).

## Extended Data Figures

**Extended Data Figure 1.**
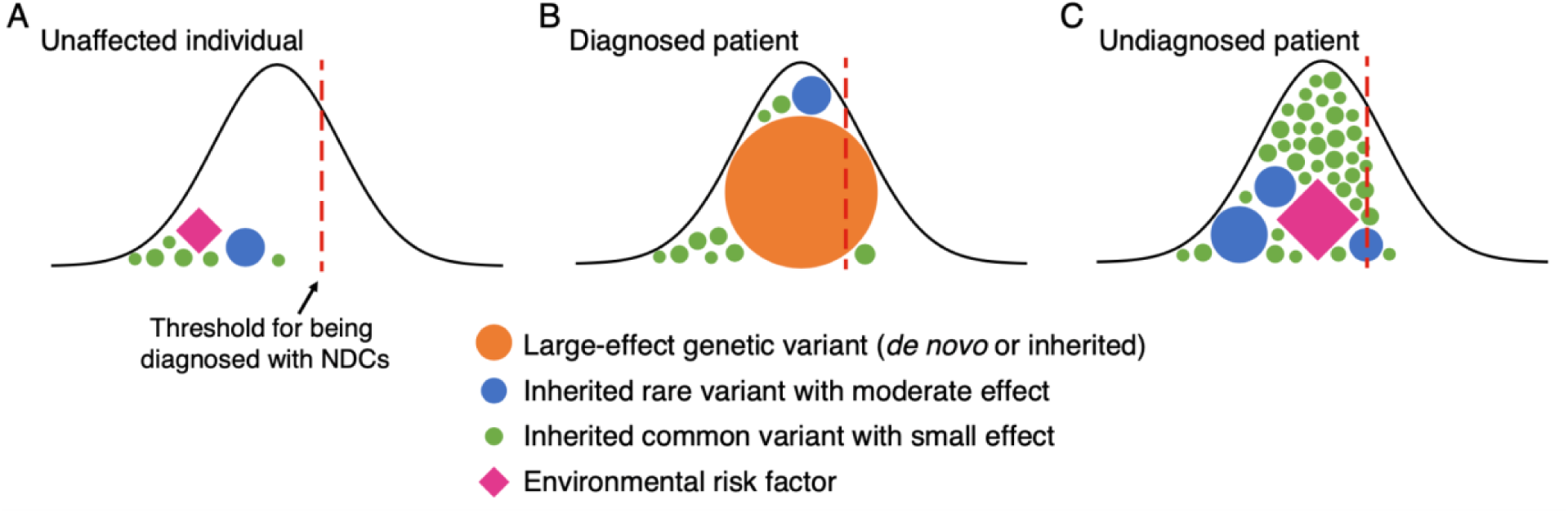
Schematic of the liability threshold model for rare neurodevelopmental disorders, illustrating why one might expect patients with a monogenic diagnosis to have less polygenic (common variant) risk than those without a monogenic diagnosis. The normal distribution represents the underlying distribution of liability in the population, which is assumed to be Gaussian. Both genetic and environmental factors of different effects contribute to this total liability. Each panel represents a hypothetical example of one individual, either unaffected (**A**), affected and diagnosed with a monogenic cause (**B**), or affected and without a monogenic diagnosis (**C**). The red line indicates a threshold for being diagnosed with neurodevelopmental conditions. Circles represent different genetic factors, and diamonds represent environmental factors. The size of circles and diamonds represents their impact on disease risk. The undiagnosed patient (C) has more green circles (i.e. risk-increasing common variants) than the patient with a monogenic diagnosis (B), in whom the orange circle (i.e. diagnostic large-effect variant) is sufficient on its own to push the patient over the diagnostic threshold.

**Extended Data Figure 2.**
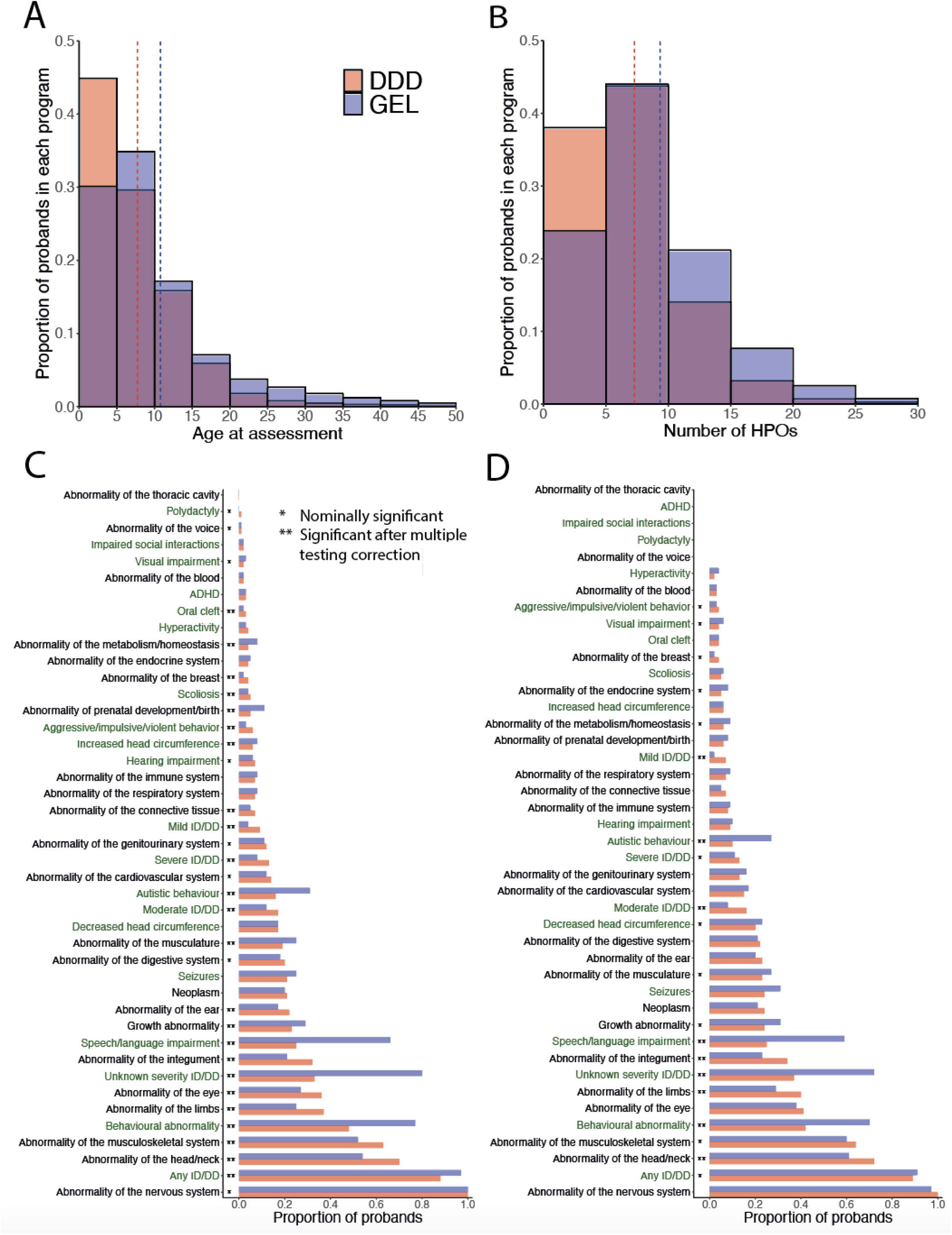
Distribution of age at assessment (**A**) and number of HPO terms (**B**) in both DDD and GEL probands with neurodevelopmental conditions who have GBR ancestry. The vertical lines indicate the means. A small number of probands in each program were aged over 50 and had more than 30 HPOs, and these have been omitted from the plot due to data sharing restrictions. **C**) Proportion of probands from each cohort with at least one HPO term within the indicated chapter (black text) or specific phenotype (green text), ordered by the prevalence in DDD. The asterisks indicate results from a logistic regression testing whether there was a significant difference in phenotype prevalence between cohorts after controlling for sex and age (** indicates p-value < 0.05/43; * indicates p-value < 0.05). **D**) Proportion of probands recruited to both DDD and GEL (N=789) with at least one HPO term within the indicated chapter (black) or specific phenotype (green text) from the phenotype information from each program, ordered by the prevalence in DDD. The same logistic regression was used as in **C**).

**Extended Data Figure 3.**
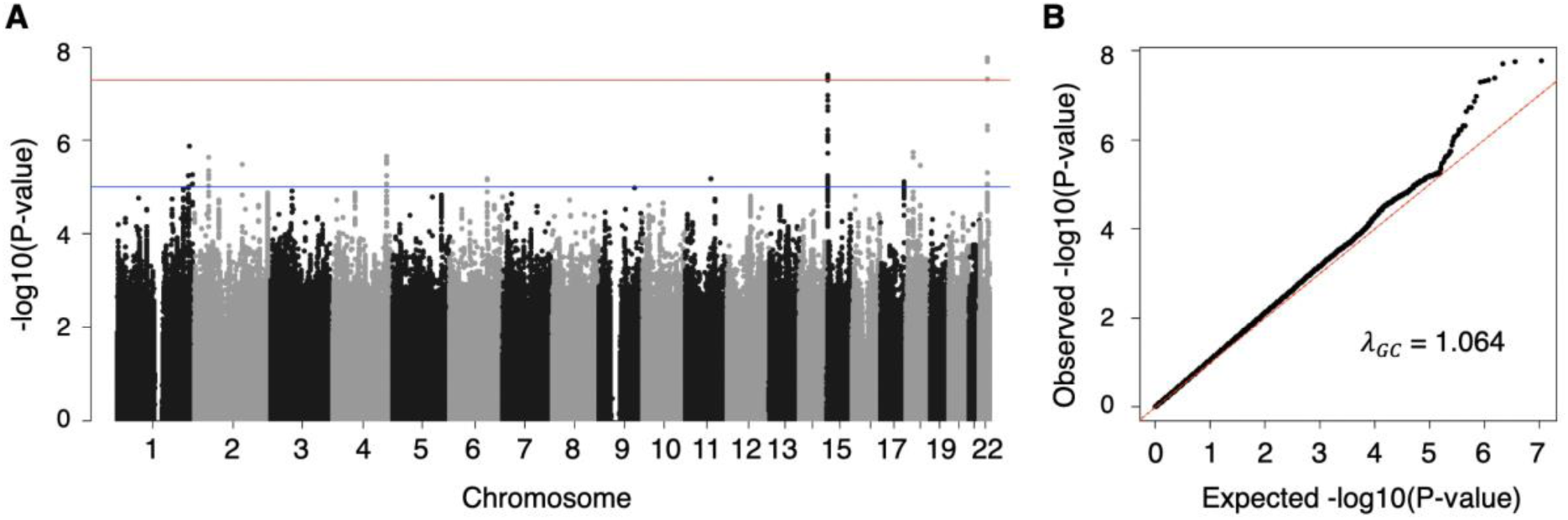
Manhattan plot (**A**) and quantile-quantile plot (**B**) of GWAS meta-analysis of neurodevelopmental conditions. We meta-analyzed the GWASs derived from DDD-UKHLS (6,397 cases with neurodevelopmental conditions and 9,270 controls from UKHLS) and GEL (3,618 cases and 13,667 controls). We used overlapping SNPs with MAF >1% in both cohorts. The red line indicates the genome-wide significance threshold (5x10^-8^).

**Extended Data Figure 4.**
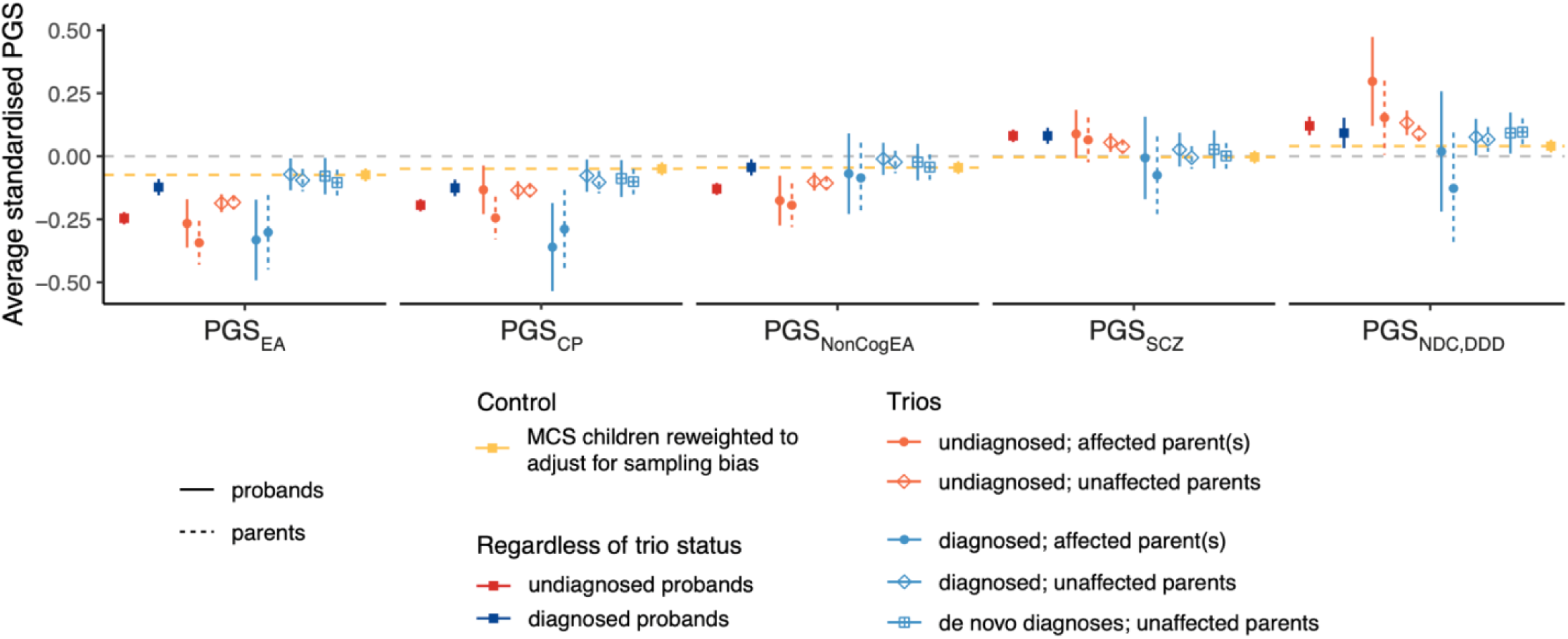
Average polygenic scores in undiagnosed (red) and diagnosed (blue) probands with neurodevelopmental conditions from DDD and GEL combined, as well as in MCS children reweighted to adjust for sampling bias and non-response bias (yellow). Subsets of probands with neurodevelopmental conditions and their parents from trios are shown in light red (undiagnosed subsets) and light blue (diagnosed subsets). The polygenic scores have been standardized such that the UKHLS+GEL controls have mean = 0 and standard deviation = 1 (except for PGS_NDC,DDD_ for which only GEL controls were used to standardise). Yellow horizontal lines indicate weighted average polygenic scores in MCS children, which should reflect an unbiased estimate for the background population. PGS_NDC,DDD_ was tested in a held-out set of patients in DDD. Error bars show 95% confidence intervals. See also <u>**Supplementary Table 5 and 7**</u> for results of statistical tests of differences between groups.

**Extended Data Figure 5.**
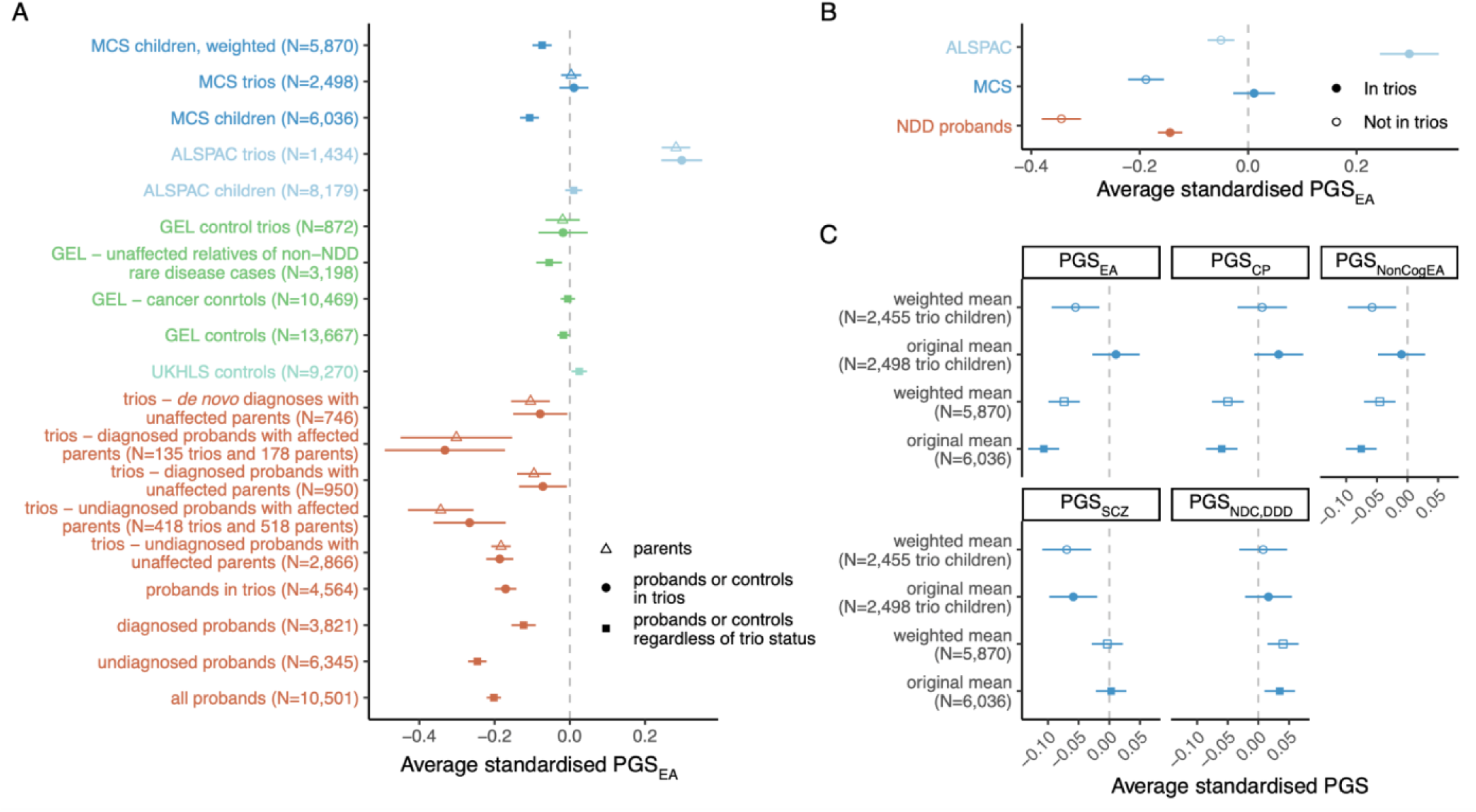
**A**) Average polygenic score for educational attainment (PGS_EA_) in different control cohorts and subsets thereof, subsets of probands with neurodevelopmental conditions, and their unaffected parents. **B**) Comparing average PGS_EA_ in trio probands and probands who did not have genetic data on both parents in ALSPAC, MCS, and affected patients from DDD and GEL. Note that in the case of DDD, “in trios” refers to those who had *exome sequence* data on both parents (only a subset of which also had genotype array data, since we prioritized genotyping full trios for which the child was undiagnosed), whereas in the rest of the manuscript (except for <u>**Figure 2B**</u> which uses the same definition as here), “trio proband” refers to those who had *genotype* data on both parents. **C**) Average polygenic scores for all five traits in MCS before and after reweighting to adjust for sampling bias and attrition. Note that the PGS are corrected for 20 PCs and then normalized so that a combined set of unrelated controls from UKHLS and GEL have mean = 0 and standard deviation = 1. Error bars show 95% confidence intervals.

**Extended Data Figure 6.**
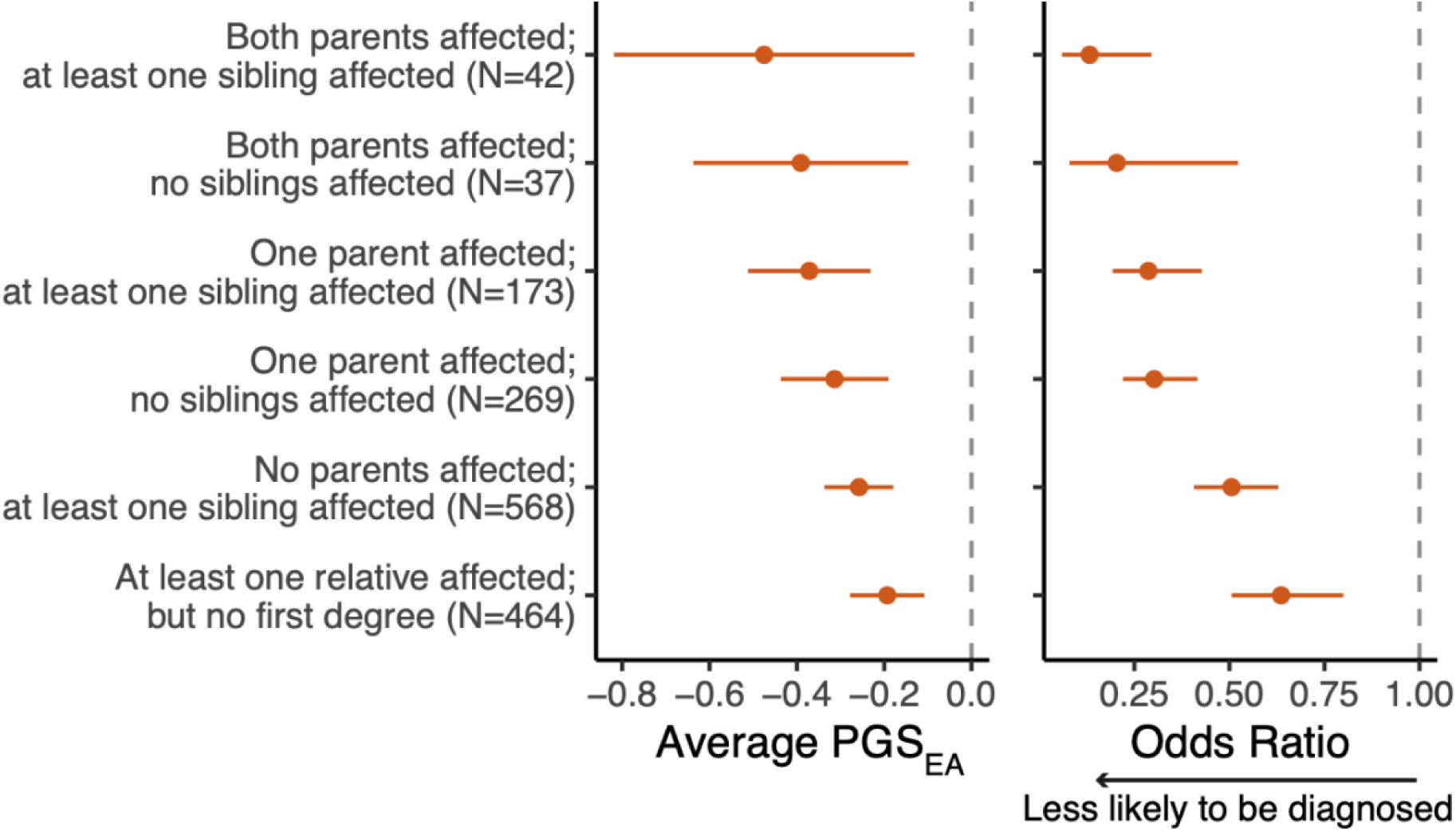
Association between different configurations of affected relatives and the child’s PGS_EA_ (left) or average diagnostic rate (right). Left: Average proband PGS_EA_ in subgroups with different configurations of affected relatives based on the number of affected parents, siblings, and more distant relatives. Right: Odds ratio for having a monogenic diagnosis, compared to probands with no affected relatives. See **<u>Supplementary Methods</u>** for a description of how this was calculated.

**Extended Data Figure 7.**
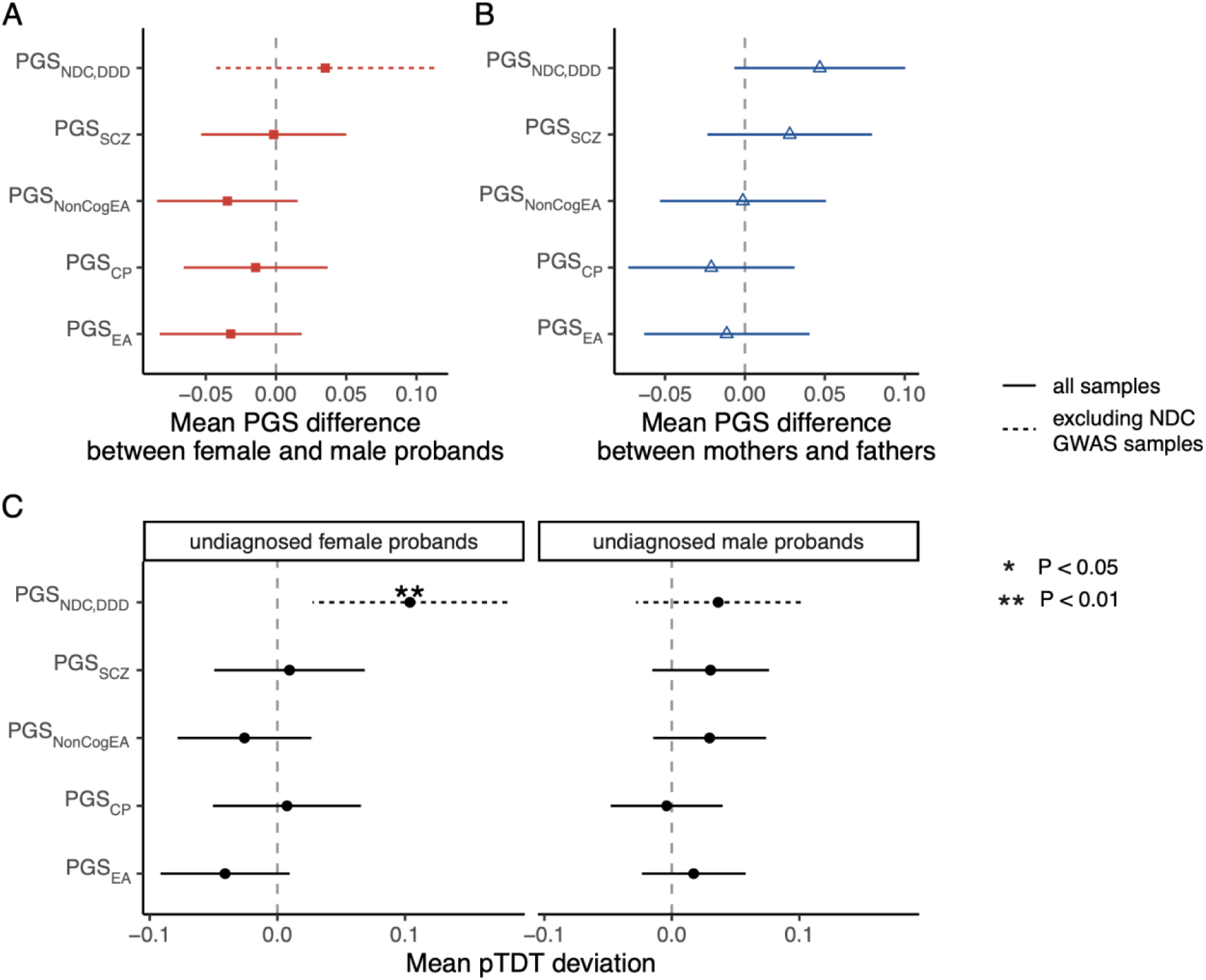
**A**) Comparison of polygenic scores between undiagnosed male and female probands in DDD and GEL combined. We used all undiagnosed probands with neurodevelopmental conditions regardless of trio status in this analysis (N=1,426 females and N=2,427 males in DDD; N=112 females and N=146 males in DDD excluding GWAS samples; N=918 females and N=1,574 males in GEL). A positive difference indicates that female probands have higher PGS than male probands. **B**) Comparison of polygenic scores between unaffected mothers and fathers of undiagnosed probands from a combined sample of 1,523 trios and 1,343 trios from DDD and GEL, respectively. A positive difference indicates that mothers have higher PGS than fathers. **C**) pTDT results in undiagnosed female and male probands with unaffected parents (N=586 females and N=937 males in DDD; N=99 females and N=125 males in DDD excluding GWAS samples; N=490 females and N=853 males in GEL). Error bars show 95% confidence intervals. The significant result that passes Bonferroni correction of five tests is highlighted by two asterisks.

**Extended Data Figure 8.**
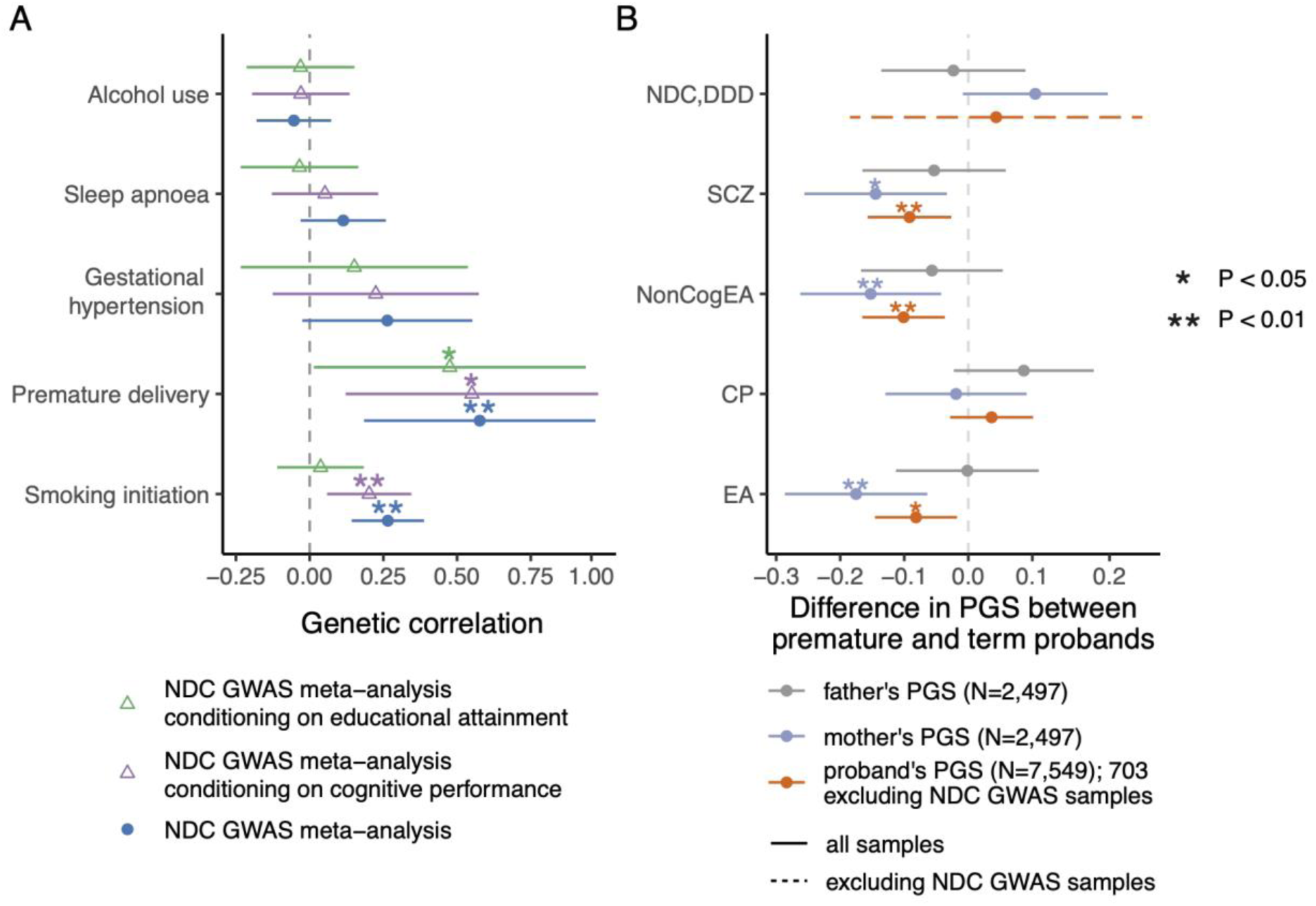
Exploring prenatal factors that may influence risk of neurodevelopmental disorders. (**A**) Genetic correlations between neurodevelopmental conditions and prenatal risk factors, before and after conditioning on educational attainment or cognitive performance. Genetic correlations with our GWAS meta-analysis for neurodevelopmental conditions was estimated using Linkage Disequilibrium Score Regression. Those conditioned on the GWAS summary statistics for educational attainment or cognitive performance were estimated using GenomicSEM. (**B**) Association between PGSs and prematurity, a risk factor for neurodevelopmental conditions, estimated in DDD. See <u>**Supplementary Table 8**</u> for sample sizes. Note that for PGS_NDC,DDD_, probands who were included in the GWAS were not tested, which left 703 probands, of which 83 were born prematurely. A negative estimate indicates that probands who were born prematurely or their parents had a lower polygenic score. Associations that pass Bonferroni correction for five traits in (A) or five polygenic scores in (B) are indicated by two asterisks and nominally significant results by one asterisk. Error bars show 95% confidence intervals.

**Extended Data Figure 9.**
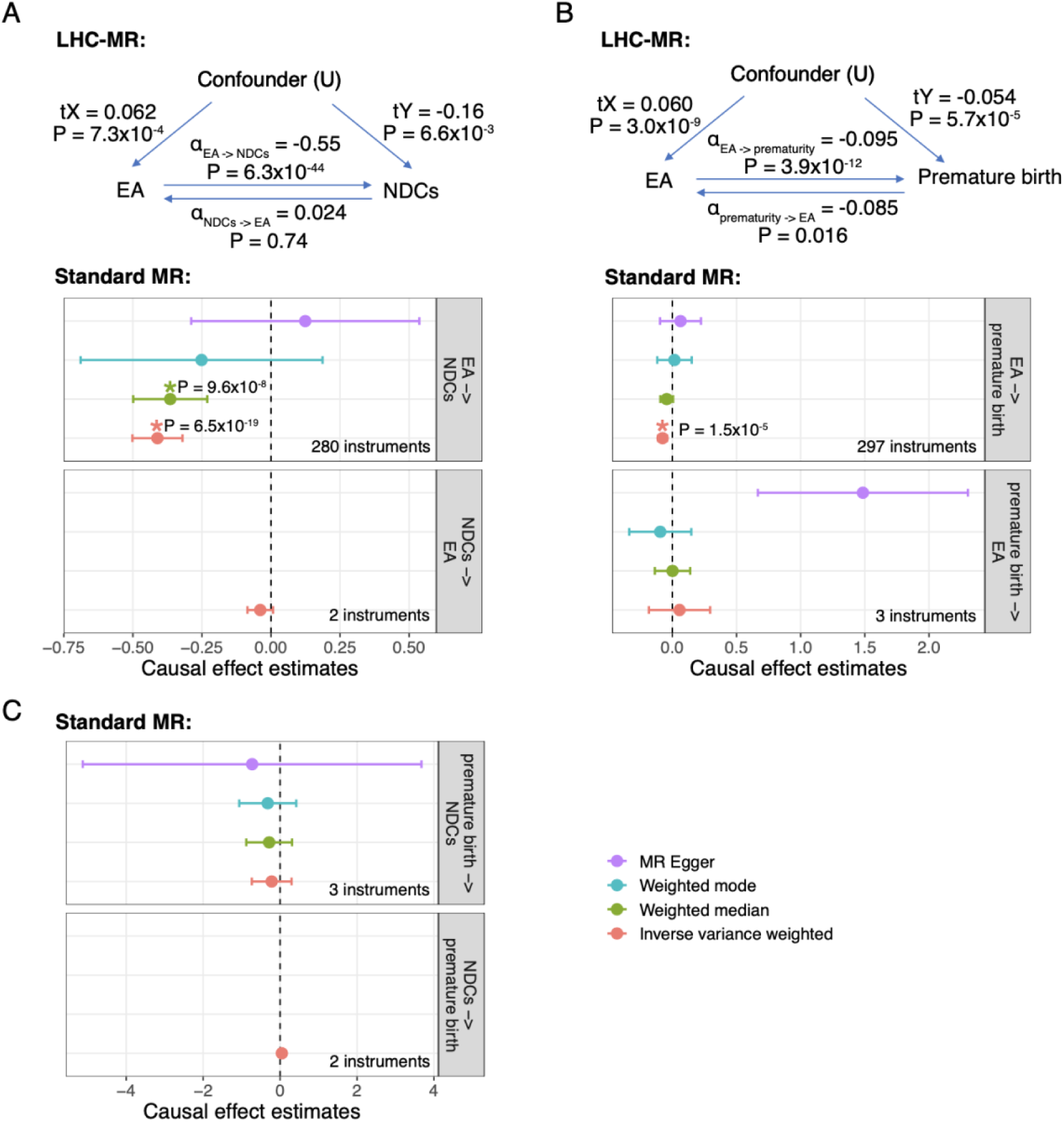
Causal effect estimates between educational attainment, neurodevelopmental conditions, and preterm birth from Mendelian randomization. The top panels show bi-directional relationships between educational attainment and giving birth prematurely, and between educational attainment and neurodevelopmental conditions, inferred by the Latent Heritable Confounder-Mendelian randomization method (LHC-MR), which uses all genome-wide SNPs. α_X->Y_ indicates the causal effect of the exposure (X) on the outcome (Y) and α_Y->X_ indicates the reverse causal effect. The causal effects of the heritable confounder on the exposure and the outcome are annotated as tX and tY, respectively. The forest plots on the bottom show the causal effects inferred using the standard Mendelian randomization methods. Up to four different methods were used, as indicated in the legend, but not all were used to test each hypothesis, depending on the number of instruments available (see **<u>Supplementary</u> <u>Methods</u>**). The dots show point estimates and the lines are 95% confidence intervals calculated using standard errors. Estimates that are significant are highlighted with an asterisk and exact p-values are annotated.

**Extended Data Figure 10.**
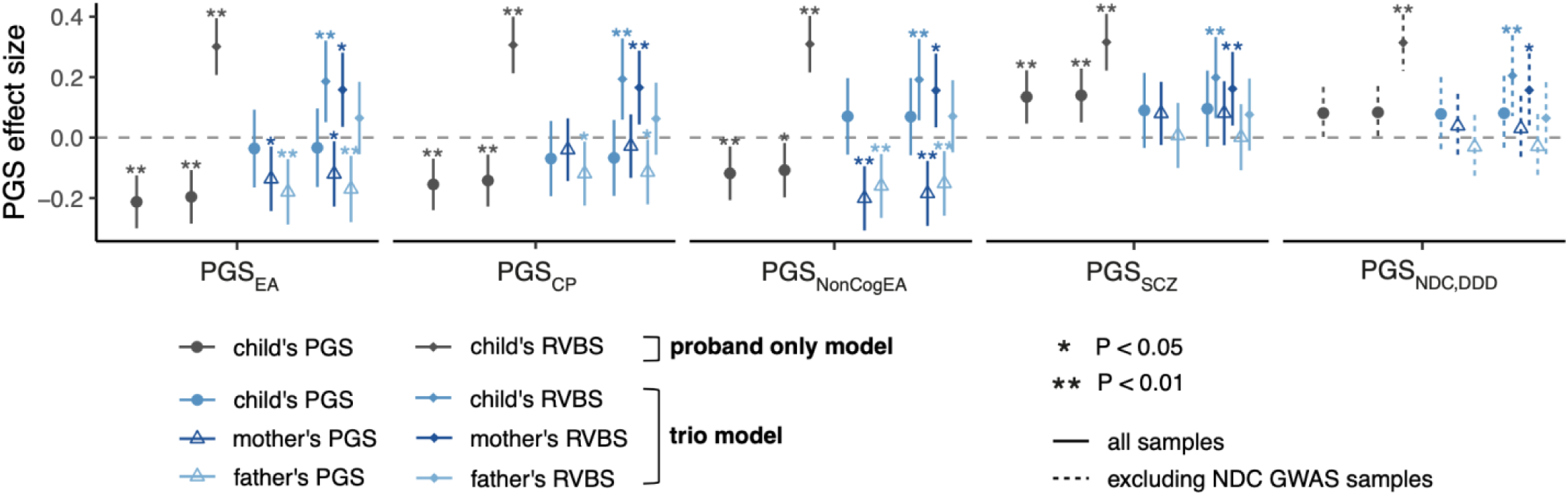
Association coefficients of polygenic scores (PGSs) and rare variant burden scores (RVBS) in the ‘proband only’ and ‘trio’ models, from logistic regressions of case/control status within GEL (N=1,343 trios in which the proband with a neurodevelopmental condition is undiagnosed and parents are unaffected and 872 trios without neurodevelopmental conditions). Case/control status was regressed on either the child’s PGS, the child’s PGS and child’s RVBS, all three trio members’ polygenic scores (trio model), or all three trio members’ polygenic scores and RVBSs (trio model+RVBS). The RVBS was defined as the number of rare damaging PTVs and missense variants in constrained genes (requiring these to be inherited in the child), corrected for genetic principle components.

## Supplementary Data

**Supplementary Data 1.** Summary statistics from the GWAS of neurodevelopmental conditions comparing cases to controls within the Genomics England (GEL) 100,000 Genomes Project.

**Supplementary Data 2.** Summary statistics from the GWAS of neurodevelopmental conditions comparing DDD cases to UKHLS controls, excluding the Scottish samples from DDD.

**Supplementary Data 3.** Summary statistics from the GWAS meta-analysis of neurodevelopmental conditions combining the DDD and GEL GWASs.

## Supplementary Tables

## Supplementary Methods

## Supplementary Figures

## Supplementary Notes

Supplementary Note 1: Phenotypic comparisons of the cohorts

Supplementary Note 2: Genome-wide significant hits from the GWAS meta-analysis of neurodevelopmental conditions

Supplementary Note 3: Potential ascertainment biases in control cohorts and their effects

Supplementary Note 4: Examining sex differences in polygenic risk

Supplementary Note 5: Exploring the role of prenatal risk factors in mediating common variant risk

Supplementary Note 6: Role of PGS in modifying the penetrance of rare variants

## Notes

### Competing Interest Statement

Matthew Hurles is non-executive director and holds stock in Congenica Inc., and is also a consultant for AstraZeneca.

### Author Declarations

The Cambridge South Research Ethics Committee gave ethical approval for this work The Republic of Ireland Research Ethics Committee gave ethical approval for this work The East of England-Cambridge Central Research Ethics Committee gave ethical approval for this work

## References

1. Nguengang Wakap, S., et al. Estimating cumulative point prevalence of rare diseases: analysis of the Orphanet database. Eur. J. Hum. Genet. 28, 165–173 (2020).

2. Sanders, S. J. et al. A framework for the investigation of rare genetic disorders in neuropsychiatry. Nat. Med. 25, 1477–1487 (2019).

3. Manickam, K. et al. Exome and genome sequencing for pediatric patients with congenital anomalies or intellectual disability: an evidence-based clinical guideline of the American College of Medical Genetics and Genomics (ACMG). Genet. Med. 23, 2029–2037 (2021).

4. Srivastava, S. et al. Correction: Meta-analysis and multidisciplinary consensus statement: exome sequencing is a first-tier clinical diagnostic test for individuals with neurodevelopmental disorders. Genet. Med. 22, 1731–1732 (2020).

5. Wright, C. F. et al. Genomic Diagnosis of Rare Pediatric Disease in the United Kingdom and Ireland. N. Engl. J. Med. 388, 1559–1571 (2023).

6. Niemi, M. E. K. et al. Common genetic variants contribute to risk of rare severe neurodevelopmental disorders. Nature 562, 268–271 (2018).

7. Kurki, M. I. et al. Contribution of rare and common variants to intellectual disability in a sub-isolate of Northern Finland. Nat. Commun. 10, 410 (2019).

8. Gardner, E. J. et al. Reduced reproductive success is associated with selective constraint on human genes. Nature 603, 858–863 (2022).

9. Chen, C.-Y. et al. The impact of rare protein coding genetic variation on adult cognitive function. Nat. Genet. 55, 927–938 (2023).

10. Kingdom, R., Beaumont, R. N., Wood, A. R., Weedon, M. N. & Wright, C. F. Genetic modifiers of rare variants in monogenic developmental disorder loci. medRxiv (2022) doi:10.1101/2022.12.15.22283523.

11. Rolland, T. et al. Phenotypic effects of genetic variants associated with autism. Nat. Med. 29, 1671–1680 (2023).

12. 12. Fenner, E., et al. Rare coding variants in schizophrenia-associated genes affect generalised cognition in the UK Biobank. *bioRxiv* (2023) doi:10.1101/2023.08.14.23294074.

13. Murray, R. M., Bhavsar, V., Tripoli, G. & Howes, O. 30 Years on: How the Neurodevelopmental Hypothesis of Schizophrenia Morphed Into the Developmental Risk Factor Model of Psychosis. Schizophr. Bull. 43, 1190–1196 (2017).

14. O’Brien, H. E. et al. Expression quantitative trait loci in the developing human brain and their enrichment in neuropsychiatric disorders. Genome Biol. 19, 194 (2018).

15. Mallard, T. T. et al. Multivariate GWAS of psychiatric disorders and their cardinal symptoms reveal two dimensions of cross-cutting genetic liabilities. Cell Genom 2, (2022).

16. Wolstencroft, J. et al. Neuropsychiatric risk in children with intellectual disability of genetic origin: IMAGINE, a UK national cohort study. Lancet Psychiatry 9, 715–724 (2022).

17. Marquis, S. M., McGrail, K. & Hayes, M. V. A population-level study of the mental health of siblings of children who have a developmental disability. SSM Popul Health 8, 100441 (2019).

18. Sullivan, P. F. et al. Family history of schizophrenia and bipolar disorder as risk factors for autism. Arch. Gen. Psychiatry 69, 1099–1103 (2012).

19. Baker, K. et al. Childhood intellectual disability and parents’ mental health: integrating social, psychological and genetic influences. Br. J. Psychiatry 218, 315–322 (2021).

20. Zarrei, M. et al. Gene copy number variation and pediatric mental health/neurodevelopment in a general population. Hum. Mol. Genet. 32, 2411–2421 (2023).

21. Alexander-Bloch, A. et al. Copy Number Variant Risk Scores Associated With Cognition, Psychopathology, and Brain Structure in Youths in the Philadelphia Neurodevelopmental Cohort. JAMA Psychiatry 79, 699–709 (2022).

22. Chawner, S. J. R. A. et al. Genotype-phenotype associations in children with copy number variants associated with high neuropsychiatric risk in the UK (IMAGINE-ID): a case-control cohort study. Lancet Psychiatry 6, 493–505 (2019).

23. Falconer, D. S. The inheritance of liability to certain diseases, estimated from the incidence among relatives. Ann. Hum. Genet. 29, 51–76 (1965).

24. The genetics of neurodevelopmental disorders: Mitchell/the genetics of neurodevelopmental disorders. (John Wiley & Sons, 2015).

25. Bergen, S. E. et al. Joint Contributions of Rare Copy Number Variants and Common SNPs to Risk for Schizophrenia. Am. J. Psychiatry 176, 29–35 (2019).

26. Antaki, D. et al. A phenotypic spectrum of autism is attributable to the combined effects of rare variants, polygenic risk and sex. Nat. Genet. 1–9 (2022).

27. Wright, C. F. et al. Making new genetic diagnoses with old data: iterative reanalysis and reporting from genome-wide data in 1,133 families with developmental disorders. Genet. Med. 20, 1216–1223 (2018).

28. Kuchenbaecker, K. B. et al. Evaluation of Polygenic Risk Scores for Breast and Ovarian Cancer Risk Prediction in BRCA1 and BRCA2 Mutation Carriers. J. Natl. Cancer Inst. 109, (2017).

29. Kong, A. et al. The nature of nurture: Effects of parental genotypes. Science 359, 424– 428 (2018).

30. Okbay, A. et al. Polygenic prediction of educational attainment within and between families from genome-wide association analyses in 3 million individuals. Nat. Genet. 54, 437–449 (2022).

31. Young, A. I. et al. Mendelian imputation of parental genotypes improves estimates of direct genetic effects. Nat. Genet. 54, 897–905 (2022).

32. Howe, L. J. et al. Within-sibship genome-wide association analyses decrease bias in estimates of direct genetic effects. Nat. Genet. 54, 581–592 (2022).

33. Demange, P. A. et al. Estimating effects of parents’ cognitive and non-cognitive skills on offspring education using polygenic scores. Nat. Commun. 13, 4801 (2022).

34. Bates, T. C. et al. Social Competence in Parents Increases Children’s Educational Attainment: Replicable Genetically-Mediated Effects of Parenting Revealed by Non-Transmitted DNA. Twin Res. Hum. Genet. 22, 1–3 (2019).

35. Wang, B. et al. Robust genetic nurture effects on education: A systematic review and meta-analysis based on 38,654 families across 8 cohorts. Am. J. Hum. Genet. 108, 1780–1791 (2021).

36. 36. 1000 Genomes Project Consortium et al. A global reference for human genetic variation. Nature 526, 68–74 (2015).

37. Lee, J. J. et al. Gene discovery and polygenic prediction from a genome-wide association study of educational attainment in 1.1 million individuals. Nat. Genet. 50, 1112–1121 (2018).

38. Trubetskoy, V. et al. Mapping genomic loci implicates genes and synaptic biology in schizophrenia. Nature 604, 502–508 (2022).

39. Davies, G. et al. Study of 300,486 individuals identifies 148 independent genetic loci influencing general cognitive function. Nat. Commun. 9, 2098 (2018).

40. Lee, J. J. et al. Gene discovery and polygenic prediction from a 1.1-million-person GWAS of educational attainment. Nat. Genet. 50, 1112 (2018).

41. Demontis, D. et al. Genome-wide analyses of ADHD identify 27 risk loci, refine the genetic architecture and implicate several cognitive domains. Nat. Genet. 55, 198–208 (2023).

42. Demange, P. A. et al. Investigating the genetic architecture of noncognitive skills using GWAS-by-subtraction. Nat. Genet. 53, 35–44 (2021).

43. Brainstorm Consortium et al. Analysis of shared heritability in common disorders of the brain. Science 360, (2018).

44. Grotzinger, A. D. et al. Genomic structural equation modelling provides insights into the multivariate genetic architecture of complex traits. Nat Hum Behav 3, 513–525 (2019).

45. Joseph, R. M. et al. Neurocognitive and Academic Outcomes at Age 10 Years of Extremely Preterm Newborns. Pediatrics 137, (2016).

46. Aarnoudse-Moens, C. S. H., Weisglas-Kuperus, N., van Goudoever, J. B. & Oosterlaan, J. Meta-analysis of neurobehavioral outcomes in very preterm and/or very low birth weight children. Pediatrics 124, 717–728 (2009).

47. Huang, J., Zhu, T., Qu, Y. & Mu, D. Prenatal, Perinatal and Neonatal Risk Factors for Intellectual Disability: A Systemic Review and Meta-Analysis. PLoS One 11, e0153655 (2016).

48. Crequit, S. et al. Association between social vulnerability profiles, prenatal care use and pregnancy outcomes. BMC Pregnancy Childbirth 23, 465 (2023).

49. Morelli, S., Nolan, B., Palomino, J. C. & Van Kerm, P. The Wealth (Disadvantage) of Single-Parent Households. Ann. Am. Acad. Pol. Soc. Sci. 702, 188–204 (2022).

50. Weiner, D. J. et al. Polygenic transmission disequilibrium confirms that common and rare variation act additively to create risk for autism spectrum disorders. Nat. Genet. 49, 978– 985 (2017).

51. 51. Nivard, M. G., et al. Neither nature nor nurture: Using extended pedigree data to elucidate the origins of indirect genetic effects on offspring educational outcomes. https://psyarxiv.com/bhpm5/download?format=pdf (2022).

52. Young, A. I. et al. Relatedness disequilibrium regression estimates heritability without environmental bias. Nat. Genet. 50, 1304–1310 (2018).

53. Young, A. S. Estimation of indirect genetic effects and heritability under assortative mating. bioRxiv 2023.07.10.548458 (2023) doi:10.1101/2023.07.10.548458.

54. Nivard, M. G. et al. Neither nature nor nurture: Using extended pedigree data to understand indirect genetic effects on offspring educational outcomes. (2022) doi:10.31234/osf.io/bhpm5.

55. Solé-Navais, P. et al. Genetic effects on the timing of parturition and links to fetal birth weight. Nat. Genet. 55, 559–567 (2023).

56. Granés, L., Torà-Rocamora, I., Palacio, M., De la Torre, L. & Llupià, A. Maternal educational level and preterm birth: Exploring inequalities in a hospital-based cohort study. PLoS One 18, e0283901 (2023).

57. Yengo, L. et al. Imprint of assortative mating on the human genome. Nat Hum Behav 2, 948–954 (2018).

58. Reynolds, C. A., Baker, L. A. & Pedersen, N. L. Multivariate models of mixed assortment: phenotypic assortment and social homogamy for education and fluid ability. Behav. Genet. 30, 455–476 (2000).

59. van Leeuwen, M., van den Berg, S. M. & Boomsma, D. I. A twin-family study of general IQ. Learn. Individ. Differ. 18, 76–88 (2008).

60. Mascie-Taylor, C. G. Spouse similarity for IQ and personality and convergence. Behav. Genet. 19, 223–227 (1989).

61. Jencks, C. Inequality: A Reassessment of the Effect of Family and Schooling in America. (Allen Lane, 1973).

62. Loehlin, J. C. Heredity-environment analyses of Jencks’s IQ correlations. Behav. Genet. 8, 415–436 (1978).

63. Horwitz, T. B., Balbona, J. V., Paulich, K. N. & Keller, M. C. Evidence of correlations between human partners based on systematic reviews and meta-analyses of 22 traits and UK Biobank analysis of 133 traits. Nat Hum Behav (2023) doi:10.1038/s41562-023-01672-z.

64. Sunde, H. F. et al. Genetic similarity between relatives provides evidence on the presence and history of assortative mating. bioRxiv 2023.06.27.546663 (2023) doi:10.1101/2023.06.27.546663.

65. Nordsletten, A. E. et al. Patterns of Nonrandom Mating Within and Across 11 Major Psychiatric Disorders. JAMA Psychiatry 73, 354–361 (2016).

66. Greve, A. N. et al. A Nationwide Cohort Study of Nonrandom Mating in Schizophrenia and Bipolar Disorder. Schizophr. Bull. 47, 1342–1350 (2021).

67. Cabrera-Mendoza, B., Wendt, F. R., Pathak, G. A., Yengo, L. & Polimanti, R. The impact of assortative mating, participation bias, and socioeconomic status on the polygenic risk of behavioral and psychiatric traits. bioRxiv (2022) doi:10.1101/2022.11.29.22282912.

68. Smolen, C. et al. Assortative mating and parental genetic relatedness drive the pathogenicity of variably expressive variants. medRxiv (2023) doi:10.1101/2023.05.18.23290169.

69. Kingdom, R. et al. Rare genetic variants in genes and loci linked to dominant monogenic developmental disorders cause milder related phenotypes in the general population. The American Journal of Human Genetics Preprint at 10.1016/j.ajhg.2022.05.011 (2022).

70. 70. Deciphering Developmental Disorders Study. Prevalence and architecture of de novo mutations in developmental disorders. Nature 542, 433–438 (2017).

71. Border, R. et al. Cross-trait assortative mating is widespread and inflates genetic correlation estimates. Science 378, 754–761 (2022).

72. Balbona, J. V., Kim, Y. & Keller, M. C. Estimation of Parental Effects Using Polygenic Scores. Behav. Genet. 51, 264–278 (2021).

73. Potharst, E. S. et al. High incidence of multi-domain disabilities in very preterm children at five years of age. J. Pediatr. 159, 79–85 (2011).

74. Cheong, J. L. Y. et al. Changing Neurodevelopment at 8 Years in Children Born Extremely Preterm Since the 1990s. Pediatrics 139, (2017).

75. Beauregard, J. L., Drews-Botsch, C., Sales, J. M., Flanders, W. D. & Kramer, M. R. Does Socioeconomic Status Modify the Association Between Preterm Birth and Children’s Early Cognitive Ability and Kindergarten Academic Achievement in the United States? Am. J. Epidemiol. 187, 1704–1713 (2018).

76. Lacalle, L., Martínez-Shaw, M. L., Marín, Y. & Sánchez-Sandoval, Y. Intelligence Quotient (IQ) in school-aged preterm infants: A systematic review. Front. Psychol. 14, 1216825 (2023).

77. Wong, H. S. & Edwards, P. Nature or nurture: a systematic review of the effect of socio-economic status on the developmental and cognitive outcomes of children born preterm. Matern. Child Health J. 17, 1689–1700 (2013).

78. Crump, C., Sundquist, J. & Sundquist, K. Preterm or early term birth and risk of attention-deficit/hyperactivity disorder: a national cohort and co-sibling study. Ann. Epidemiol. 86, 119–125.e4 (2023).

79. Husby, A., Wohlfahrt, J. & Melbye, M. Gestational age at birth and cognitive outcomes in adolescence: population based full sibling cohort study. BMJ 380, e072779 (2023).

80. Thoma, M. E., Copen, C. E. & Kirmeyer, S. E. Short Interpregnancy Intervals in 2014: Differences by Maternal Demographic Characteristics. NCHS Data Brief 1–8 (2016).

81. Kandel, D. B., Griesler, P. C. & Schaffran, C. Educational attainment and smoking among women: risk factors and consequences for offspring. Drug Alcohol Depend. 104 **Suppl 1**, S24–33 (2009).

82. Goldenberg, R. L., Culhane, J. F., Iams, J. D. & Romero, R. Epidemiology and causes of preterm birth. Lancet 371, 75–84 (2008).

83. Madley-Dowd, P. et al. Maternal smoking during pregnancy and offspring intellectual disability: sibling analysis in an intergenerational Danish cohort. Psychol. Med. 52, 1847– 1856 (2022).

84. Havdahl, A. et al. Associations Between Pregnancy-Related Predisposing Factors for Offspring Neurodevelopmental Conditions and Parental Genetic Liability to Attention-Deficit/Hyperactivity Disorder, Autism, and Schizophrenia: The Norwegian Mother, Father and Child Cohort Study (MoBa). JAMA Psychiatry 79, 799–810 (2022).

85. 85. van Alten, S., Domingue, B. W., Galama, T. & Marees, A. T. Reweighting the UK Biobank to reflect its underlying sampling population substantially reduces pervasive selection bias due to volunteering. bioRxiv (2022) doi:10.1101/2022.05.16.22275048.

86. Wright, C. F. et al. Genetic diagnosis of developmental disorders in the DDD study: a scalable analysis of genome-wide research data. Lancet 385, 1305–1314 (2015).

87. DECIPHER: Database of Chromosomal Imbalance and Phenotype in Humans Using Ensembl Resources. Am. J. Hum. Genet. 84, 524–533 (2009).

88. Köhler, S. et al. The Human Phenotype Ontology project: linking molecular biology and disease through phenotype data. Nucleic Acids Res. 42, D966–74 (2014).

89. 89. Deciphering Developmental Disorders Study. Large-scale discovery of novel genetic causes of developmental disorders. Nature 519, 223–228 (2015).

90. Turnbull, C. et al. The 100 000 Genomes Project: bringing whole genome sequencing to the NHS. BMJ 361, k1687 (2018).

91. Turro, E. et al. Whole-genome sequencing of patients with rare diseases in a national health system. Nature 583, 96–102 (2020).

92. Aggregated variant calls - genomics England trusted research environment user guide. https://re-docs.genomicsengland.co.uk/aggv2/.

93. Chang, C. C. et al. Second-generation PLINK: rising to the challenge of larger and richer datasets. GigaScience vol. 4 Preprint at 10.1186/s13742-015-0047-8 (2015).

94. Purcell, S. et al. PLINK: a tool set for whole-genome association and population-based linkage analyses. Am. J. Hum. Genet. 81, 559–575 (2007).

95. Manichaikul, A. et al. Robust relationship inference in genome-wide association studies. Bioinformatics 26, 2867–2873 (2010).

96. McFall, S., Petersen, J., Kaminska, O. & Lynn, P. Understanding Society—The UK Household Longitudinal Study: Waves 2 and 3 Nurse Health Assessment, 2010–2012 Guide to Nurse Health …. Colchester: University of Essex.

97. Boyd, A. et al. Cohort Profile: the ‘children of the 90s’--the index offspring of the Avon Longitudinal Study of Parents and Children. Int. J. Epidemiol. 42, 111–127 (2013).

98. Fraser, A. et al. Cohort Profile: the Avon Longitudinal Study of Parents and Children: ALSPAC mothers cohort. Int. J. Epidemiol. 42, 97–110 (2013).

99. Connelly, R. & Platt, L. Cohort profile: UK Millennium Cohort Study (MCS). Int. J. Epidemiol. 43, 1719–1725 (2014).

100. Joshi, H. & Fitzsimons, E. The Millennium Cohort Study: the making of a multi-purpose resource for social science and policy. Longit. Life Course Stud. 7, 409–430 (2016).

101. Northstone K, Ben Shlomo Y, Teyhan A et al. The Avon Longitudinal Study of Parents and children ALSPAC G0 Partners: A cohort profile. https://wellcomeopenresearch.org/articles/8-37/v1.

102. Fitzsimons, E. et al. Collection of genetic data at scale for a nationally representative population: the UK Millennium Cohort Study. Longit. Life Course Stud. 13, 169–187 (2021).

103. McInnes, L., Healy, J. & Melville, J. UMAP: Uniform Manifold Approximation and Projection for Dimension Reduction. arXiv [stat.ML] (2018).

104. Taliun, D. et al. Sequencing of 53,831 diverse genomes from the NHLBI TOPMed Program. Nature 590, 290–299 (2021).

105. Das, S. et al. Next-generation genotype imputation service and methods. Nat. Genet. 48, 1284–1287 (2016).

106. McCarthy, S. et al. A reference panel of 64,976 haplotypes for genotype imputation. Nat. Genet. 48, 1279–1283 (2016).

107. Willer, C. J., Li, Y. & Abecasis, G. R. METAL: fast and efficient meta-analysis of genomewide association scans. Bioinformatics 26, 2190–2191 (2010).

108. Bulik-Sullivan, B. K. et al. LD Score regression distinguishes confounding from polygenicity in genome-wide association studies. Nat. Genet. 47, 291–295 (2015).

109. Grotzinger, A. D., Fuente, J. de la, Privé, F., Nivard, M. G. & Tucker-Drob, E. M. Pervasive Downward Bias in Estimates of Liability-Scale Heritability in Genome-wide Association Study Meta-analysis: A Simple Solution. Biol. Psychiatry 93, 29–36 (2023).

110. Yang, J. et al. Genetic variance estimation with imputed variants finds negligible missing heritability for human height and body mass index. Nat. Genet. 47, 1114–1120 (2015).

111. Golan, D., Lander, E. S. & Rosset, S. Measuring missing heritability: inferring the contribution of common variants. Proc. Natl. Acad. Sci. U. S. A. 111, E5272–81 (2014).

112. Vilhjálmsson, B. J. et al. Modeling Linkage Disequilibrium Increases Accuracy of Polygenic Risk Scores. Am. J. Hum. Genet. 97, 576–592 (2015).

113. 113. International HapMap 3 Consortium et al. Integrating common and rare genetic variation in diverse human populations. Nature 467, 52–58 (2010).

114. Bycroft, C. et al. The UK Biobank resource with deep phenotyping and genomic data. Nature 562, 203–209 (2018).

115. Lee, S. H., Goddard, M. E., Wray, N. R. & Visscher, P. M. A better coefficient of determination for genetic profile analysis. Genet. Epidemiol. 36, 214–224 (2012).

116. Lee, S. H., Wray, N. R., Goddard, M. E. & Visscher, P. M. Estimating missing heritability for disease from genome-wide association studies. Am. J. Hum. Genet. 88, 294–305 (2011).

117. McLaren, W. et al. The Ensembl Variant Effect Predictor. Genome Biol. 17, 1–14 (2016).

118. Karczewski, K. J. et al. The mutational constraint spectrum quantified from variation in141,456 humans. Nature 581, 434–443 (2020).

119. Lek, M. et al. Analysis of protein-coding genetic variation in 60,706 humans. Nature 536, 285–291 (2016).

120. 120. Samocha, K. E., et al. Regional missense constraint improves variant deleteriousness prediction. bioRxiv 148353 (2017) doi:10.1101/148353.

121. 121. Plewis, I. The Millennium Cohort Study: Technical Report on Sampling (4th Edition). http://doc.ukdataservice.ac.uk/doc/4683/mrdoc/pdf/mcs_technical_report_on_sampling_4t h_edition.pdf (2007).

122. Plewis, I. Non-Response in a Birth Cohort Study: The Case of the Millennium Cohort Study. Int. J. Soc. Res. Methodol. 10, 325–334 (2007).

