## Supplementary material for "Dissecting the contribution of common variants to risk of rare neurodevelopmental conditions": FAQ and lay summaries

### Lay summary of and frequently asked questions about the paper “Dissecting the contribution of polygenic background to risk and penetrance of rare neurodevelopmental conditions”

*This document was written primarily by Emilie Wigdor, Patrick Campbell and Hilary Martin, with input from other authors of the paper (particularly Elizabeth Radford and Helen Firth) as well as participant representatives from the 100,000 Genomes project (Jillian Hastings-Ward, Hannah Podd and Hannah Humphrey) and the patient organization Unique.*

**Note to readers: Part 1 provides a brief summary of the results in simple terms. It is available both in an “Easy Read” version and a slightly more technical version. Part 2 (Frequently Asked Questions) provides more detailed information about the paper in lay language, and is aimed at science journalists or people with a similar level of understanding about this topic.**

If you are a parent or relative of a child with a neurodevelopmental condition and want to know what this study means for your child and your family, please see question 15.

#### Part 1: Lay summary

##### EasyRead version of the summary

Our DNA acts as an instruction book for how to build our bodies. We all have similar DNA but there are also differences in the DNA between us. These are like different spellings in the instruction book that make the recipes for our body unique. Some of these changes in the DNA are rare and found only in a small number of people’s DNA. Others are common and found in many people’s DNA.

Rare changes in our DNA can change how our brains develop, and cause rare brain conditions in children. These rare brain conditions often cause learning difficulties. Projects like the 100,000 Genomes Project try to find these rare DNA changes and tell families about them. Common DNA changes can also affect the chance of having these conditions and less is known about these.

We are a group of scientists and doctors interested in DNA changes in people with rare brain conditions. In a recent study, we looked at data from thousands of people with rare brain conditions and their parents. We used information from two projects in the UK: the Deciphering Developmental Disorders study and the 100,000 Genomes Project. We also studied data from thousands of people without rare brain conditions. We wanted to understand how common DNA changes contribute to these conditions.

We found that certain common DNA changes are more common in people with rare brain conditions. These same common DNA changes are similar to those that increase the chance of mental health issues like depression. (See here if you are worried about what this means for you or your child). We also found that these same DNA changes are more common in people who have spent less time in education and get lower scores on IQ tests.

**Overall, common DNA changes have only a small effect on the chance of having a rare brain condition.** But they might help explain why some people have rare brain conditions, especially if they don't have a rare DNA change. We found that these common DNA changes in parents might affect how their child's brain develops, even if the child doesn't directly inherit them. But there might be other reasons for our findings. More research is needed to understand them.

Overall, we found that common DNA changes play a small role in rare brain conditions. So, doctors probably won't use them to help patients understand the cause of their rare brain conditions any time soon. This study helps us understand how DNA and the environment work together to cause rare brain conditions. In the future, this might help families and doctors better understand, diagnose, and treat these conditions.

#### Lay summary in slightly more technical language

Rare neurodevelopmental conditions affect the growth and development of the brain in childhood. They often lead to learning difficulties and/or seizures. They are often caused by a single rare genetic change. Studies such as the 100,000 Genomes Project and the Deciphering Developmental Disorders study try to identify these rare genetic changes causing patients' conditions ("genetic diagnoses") and report them back to families. Many families find it helpful if they understand the reason why their child has additional challenges and it can also be important for their healthcare. So, it is important to do research to better understand the many different factors that contribute to neurodevelopmental conditions. Genetic changes that are **common** in the general population are known to have small effects on the chance of developing a neurodevelopmental condition. Currently, genetic diagnosis or clinical care would not be based on these common genetic changes.

In this study, we (a group of scientists and doctors) analyzed data from more than 11,500 people with a neurodevelopmental condition and 9,100 of their parents. The data were collected by two projects on rare conditions in the UK: the Deciphering Developmental Disorders (DDD) study and the 100,000 Genomes Project. We also looked at data from 26,800 people without neurodevelopmental conditions. Our goal was to better understand how common genetic changes contribute to these conditions.

We found that the common genetic changes that contribute to these rare, early-onset neurodevelopmental conditions overlap with those that increase the chance of developing later-onset mental health conditions (e.g. ADHD). (See here if you are worried about what this means for you or your child). They also overlap with the common genetic changes that are more likely to be found in people with fewer years in formal education and who get lower scores in tests of mental processes including memory and problem-solving abilities ("cognitive performance"). However, importantly, common genetic changes had only a small impact overall on the chance of someone developing a neurodevelopmental condition.

We found that common genetic changes can help to explain why some people have a neurodevelopmental condition - especially if they have not got a rare genetic diagnosis. We also learned more about how common genetic changes that affect the number of years people spend in education affect the chance of having a neurodevelopmental condition. In particular, our findings suggest that these common genetic changes, when present in the parents, may affect their child's neurodevelopmental condition, even if the child does not inherit those genetic changes directly. However, alternative technical explanations may instead be driving the findings. More work is needed to understand them.

There are limitations to this work. We showed that the role of common genetic changes in neurodevelopmental conditions is small. Because of this, they are unlikely to be used by doctors to diagnose or help patients in the near future. Nevertheless, this study brings us closer to fully understanding how different genetic and environmental factors may work together to cause neurodevelopmental conditions. In the longer term, these findings may help families and doctors to better understand, diagnose and manage these conditions.

### Part 2: Some questions and answers about the paper

#### Section 1: Introduction to neurodevelopmental conditions and their causes

By "neurodevelopmental conditions", we mean conditions that are first noticed during childhood and that affect the growth and development of the brain. They often cause intellectual disability and delays in achieving developmental milestones. Question 20 gives more information about neurodevelopmental conditions.

##### 1. What most often causes rare neurodevelopmental conditions?

Rare neurodevelopmental conditions are most often due to genetic changes. In a large percentage of people with neurodevelopmental conditions (probably at least 40%), their condition is due to a DNA change (variant) that happened in the child with the condition and is not present in the parents. These are called *de novo* variants. If they occur in one of several hundred genes, they can cause a neurodevelopmental condition. However, neurodevelopmental conditions can also be caused by rare inherited variants in a gene (by "rare", we mean variants typically seen in <1% of people (one in a hundred)). More information on how this can occur can be found here. If we can find the single rare variant (or sometimes, pair of variants) causing an individual's neurodevelopmental condition, this is referred to as a "genetic diagnosis".

##### 2. Does everyone who has a rare variant have a rare neurodevelopmental condition?

No. We all have millions of rare variants in our DNA, and most of these don't cause neurodevelopmental conditions or other conditions. These rare variants are part of the reason we are different from one another (e.g. in height and hair colour). However, it is sometimes possible to have a rare genetic change of the type that usually causes neurodevelopmental conditions but to not actually have one of these conditions. When this happens, we describe the genetic variants as having *incomplete penetrance*. This can happen because the impact of these specific rare genetic changes is influenced by other genetic changes in our DNA, as well as by our environment and random chance.

#### 3. What role does the environment play in neurodevelopmental conditions?

There are known environmental exposures that cause or contribute to the likelihood of developing a neurodevelopmental condition, without causing genetic changes. These typically occur during development when the child is still in the womb, or shortly after birth. One of the most common environmental influences that can cause neurodevelopmental conditions is being born prematurely. Unfortunately, we often do not have a good understanding of the exact causes of premature birth, and usually nothing can be done to prevent it. Exposure to certain medications or substances (e.g. certain drugs, alcohol) and viruses (e.g. Rubella virus) whilst in the womb can also cause neurodevelopmental conditions. Again, often nothing can be done to avoid these exposures.

Exposure to each of these environmental factors does not always cause neurodevelopmental conditions. **In the case of a child with a neurodevelopmental condition, it is important to have an assessment with a specialist medical professional, including a thorough medical history and examination, before any link between an individual neurodevelopmental condition and an environmental factor is made.**

See here if you have a child with a neurodevelopmental condition and are concerned about the possible role of environmental factors such as prematurity, medications, smoking or alcohol.

#### 4. What is meant by the "genetic architecture" of neurodevelopmental conditions?

This paper is broadly about an aspect of what is called the "genetic architecture" of neurodevelopmental conditions. When we talk about "genetic architecture", we mean understanding how many variants influence the chance of developing a condition, how big their individual effects are, how common they are in the population, and where in the DNA they lie (e.g. in genes or outside genes).

The picture below illustrates a model for some of the different ways it is thought neurodevelopmental conditions may arise. This picture focuses on genetic causes, since this section is about genetic architecture, but as noted in question 3, environmental causes play a role in some neurodevelopmental conditions.

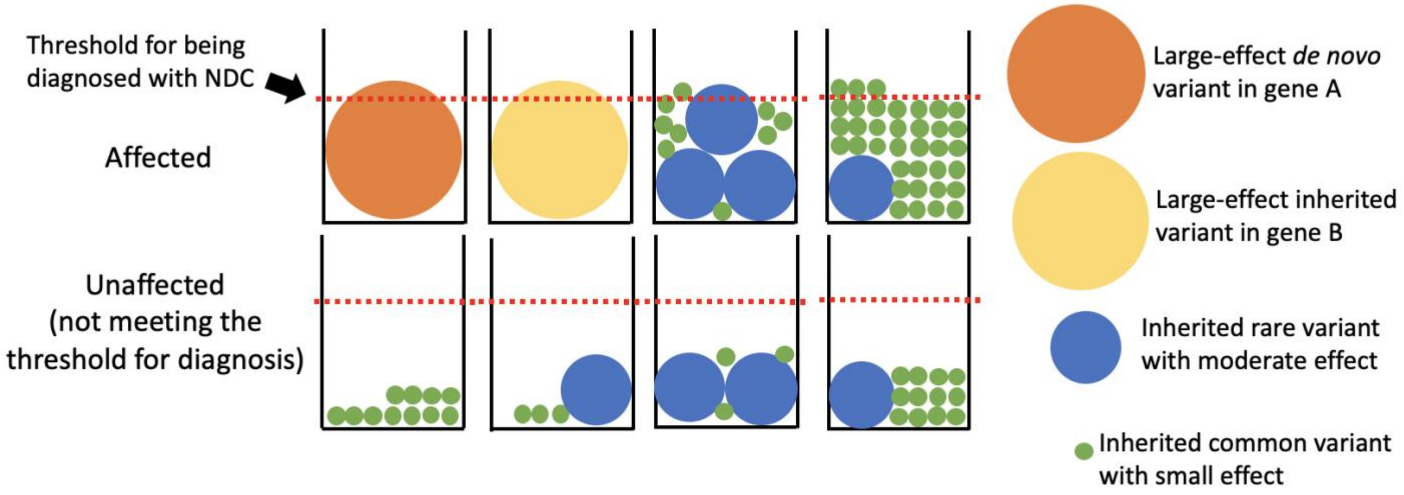


Figure 1

*In this picture, each jar represents a hypothetical example of one person. Each person carries one or more genetic variants (represented by the coloured circles) which contribute to their chance of developing a neurodevelopmental condition. The size of the circles represents the amount by which each variant influences this chance. The people in whom the circles cross the red line develop a neurodevelopmental condition; this line represents something called the "liability threshold" (see* <u>*question 23*</u> *if you want more information on this). The big orange and yellow circles represent de novo and rare inherited variants that have large enough effects on their own to cause a neurodevelopmental condition (as mentioned in* <u>*question 1*</u> *above). The middle-sized blue circles represent incompletely penetrant rare variants, which have moderate effects on the chance of developing a neurodevelopmental condition (as mentioned in* <u>*question 2*</u>*). The small green circles represent inherited common variants (seen in e.g. >1% of people in the population) which have individually very small effects on the chance of developing a neurodevelopmental condition, but which may collectively have an important effect if an individual has enough of them (e.g. in the fourth affected individual). The unaffected individuals in the bottom row have some inherited rare variants and/or some inherited common variants which contribute to the chance of developing a neurodevelopmental condition; however, they don't have enough of them to cross the red line, so they haven't developed a neurodevelopmental condition.*

To describe "genetic architecture" another way, we want to understand what percentage of affected individuals have neurodevelopmental conditions due to an orange circle, a yellow circle, or some combination of blue and green circles. Some genetic variants are not sufficient to cause a neurodevelopmental condition on their own (the blue and green circles) and are also found in unaffected individuals. Because of this, when describing genetic architecture, we often talk about the percentage of *variation* in chance of developing a condition due to different genetic mechanisms (e.g. all common variants), rather than simply the percentage of people whose neurodevelopmental condition is "explained" by a variant in a single gene. To fully understand genetic architecture, we also need to understand how different types of genetic variants act together to influence the chance of developing a given condition or to influence variability in clinical features (e.g. severity of seizures).

##### 5. Why is it important to study the genetic architecture of neurodevelopmental conditions? Shouldn't we just focus on finding genetic diagnoses?

Finding genetic diagnoses for patients (i.e. large-effect genetic variants that fully or largely explain their condition - see question 1) is really important. However, understanding genetic architecture more generally is also important for several reasons. Firstly, estimating the percentage of individuals who are likely to have a particular type of genetic diagnosis (including in genes we haven't yet found) can tell us where we should invest research efforts to maximise the number of diagnoses we can identify. It might also allow us to give better advice to parents about their chance of having another affected child, even if their first child doesn't get a genetic diagnosis. Secondly, it may be that not all individuals with neurodevelopmental conditions have a "genetic diagnosis", in the sense that they don't have a single large-effect variant causing their condition - it may be that their condition has more complex causes, that may be entirely or partially genetic. Understanding the causes of these individuals' conditions is still important. Research on this may ultimately help inform parents about their chance of having another affected child, about how their child's condition is likely to change as they age, and tell us about possible prevention/management strategies.

#### Section 2: Background to the study

##### 6. Who conducted this study? What was their overarching goal?

We are a group of geneticists and clinician researchers working in several different research centres, primarily at the Wellcome Sanger Institute. Our goal was to better understand the genetic architecture of neurodevelopmental conditions (see questions 4 and 5). Specifically, we wanted to investigate how much of a role common variants play in different groups of people with neurodevelopmental conditions, and how they influence the chance of developing a neurodevelopmental condition.

##### 7. What do we already know about the contribution of common variants to rare neurodevelopmental conditions?

A previous study found that common variants make up a small part of the overall chance of developing neurodevelopmental conditions (it estimated about 7%). It also showed that the same common genetic variants that affect the chance of developing a rare neurodevelopmental condition are also more common in people who have fewer years of formal education, in people with lower scores on cognitive tests, and in people with schizophrenia. However, beyond this, nothing was known about how exactly these common variants contribute to the chance of developing a neurodevelopmental condition, and whether they contribute to different extents in different groups of people with neurodevelopmental conditions.

#### 8. What data were used in this study and how/why were they collected?

The study was conducted in over 11,500 individuals with a rare neurodevelopmental condition from two UK-based projects that focused on finding genetic diagnoses for individuals with rare disorders: the Deciphering Developmental Disorders study and the Genomics England 100,000 Genomes Project. The study also looked at over 9,100 parents of patients with neurodevelopmental conditions, and 26,800 unrelated individuals without neurodevelopmental conditions.

The Deciphering Developmental Disorders study was set up to understand the genetic causes of developmental conditions. The study brought together doctors in twenty-four regional genetics services throughout the UK and Republic of Ireland. Similarly, the 100,000 Genomes Project was an initiative to sequence the DNA of tens of thousands of people with rare diseases and cancer (together with their family members) in order to try to find the genetic causes of their conditions.

### Section 3: Study design and results

#### 9. Let's start with the big picture - how important are common variants in neurodevelopmental conditions anyway?

This study confirmed the findings from the previous one, namely that **common genetic variation only accounts for a small proportion of the variation in chance of developing a neurodevelopmental condition**. Specifically, we estimated that this is about 10%. Despite the small overall contribution of common variants, we wanted to unpick this further and understand the nature of their contribution, and examine whether they contribute more or less in different groups of individuals with neurodevelopmental conditions. To do this, we used polygenic scores (see question 10).

#### 10. What are polygenic scores and how were they used in this study?

Background: Many human conditions and traits are affected by common genetic variants, including neurodevelopmental conditions, mental health conditions such as schizophrenia, and the number of years someone spends in formal education ("years of education"). Typically, individual common variants have, at most, a tiny impact on any given human trait and condition. However, the millions of common variants that are present in our DNA (sometimes called "polygenic background") can add up to have a small or moderate impact on our predisposition to a trait or condition.

We can add up the predicted impact of all common variants an individual has into something called a "polygenic score" to predict the likelihood of them having a particular condition (e.g. a neurodevelopmental condition) or trait (e.g. going to university). These polygenic scores are better predictors than any single common variant. **However, although polygenic scores are useful research tools for prediction at the population level, they are not**

**good predictors of whether an individual person is likely to e.g. develop a neurodevelopmental condition or go to university.**

In this study, for each individual, we calculated polygenic scores for neurodevelopmental conditions based on the degree of correlation between common variants and the chance of developing these conditions. We also calculated polygenic scores for some other traits and conditions that are relevant to neurodevelopmental conditions (i.e. there is an overlap in common genetic variants that contribute to the conditions/traits). These include years of education (see question 24 for more information on why we included this) and cognitive performance (i.e. performance on cognitive tests), as well as schizophrenia. We used these polygenic scores as proxies for individuals' genetic predisposition for conditions/propensity towards traits. We compared the polygenic scores between different groups of people with and without neurodevelopmental conditions.

We emphasize that **these polygenic scores cannot be used to accurately predict any individual's propensity to a trait or condition, although they are quite useful predictors at a population level.** For example, the polygenic score for neurodevelopmental conditions only explains ~0.1% of the variation in chance of developing neurodevelopmental conditions, and the polygenic score for years of education only explains ~13% of variation in the number of years someone spends in education and ~0.5% of the variation in chance of developing neurodevelopmental conditions.

#### 11. What did you learn about the contribution of common variants in people with neurodevelopmental conditions who have a genetic diagnosis *versus* those who don't?

We found that people who had a rare variant identified as causing their neurodevelopmental condition ('diagnosed') had, on average, a higher polygenic score for years of education and cognitive performance than those who did not ('undiagnosed'). This means that their common genetic variants predispose them to spending longer in education and scoring better on cognitive tests than individuals with a neurodevelopmental condition who don't have a genetic diagnosis. (Having said that, many people with neurodevelopmental conditions will not attend regular schools and cognitive testing may not be appropriate.) This is probably because in diagnosed individuals, a rare genetic variant has already given them a high chance of developing a neurodevelopmental condition and therefore they are expected to require a lesser contribution from common variants. (In other words, it is in keeping with the "liability threshold model" explained below in question 23, and shown in Figure 1 above.)

We found that individuals with a genetic diagnosis whose parents were unaffected did not have significantly different polygenic scores from unrelated individuals without neurodevelopmental conditions ("controls") - in other words, in individuals with a very large-effect rare variant whose parents are unaffected, common variants are not contributing to their chance of developing a neurodevelopmental condition. However, those with a genetic diagnosis for whom one or both parents were affected had similar polygenic scores to affected individuals without a genetic diagnosis; thus, they have lower polygenic scores for years of education and cognitive performance than people without neurodevelopmental conditions (meaning that their common genetic variants predispose them to fewer years in education and lower performance on cognitive tests). Thus, we learnt that common genetic variants contribute to the chance of neurodevelopmental conditions in patients without a genetic diagnosis and in patients with a genetic diagnosis who have a clinically affected parent. Figure 2 below shows some hypothetical examples of families to illustrate these trends.

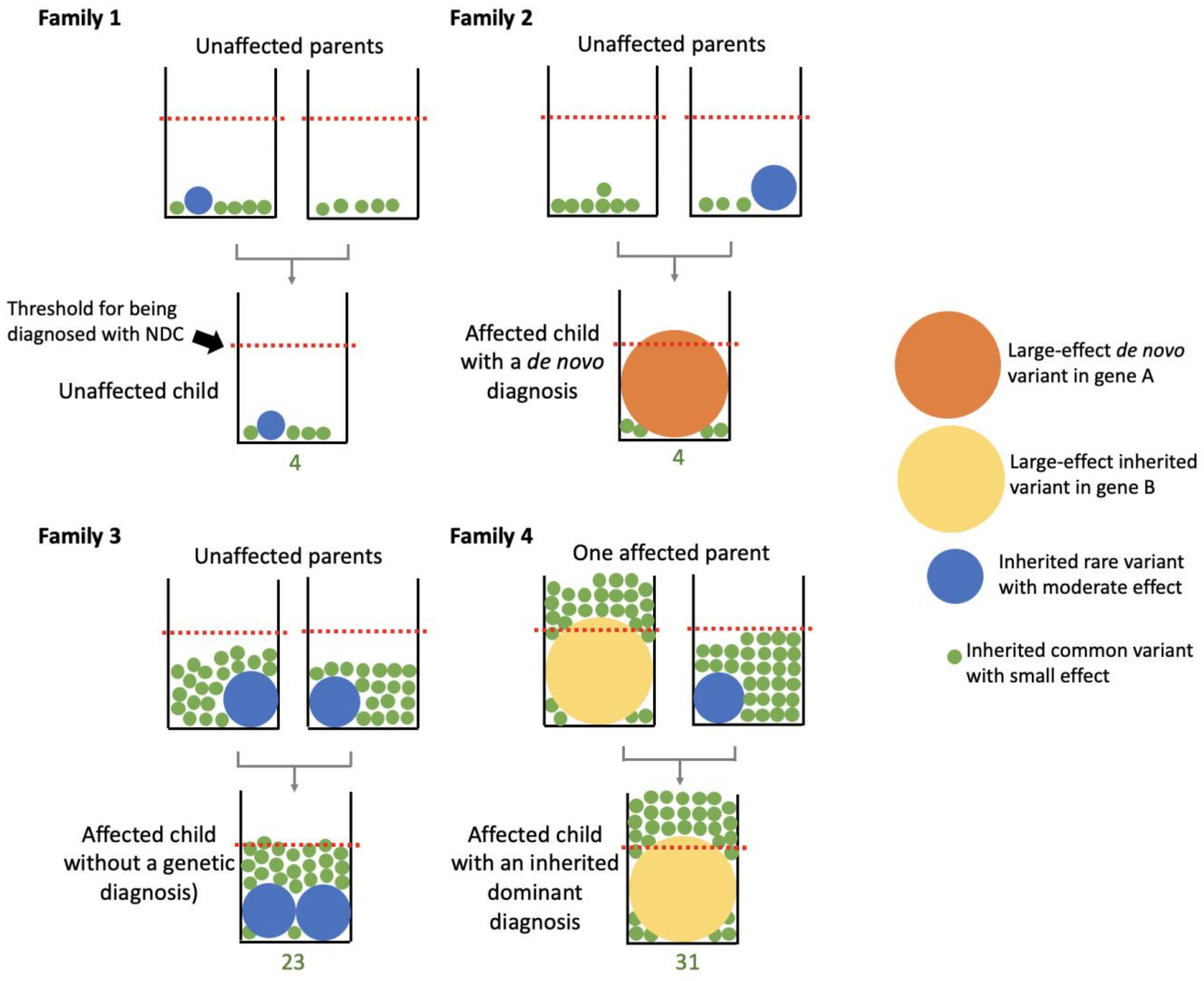


Figure 2

*In this picture, we show three hypothetical families with a child affected by a neurodevelopmental condition (families 2-4) and one hypothetical family with an unaffected child (family 1). The purpose of this figure is to exemplify the average differences in polygenic scores predisposing to neurodevelopmental conditions that we found between individuals with different characteristics (undiagnosed patients versus controls, patients diagnosed with a de novo mutation versus inherited dominant diagnosis). The number underneath each child indicates the count of green dots, which represents the degree of polygenic predisposition from common variants. In family 2, both parents are unaffected and their child has a de novo diagnosis. The polygenic score is similar to the unaffected child in family 1. In family 3, the child does not have a genetic diagnosis and has higher polygenic predisposition from common variants in addition to inherited rare variants of moderate effect. Together, these raise the child's predisposition above the threshold for developing an NDC. In family 4, one of the parents is affected as well, and the child has an inherited dominant diagnosis of a large-effect rare variant. Such children have on average the highest polygenic predisposition among all patients.*

#### 12. What did you learn from comparing the polygenic scores of people with neurodevelopmental conditions, their unaffected parents, and controls?

Most parents of people with neurodevelopmental conditions that we studied are clinically unaffected. To our surprise, we found that, on average, unaffected parents of undiagnosed children with a neurodevelopmental condition have lower polygenic scores for years of education and cognitive performance than unrelated individuals without neurodevelopmental conditions ("controls"), similar to their affected children (i.e. their common genetic variants predispose them to spending fewer years in education and performing less well on cognitive tests). This result suggests that common genetic variants in the parents may contribute to their children's chance of developing a neurodevelopmental condition, given they have a higher predisposition than controls. However, these common variants are presumably not sufficient to cause the neurodevelopmental condition observed in their child, since the parents remain unaffected.

There could be several possible explanations for this observation. To explore one of these, we wanted to test whether the common genetic variants in the parents impacted their children's chance of developing neurodevelopmental conditions *over and above* the direct effects of the variants passed on to the children. To do this, we tested whether the parents' polygenic scores for neurodevelopmental conditions and related traits were correlated with whether or not their child had a neurodevelopmental condition, after adjusting for their child's polygenic scores. We found that for several traits related to neurodevelopmental conditions (including cognitive performance and years of education), the child's polygenic score was no longer correlated with having a neurodevelopmental condition after taking into account the parents' polygenic scores. This suggests that the common variants associated with cognitive performance and years of education *do not directly* affect a child's predisposition to neurodevelopmental conditions. However, the parents' polygenic scores were associated with whether or not their child had a neurodevelopmental condition even after taking into account the child's polygenic score. This implies that common variants that are present in the parents but not passed onto the child ("non-transmitted variants") *may* be influencing the chance of the child having a neurodevelopmental condition - see question 13 for consideration of how this could happen. However, there is another possible explanation which we also discuss below in question 13.

For the polygenic score for neurodevelopmental conditions, we saw that there was still a significant effect of the child's polygenic score on whether or not they had a neurodevelopmental condition after adjusting for the parents' polygenic scores. This implies that the common genetic variants making up this polygenic score *do* have a direct effect on predisposition to neurodevelopmental conditions in the child.

#### 13. Why might the non-transmitted common variants associated with cognitive performance and years of education in parents be correlated with their children's chance of having a rare neurodevelopmental condition?

This finding (described in question 12 above) could be due to two potential influences: *indirect genetic effects* and *parental similarity*.

*Indirect genetic effects* refer to the phenomenon in which an individual's predisposition for a trait or condition is influenced by aspects of the prenatal or familial environment that are related to their parents' or relatives' genetics. We hypothesized that one way this might happen in this context is if the common variants associated with neurodevelopmental conditions influence the mother's chance of having a premature baby. Premature birth increases the chance of the child having a neurodevelopmental condition, and we know that the mother's genetics affect her chance of having a premature birth. However, **we didn't find any evidence for indirect genetic effects acting through increased likelihood of premature birth.** In theory, other mechanisms could drive indirect genetic effects, such as deprivation, which may be correlated with the parents' polygenic scores for years of education. (People with fewer years of education tend to experience more deprivation.) However, we were unable to test this hypothesis with the data we had in this study.

*Parental similarity* (referred to in the paper as "parental assortment") refers to the phenomenon in which people tend to have children with partners who have similar traits to themselves. For example, people are known to choose partners who are similar to themselves in height and in level of education. If genetics contribute to the traits for which partners are similar, partners are more *genetically* similar to each other than expected by chance. This has many implications for genetic studies. One of these is that common genetic variants captured in polygenic scores for years in education are correlated with rare genetic variants that affect years in education and related traits (including the chance of neurodevelopmental conditions) (see Figure 3 for an explanation of how this happens). Some of these rare genetic variants may be inherited by the children. This means **we are unable to confidently determine whether the correlation between non-transmitted common variants in the parents and their child's chance of neurodevelopmental conditions is driven by indirect genetic effects or whether it is due to parental similarity. It may turn out to be a mixture of both**, and hopefully future studies will shed light on this.

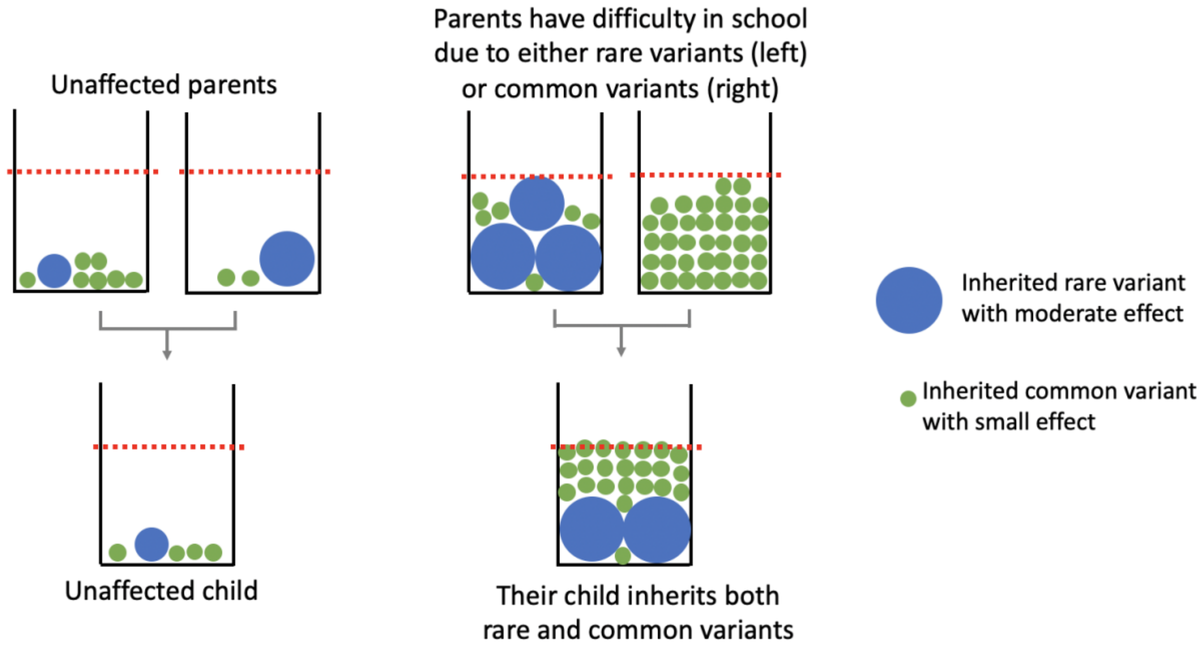


Figure 3

*Two hypothetical families in which the mother in each pair has a similar number of years in formal education to the father. Left: both parents have a low chance of developing neurodevelopmental conditions, as does their child. Right: both parents have difficulty in school due to either rare variants or common variants which contribute to their learning difficulties (reducing cognitive performance). Their child inherits some of the rare and common variants from parents. The child's risk together is then above the diagnostic threshold for a neurodevelopmental condition indicated by the red line. Imagine this with multiple couples across the population - it results in people who have more rare variants that contribute to learning difficulties also having more common variants contributing to learning difficulties.*

#### 14. What are the limitations of your study?

Like all scientific studies, there are several limitations to this work:

- As noted above, the polygenic scores we used are only very weak predictors of neurodevelopmental conditions - each of them explains <1% of the variation in chance of developing a neurodevelopmental condition. This limits our ability to detect differences in the polygenic scores between different groups. Furthermore, the predictive power of polygenic scores depends partly on the number of samples in the original genetic study from which they were derived (i.e. genome-wide association studies, described below). The sizes of the underlying genetic studies differ between traits, ranging from several hundred thousand to over one million people. This makes comparisons between the relative effects of different polygenic scores difficult.
- We have brought together a range of rare neurodevelopmental conditions in this analysis (for example, individuals with varying levels of intellectual disability, individuals with seizures versus without seizures), which may have limited our ability

to detect effects. It may be that the contribution of common genetic variants differs between different neurodevelopmental conditions.

- Parental similarity for years in education and/or cognitive ability could be influencing our results. As described in question 13, parental similarity could cause the correlation between non-transmitted common variants in the parents and whether or not their children have a neurodevelopmental condition. More generally, the correlation we observed between common and rare variants predisposing to neurodevelopmental conditions means that the findings in the paper about common genetic variants may be partly driven by the rare genetic variants which are correlated with them, which are the ones actually predisposing to neurodevelopmental conditions.

### Section 4: Implications of the study

#### 15. My child has a neurodevelopmental condition. What do the findings of this study mean for my child and our family?

**The findings from this study are not informative about any individual family or person with a neurodevelopmental condition.** This study describes a series of statistical trends relating common genetic variation to neurodevelopmental conditions at the population level. However, although statistically significant, the relationships we found between common genetic variants and neurodevelopmental conditions are weak, meaning we cannot make accurate statements about any particular individual and their chance of a neurodevelopmental condition based on their common variants, nor can we decide on treatment or clinical management based on their common variants. It is possible that in the future, after more research is done, doctors might start to use common variants to help uncover the cause/s of an individual's neurodevelopmental condition, to inform parents about how their child's condition is likely to change as they age, to tell us about possible prevention/management strategies, or to give parents advice about their chance of having another affected child. However, currently, this is not possible.

#### 16. Can you now predict who will have a neurodevelopmental condition based on their polygenic scores?

No, we cannot predict who will have a neurodevelopmental condition based on their polygenic scores. First, as explained in question 1, many neurodevelopmental conditions are caused by a single genetic variant that is not commonly found in the population, and indeed may only be seen in a single individual. Additionally, as explained in question 2, some neurodevelopmental conditions are caused by environmental factors (e.g. fetal alcohol syndrome, or Zika virus). Polygenic scores can only explain a trait to the extent that the trait is influenced by common genetic variants. Our estimates suggest that about 10% of the predisposition to neurodevelopmental conditions is due to common genetic variants, and polygenic scores currently only capture a small part of this (see questions 9 and 10 above). Thus, we will never be able to predict neurodevelopmental conditions very well using common genetic variants alone. **It is unlikely that common genetic variants alone are the sole cause of the condition for any of the individuals included in this study.**

#### 17. If the association between polygenic scores and predisposition for neurodevelopmental conditions is so small at the population level, why is this interesting?

There are many reasons why this is still interesting despite the small associations between polygenic scores and neurodevelopmental conditions:

i) While the contributions of common genetic variants may be small, it does not mean they are unimportant. All predisposing factors must be considered if we are to fully understand the causes of neurodevelopmental conditions. Despite huge advances in finding genetic causes for rare conditions, the majority of individuals with neurodevelopmental conditions remain undiagnosed. There is still a lot we do not know about potential causes of these undiagnosed conditions, and contributions from polygenic scores composed of common genetic variants may be a factor.

ii) These associations are small in part because we do not have enough data to accurately estimate the effects of common genetic variants on neurodevelopmental conditions and related traits. As the number of individuals included in genome-wide association studies increases, we will be able to produce more predictive polygenic scores. However, as noted in question 16, since common variants in total explain a small proportion (currently estimated about 10%) of the variation in chance of developing a neurodevelopmental condition, polygenic scores will never be very accurate predictors of who will develop a neurodevelopmental condition.

#### 18. How could the results of this study be used to help patients with rare neurodevelopmental conditions and their families?

Clinicians and families often find that understanding the reasons an individual has a neurodevelopmental condition is beneficial, both psychologically and medically. These results bring us closer to understanding all the factors that contribute to neurodevelopmental conditions, particularly in those who are currently undiagnosed by modern genetic techniques. However, the findings from this study are unlikely to be incorporated in clinical settings in the near future. In the longer term, as our understanding of the role of polygenic score in neurodevelopmental conditions improves, these findings might help provide more families with an explanation for their child’s neurodevelopmental condition and become part of precision testing strategies which look to incorporate the full picture of a person’s common and rare genetic variants and environmental influences on their neurodevelopmental condition.

#### 19. How might the results of this study or the information in this document be misinterpreted and why are these interpretations incorrect?

**Misinterpretation 1:** People develop neurodevelopmental conditions because their parents spent less time in education and don’t provide a good enough environment for them.

This is incorrect. Neurodevelopmental conditions are most often caused by a rare genetic variant, and such variants are passed on randomly from parents to children, or arise for the first time in the child (*de novo*). Occasionally neurodevelopmental conditions are caused by viruses, exposure to drugs in pregnancy or by premature birth. When identifying the cause of an individual child's neurodevelopmental condition, it is important to have a full medical assessment before any conclusions on the cause of that child's condition are made.

In this paper, we found evidence that common genetic variants that predispose to fewer years of education may also affect the chance of an individual having a neurodevelopmental condition. However, two points are important to bear in mind. Firstly, this only accounts for a very small proportion of the overall predisposition for these conditions. Secondly, parents have no control of which common genetic variants they carry or pass on to their children. Although we found that children's chances of having neurodevelopmental conditions is correlated with common genetic variants in their parents that are correlated with fewer years of education and that are not transmitted to the children, the explanation for this is unclear, as described in question 13.

**Misinterpretation 2:** No patient with a neurodevelopmental condition has a single cause for their condition i.e. a mutation in a single gene is insufficient to cause the condition - it will only do so in the context of a particular genetic background.

This is incorrect. As noted in question 1, many people with neurodevelopmental conditions have a single genetic cause for their condition, regardless of their genetic background. In this study, we mainly investigate neurodevelopmental conditions that cannot be explained by a single genetic cause.

**Misinterpretation 3:** Because my child has a neurodevelopmental condition, they or my other children are likely to develop ADHD or schizophrenia.

In this study, we found that common variants that are correlated with increased chance of neurodevelopmental conditions are also correlated with increased chance of ADHD and schizophrenia. **However, this is a result at the population level, and it does not have any meaningful implications for individuals about their or their families' chance of developing these conditions.** This is because common variants that are associated with ADHD and schizophrenia explain less than one quarter of an individual's chance of these conditions. Any increased chance from more common variants predisposing to ADHD or schizophrenia are unlikely to be meaningful for an individual as there are many other factors that make up a larger proportion of chance of these conditions.

**Misinterpretation 4:** My child developed a condition because they were born prematurely.

Premature birth, particularly before the age of 32 weeks, is known to be associated with increased chance of neurodevelopmental conditions. It may be in these cases that the complications associated with the premature birth themselves cause the condition. However, not all children born prematurely develop neurodevelopmental conditions and other factors may play a role.

Usually very premature birth is a random event that cannot be prevented and most often the cause of it is not identified. There is some evidence that certain genetic conditions may cause babies to be born prematurely, so in some children (we don't know what percentage), it may not be the premature birth itself causing the condition, but rather a genetic change may have caused both the premature birth and the condition.

The chance of premature birth causing neurodevelopmental conditions reduces with each additional week of pregnancy. Therefore, it is more likely that children born moderately premature (e.g. after 32 weeks) may have other environmental factors and/or genetic factors that interact to cause the condition.

**Misinterpretation 5:** My child developed a condition because of medications I took in pregnancy.

This is unlikely to be the case. Most medications are not known to cause neurodevelopmental conditions. A small number of medications, such as anti-epileptic drugs, are known to increase the chance of certain conditions in children, but as a consequence, doctors avoid prescribing them to pregnant women unless they are essential to the mother's health.

Even in cases where a child is exposed to a medication known to cause a neurodevelopmental condition, not all such children will develop a neurodevelopmental condition. It is important to have a thorough medical history, examination and assessment with a trained specialist before any link between an individual neurodevelopmental condition and a medication is made.

**Misinterpretation 6:** My child developed a condition because I smoked very occasionally in pregnancy/had a few glasses of wine.

This is unlikely to be the case. Heavy drinking in pregnancy can cause a condition called 'fetal alcohol syndrome' which involves neurodevelopmental problems. However, even if a mother does drink heavily while pregnant, her child will not always be born with this condition. It's unlikely that a small amount of alcohol or smoking would be a major factor contributing to your child's condition.

**It is important to have a thorough medical history, examination and assessment with a trained specialist before any link between an individual neurodevelopmental condition and use of alcohol/tobacco during pregnancy is made.**

The Royal College of Obstetricians and Gynaecologists has further information on alcohol and smoking for those who are currently pregnant .

#### Core concepts and terminology used above

##### 20. What are rare neurodevelopmental conditions?

Rare neurodevelopmental conditions are a group of conditions that are first noticed during childhood and affect the growth and development of the brain. They can cause children to have intellectual disability, seizures and a small head size, and delays in achieving developmental milestones, such as walking and talking. Children with these conditions are likely to need educational, health and social support throughout their lives. The individual conditions that make up this group are often very rare and may only affect a handful of individuals in the UK or even the world but cumulatively affect ~1% of newborns.

**The patient organisation UNIQUE exists to provide support and information to those with rare chromosomal and genetic conditions that cause developmental delay and/or an intellectual disability. More information on many individual rare neurodevelopmental conditions can be found on their website: https://rarechromo.org/ or by emailing.**

##### 21. What is a genetic "variant"?

Our DNA is essentially a long sequence of letters, and genetic variants are changes in those letters. All of us have millions of genetic variants in our DNA, and these are partly what makes us different from one another. These changes can be as small as one letter being different or missing, or can be larger changes or rearrangements of stretches of DNA, a little like a paragraph being removed, repeated or rearranged. Some genetic variants are commonly seen in the general population - "common variants" - while some are rarely seen in the general population - "rare variants". A variant that is seen in more than 1% of people is usually considered to be common.

##### 22. What are Genome Wide Association Studies?

In a genome-wide association study, researchers look at whether people with a particular trait or condition tend to have certain common genetic changes. These studies focus on single-letter changes in the DNA, known as single-nucleotide polymorphisms, which are commonly seen in the population. Genome-wide association studies have been successful in identifying common genetic changes associated with a range of different conditions and traits, and they are very commonly used in genetic research.

##### 23. What is the liability threshold model?

The liability threshold model is a theory to help understand a person's chance of developing a condition, such as a neurodevelopmental condition. In this model, an individual's genetics (both rare and common) and environmental experiences influence their brain development and add up to give their overall chance of developing a neurodevelopmental condition. If there are enough factors that affect the development of their brain, they will develop a condition. The theory has been explored in a range of different medical conditions, including schizophrenia and rare heart disease.

#### 24. Why did you look at years of education as a trait?

Individuals who have neurodevelopmental conditions often have intellectual disability. Thus, it seemed plausible that some of the common variants that affect the chance of neurodevelopmental conditions may overlap with those that affect cognitive ability in the general population. Indeed, a previous study found that common variants that affect scores on cognitive tests in the general population also affect the chance of an individual developing a rare neurodevelopmental condition, as do the common variants that affect the number of years someone spends in education. In genetic and social science research, the number of years someone spends in education is often used as an imperfect proxy for cognitive ability, because it is correlated with it and it is much easier to collect data on years of education (by simply asking individuals how many years they spent in education) than to administer time-consuming cognitive tests. This means that we have much larger sample sizes available for genome-wide association studies of years of education (currently around 3 million people) than cognitive ability (around 300, 000 people), which better enables us (gives us much more statistical power) to estimate the effect of genetic variants on this trait. We made use of these genetic studies to allow us to calculate polygenic scores for years of education, which we used in our study as predictors (albeit not very good ones) of people's genetic propensity towards education (as a proxy for their genetic propensity for cognitive ability).

#### 25. What do you mean by "ancestry" in the paper, why is it important in genetic studies, and why did you focus on people with British ancestry?

"Ancestry" is a construct based on shared DNA inherited from one's ancestors. It is not the same as race or ethnicity, which are social constructs used to categorize people based on a shared history, appearance, geography, culture, language or other factors. Ancestry is not a clear-cut category. A group of individuals can be divided into numerous "ancestry groups" based on genetic similarities. In this paper, we refer to "genetic ancestry" based on genetic similarity between the individuals under study and a set of reference individuals who are known to originate from particular geographic regions.

Often, to understand a trait or condition, we look for how common particular genetic variants are between two groups, for example, between patients with neurodevelopmental conditions and individuals without neurodevelopmental conditions from the general population ("controls"). The goal is to find variants that are more commonly seen among people with the trait/condition and that are likely part of the reason that person has the trait/condition. If the two groups we are comparing have genetic differences *by chance* due to having different genetic ancestries, this will give us misleading results. Specifically, if the trait/condition is influenced by environmental factors that are correlated with ancestry, we will see many genetic changes that are also correlated with the trait/condition, but they do not causally affect it.

To prevent our results being confused in this way by genetic ancestries, standard practice in genetic studies (including genome-wide association studies, results from which are used to construct polygenic scores) has been to only analyze the largest genetically homogenous subset of the data, which in practice means only including individuals from the ancestry group most heavily represented in the sample. Most individuals in the Deciphering Developmental Disorders study and the 100,000 Genomes Project have genetic ancestries similar to those whose grandparents originate from the UK. Thus, we restricted our analyses to this subset of individuals, whom we identified based on their genetic similarity to people from an external reference dataset who were known to have British ancestries.

It is well recognised in genetic research that there is an underrepresentation of people from Non-European ancestries. There are a number of large scale efforts to try to correct this. Moreover, many efforts are currently underway to develop statistical methods to analyze multi-ancestry cohorts and individuals whose genetic ancestry is a mix of different populations. However, at present, for this study, we unfortunately do not have enough samples to perform a meaningful statistical analysis of the role of common variation in neurodevelopmental conditions for individuals other than those with British ancestries. We hope this will be possible in the future and highlight it as future work.
