## Supplementary Information for "Dissecting the contribution of common variants to risk of rare neurodevelopmental conditions"

### Supplementary Methods

#### Quality control of genetic data

##### DDD

The DDD cohort was genotyped on three genotyping arrays: the Illumina HumanCoreExome chip (CoreExome), the Illumina OmniChipExpress chip (OmniChip), and the Illumina Infinium Global Screening Array (GSA). Some probands were genotyped on more than one chip, as shown in **Supplementary Figure 9**.

###### CoreExome and OmniChip

Quality control (QC) of CoreExome (including DDD patients and 9,270 UKHLS controls genotyped on the same chip) and OmniChip data were performed by Niemi et al. in each dataset separately[1]. Briefly, samples with sex discrepancies, high missingness (≥3%) in variants with

MAF ≥10%, or high or low heterozygosity (+/- 3 SDs from the mean) were removed. Individuals who had genetically inferred white European ancestry, defined using reference samples from the 1,000 Genomes project, were kept. One individual from each pair of related individuals (identical by descent ≥12% using PLINK) were removed from amongst the CoreExome samples, and those who were related to trios genotyped on Omnichip were also removed. Trios with > 2,000 Mendelian errors were removed. We removed variants with minor allele frequency (MAF) < 0.5%, missingness ≥3%, or a Hardy-Weinberg equilibrium (HWE) test p-value < $1x10^{-5}$. Variants with matched alleles between DDD CoreExome and UKHLS were kept so that imputation could be conducted from a common set of variants for both cohorts.

##### Global Screening Array

Global Screening Array (GSA) samples from DDD were genotyped in two batches. We removed samples that were discordant with exome sequencing data previously generated on all DDD individuals[2]. Sample swaps, duplicates, those with sex discrepancies, and missingness ≥ 5% were removed. We examined heterozygosity rate and removed outliers with a rate < 0.158 or > 0.170. Palindromic, duplicated and multiallelic variants were removed, as well as indels. Variants with either a call rate < 95% or with significant deviation from HWE ($p < 1\times10^{-6}$) were also removed. The two batches were then merged to overlapping SNPs. Variants with a significantly different genotype rate ($p < 1\times10^{-50}$) and allele frequency between the two batches were removed. Trios with > 200 Mendelian errors were removed, as were SNPs with Mendelian errors in >1% of trios. Variants were again filtered to remove those with a significant deviation from HWE ($p < 1\times10^{-6}$) and subset to those with a MAF > 1%. The number of samples remaining before and after these QC steps are given in **Supplementary Tables 10 and 11**.

To identify GSA individuals of genetically inferred European ancestry, we first projected the post-QC samples (N=9,572) onto 1,000 Genomes phase 3 individuals[3] using the smartpca function from EIGENSOFT version 7.2.1[4]. We used linkage disequilibrium (LD)-pruned SNPs (pairwise $r^2$ <0.2 in batches of 50 SNPs with sliding windows of 5) with MAF > 5% and removed 24 regions with high or long-range LD, including the HLA[5], leaving 90,563 variants. We identified a subset of samples that projected onto European ancestry samples from 1,000 Genomes (PC2 > 0.0175), leaving 9,534 European ancestry DDD individuals (**Supplementary Figure 10**). We then performed another principal component analysis (PCA) in the unrelated individuals within the loosely-defined European ancestry subset, projecting related individuals onto them using smartpca[4]. We applied Uniform Manifold Approximation and Projection (UMAP)[6] using the first ten PCs and identified a homogeneous subgroup of 8,489 individuals (**Supplementary Figure 11**). Since we intended to merge trio data genotyped on GSA and Omnichip array chips in downstream analysis, we conducted a PCA to further confirm that the GSA individuals were well matched for ancestry with OmniChip individuals previously identified to have GBR ancestry[1] (**Supplementary Figure 12**).

##### GEL

Variant calling and initial QC were performed by Genomics England. We used 78,195 post-QC germline genomes from the Aggregated Variant Calls (aggV2) prepared by the GEL team. All samples were sequenced with 150bp paired-end reads using Illumina HiSeqX and processed on the Illumina North Star Version 4 Whole Genome Sequencing Workflow, comprising the iSAAC Aligner and Starling Small Variant Caller. Aggregation of single-sample gVCFs were performed using the Illumina software gVCF genotyper. We retained 78,195 samples that passed various QC filters including sample contamination <3%, ratio of SNV heterozygous to homozygous calls <3, total number of SNVs between 3.2 to 4.7 million, array concordance > 90%, and median fragment size >250bp, excess of chimeric reads <5%, percentage of mapped reads >90%, and percentage at dropout < 10%. We kept variants that passed the QC filters shown in **Supplementary Table 12**.

For GWAS and PGS analyses, we applied additional QC following Kousathanas *et al.*[7]. Low-quality genotypes were masked using the bcftools setGT module performed by the GEL team. Specifically, genotypes with depth < 10 or GQ < 20, as well as heterozygous genotypes failing an allele balance binomial test with P-value < $1x10^{-3}$ were set to missing. The masked VCF files were converted to PLINK pgen format using PLINK v2.0[8]. We removed samples with mean autosomal coverage <25x. Within GEL samples who had genetically inferred European ancestry (inferred as described below), we further removed samples that had four median absolute deviations (MADs) above or below the median for the following metrics: ratio of insertions to deletions, ratio of transitions to transversions, total deletions, total insertions, total heterozygous SNPs, total homozygous SNPs, total transitions, and total transversions. For the number of total singletons (SNPs), samples were removed that were more than eight MADs above the median. For the ratio of heterozygous to homozygous SNPs, samples were only removed that were more than four MADs above the median, so as not to remove samples that simply had high autozygosity.

We used GEL individuals with genetically inferred European ancestry, which were identified by the GEL bioinformatics team. A list of high quality LD-pruned autosomal biallelic SNPs were used in ancestry prediction: MAF >5% in both aggV2 and 1,000 Genomes project phase 3, missingness <1%, median GQ ≥30, median depth ≥30, non-palindromic, and HWE test p-value $>1x10^{-5}$. LD pruning with an $r^2$ 0.1 in 500kb windows was performed using PLINK 1.9 after removing SNPs located in long-LD regions. GCTA[9] was used to calculate 20 PCs in 1,000 Genome phase 3 samples, and GEL aggV2 samples were projected onto the PC loadings. A random forest model based on eight PCs was trained to assign an individual's probability of being from 1,000 Genomes super-populations. A cut-off of 0.8 was used to identify individuals of European ancestry. PCA within the GEL individuals predicted to have European ancestry showed population structure within this group (**Supplementary Figure 13**). To obtain a homogeneous subset that represents white British individuals, we kept samples with a probability of being from the 1,000 Genomes GBR-ancestry sub-population >0.1. Prediction of 1,000 Genomes sub-populations was performed using a random forest model trained based on PCs calculated within each predicted super-population using high quality SNPs with MAF >1%. We further removed samples with PC2 from the within-European PCA less than -0.015 to make sure the GEL samples were homogeneous (**Supplementary Figure 13**). This left 56,249 individuals in GEL.

##### ALSPAC

Data we received from ALSPAC were processed in two batches[10]. We had post-QC array data for G0 mothers and G1 children (N=17,816) in the first batch. Mothers (N=8,884) were genotyped on the Illumina Human 660W chip and children (N=8,932) were genotyped on the HumanHap550 quad chip. QC was performed by ALSPAC in the two datasets separately, as follows. Sample QC included removing samples with missingness rate <3%, heterozygosity outliers, or sex mismatches. Variant QC filters included missingness rate <5%, MAF >1%, and

HWE test P-value >1x10$^{-6}$. After merging the two datasets, SNPs with missingness rate >1% across all samples were removed.

Another 2,198 parents (G0 mothers and G0 partners) were genotyped on the CoreExome array chip in the second batch. On top of this initial QC that ALSPAC has done, we further removed seven samples with high missingness (>3%). We kept autosomal SNPs with MAF >0.5%, missingness rate <3%, and that passed the HWE test (p-value > 1x10$^{-5}$). Array data received from the ALSPAC all had genetically predicted European ancestry, so we did not perform any filtering based on genetic ancestry.

We merged the two batches and estimated genetic kinship to check sample swaps. We used KING[11] to estimate kinship using 113,090 overlapping SNPs. We removed 152 samples who had unexpected first-degree relationships but were from different families. In addition, we removed 16 samples who did not match available exome sequencing data supposedly for the same individuals; we did not remove array samples with mismatched exome data when they had a first-degree relative in the array dataset who could confirm their kinship. This left 17,656 and 2,183 samples from the two batches respectively. Among them, 8,831 were children, 9,302 were mothers, and 1,706 were fathers.

#### MCS

##### Genotype chip data

We received data from MCS for 21,181 samples genotyped using the GSA array chip that passed initial sample QC[12]. Samples with missingness rate > 20%, high or low heterozygosity (+/- 5 SDs), and sex mismatches were removed. We kept autosomal SNPs with missing rate <5%, MAF >0.5%, and had HWE test p-value > 1x10$^{-5}$ in a subset of unrelated individuals whose genetically inferred ancestry was similar to European samples from the 1,000 Genomes project. (See below for how ancestry was predicted). For duplicated variants, we kept the one with higher call rate. We further removed 283 samples with missingness rate >5%.

To identify MCS individuals with genetically inferred European ancestry, we performed a PCA in 1,000 Genomes Project phase 3 samples and projected MCS samples onto the PC space. In the PCA, we used SNPs with MAF >5% and missingness rate <1% that passed a lenient HWE test (p-value > 1x10$^{-20}$) in all MCS samples from diverse ancestries. We matched with SNPs that passed similar QC in the 1,000 Genomes. LD pruning was performed in MCS with $r^2$ <0.2 in windows of 50 SNPs. This left 87,738 SNPs in the PCA. We applied UMAP[6] using the first four PCs which differentiated continental-level populations in the 1,000 Genomes project (**Supplementary Figure 14**). 17,599 samples clustered together with non-Finnish European samples from the 1,000 Genomes project. Among them, 17,288 individuals were reported to have White ethnicity, and we restricted to these. To get a homogeneous subset, we performed another PCA within these 17,288 samples by projecting the relatives onto the PC space calculated from the unrelated subset (**Supplementary Figure 15A**). We performed UMAP on four PCs, and kept 16,803 samples from the main cluster (**Supplementary Figure 15B**).

We next checked sample swaps using genetically inferred kinship between family members estimated using KING[11]. We removed individuals whose family relationship data were missing and individuals who had unexpected first-degree relatives: parent-offspring pairs within a family that were not confirmed by genetic data, individuals from different families had a parent-offspring relationship, singleton probands who had a full sibling assigned with a different family ID. This left 16,634 individuals; among them 6,153 were children, 6,646 were mothers and 3,835 were fathers.

##### Exome sequence data

14,791 individuals from MCS, including 7,807 children and 6,975 of their parents, were exome-sequenced using TWIST capture baits (Twist Custom Panel: Core exome plus Broad panel; Twist Design ID: NGSTECustom_0001418) and Illumina NovaSeq S4 100PE, to an average depth of ~68X. We removed samples with VerifyBamID score > 0.05 due to having possible contamination. BWA-MEM was used to map the reads to GRCh38 with BWA-MEM, then SNV and indel calling was conducted with GATK HaplotypeCaller, GenomicsDBImport and GenotypeGVCFs (GATK version 4.2.4.0), following GATK best practices. Hail v0.2.105 was used to conduct sample, variant and genotype QC, as described below.

###### *Sample QC*

For the purposes of sample QC, we first filtered the data to include only biallelic SNVs and to remove variants with an internal allele frequency of ≤ 0.001 and variants with a call rate of ≤ 0.99, which reduced the number of variants from 4,920,291 to 386,148. We merged the MCS data with data from 1,000 Genomes phase 3, retaining variants present in both. We then removed variants that had low call rate (< 0.99), low allele frequency (< 0.05) or low Hardy-Weinberg equilibrium p-value (< $1x10^{-5}$), variants in long range linkage disequilibrium regions and palindromic SNVs. We ran a PCA using Hail's hwe_normalized_pca function, and then used gnomad's `assign_population_pcs` function on first ten principal components to predict which superpopulation (European, South Asian, East Asian, African, American, or other) each MCS sample was most similar to. 12,851 MCS samples were assigned as being most similar to the European samples from 1,000 Genomes.

Next we ran the `sample_qc` function in Hail and stratified the output by superpopulation. We first removed calls with DP (depth) < 20, GQ (genotype quality) < 20 or VAF (variant allele fraction) < 0.25, and then calculated the following metrics per sample: number of SNVs, Transition/Transversion ratio, het/hom ratio, heterozygosity rate, number of transitions, number of transversions, number of insertions, number of deletions, and insertion/deletion ratio. We filtered out 302 samples who fell outside of the median +/-4 median absolute deviations compared to samples from the same superpopulation for at least one metric.

###### *Variant and genotype QC*

For variant QC, we used a random forest model trained on various metrics to distinguish likely true positive from likely false positive variants. Variants in the following high-quality datasets were identified in our data and treated as true positive variants:

- High confidence sites discovered in 1,000 Genomes
- SNVs found in 1,000 Genomes that are present on the Omni 2.5 genotyping array
- Indels present in the Mills and Devine data [13]
- SNVs and indels from HapMap3

As false positive variants, we took variants failing this set of hard filters: QD (quality by depth) < 2 or FS (FisherStrand i.e. Phred-scaled p-value of Fisher’s exact test to detect strand bias) > 60 or MQ (mapping quality) < 30.

We trained a random forest model on chromosome 20 using the true positive and false positive annotations above, then applied it to the whole dataset. Most of the features used in the random forest were those used by gnomAD, and they are listed here: QD (quality by depth), meanHetAB (mean heterozygous allele balance), is_CA (is a C>A SNV), SOR (strand odds ratio), variant_type (SNV/indel/multiallelic SNV/multiallelic indel/multiallelic mixed), ReadPosRankSum (Rank sum test for relative positioning of REF versus ALT alleles within reads), was_split (is a multiallelic site), has_star (alleles at this side include a ‘*’ allele), n_alt_alleles (number of ALT alleles at a site), MQ (mapping quality), MQRankSum (Rank sum test for mapping qualities of REF versus ALT reads), allele type (SNV/insertion/deletion), and was_mixed (Multiallelic site containing SNV(s) and indel(s)) .

We included the metrics is_CA and meanHetAB in order to remove a specific artefact in the dataset characterized by a preponderance of C>A errors, which arose through a step in library preparation.

We ranked the variants by their random forest score and binned them. To decide on provisional random forest score thresholds for SNVs and indels, we manually evaluated plots of the cumulative number of true positive variants per bin and the cumulative number of false positive variants per bin (for both SNVs and indels), and of the transmitted/untransmitted ratio for synonymous singletons (SNVs only; i.e. those seen in only one parent in the sample, using 3,132 trios).

To decide on suitable hard filters for variants and genotypes, we tested different combinations of random forest bin (i.e. a variant-level metric) with various genotype quality metrics: DP (depth), GQ (genotype quality) and HetAB (heterozygous allele balance i.e. the fraction of reads carrying the ALT allele at a heterozygous genotype). For variants passing a given random forest bin filter, we set genotypes to missing if they had GQ, DP or HetAB less than the specified threshold. For each combination of filters, we calculated various metrics:

- precision and recall of variants found in the Genome in a Bottle sample NA12878
- the proportion of true positive and false positive variants from the random forest annotation remaining
- the ratio of transmitted to untransmitted variants for synonymous singletons

For SNVs, the final filters chosen were: random forest bin < 82, DP < 5, GQ < 15, and HetAB < 0.2. This gave a precision and recall of 0.931 and 0.953 respectively, captured 97.87% of true positives and 0.27% of false positives, and gave a transmitted:untransmitted ratio of 0.998 for synonymous singletons. For indels, the final filters were: random forest bin < 58, DP < 10, GQ < 20, and HetAB < 0.3. This gave a precision and recall of 0.774 and 0.691 respectively, and retained 91.185% of true positives and 0.274% of false positives.

#### Identifying relatives across cohorts

##### Identifying DDD-GEL duplicates and relatives

We suspected that there would be overlapping patients and relatives between DDD and GEL, and wanted to remove these to ensure that the samples analysed from the two cohorts were independent. Since the GEL data cannot be removed from the GEL Research Environment, we moved DDD genetic data from English and Welsh samples into it after obtaining ethical permission. We did not have consent to move DDD Scottish samples to the GEL Research Environment, so we could not remove GEL samples who were related to them. Thus, we removed Scottish individuals from the DDD cohort in GWAS and PGS analyses, and focused on identifying and removing GEL participants who were duplicates of or related to the remaining individuals from DDD. We used two approaches to identify DDD duplicates, siblings and more distant relatives in GEL (described below), and removed GEL individuals who were identified as related to DDD individuals by either approach.

We first used the DDD exome-sequence data to identify overlapping samples between DDD and GEL, and pairs of individuals from the two cohorts who were siblings. DDD exome-sequenced samples (N=32,369) were used to create informative genotype barcodes (2,466 SNPs). After excluding probands from Scotland, Northern Ireland and Dublin (whom we assumed would have not have been cross-recruited to GEL), 11,941 DDD probands were transferred to the GEL Research Environment and matched using bcftools gtcheck -e 0 --genotypes DDD.bcf GEL-sample.vcf.gz. To account for missing data, the average number of mismatching genotypes was then used to determine identical samples and siblings. We removed 1,752 GEL patients who were identified as duplicates of DDD probands and ten GEL participants who were identified as siblings of DDD patients. (Using the array data which had higher resolution (see below), two of the putative duplicates and one of the putative siblings were subsequently determined to actually be siblings and second-degree relatives of the DDD patients, respectively.)

We moved array data for 18,569 participants from DDD excluding those recruited from Scottish centres and 9,270 UKHLS controls into the GEL Research Environment. We matched SNPs from the three array chips with GEL, which left 85,092 SNPs. Up to third-degree relatives (kinship coefficient > 0.0442) were identified using KING v2.2.4[11]. Among the GEL participants with genetically-predicted European ancestry, we further removed 2,525 individuals who were related to DDD participants as well as 235 individuals who were related to UKHLS controls.

##### Integration of birth cohorts and DDD

We removed individuals from ALSPAC and MCS who were related to each other (across cohorts) or to DDD individuals. To do that, we first merged array data across DDD (CoreExome, GSA, and Omnichip), ALSPAC (both batches), and MCS (GSA). There were 45,295 SNPs that passed QC in all datasets. We compared the kinship estimated using this list of overlapping SNPs with that estimated from all available SNPs within MCS, and found that we could infer kinship relationships up to second-degree relatives accurately using the small number of overlapping SNPs. We then removed 33 samples from ALSPAC and 46 samples from MCS that had a second-degree relative or closer in DDD. We had 1,459 and 2,523 parent-offspring trios in ALSPAC and MCS, respectively. We finally removed samples from MCS or ALSPAC to make sure that children in those trios from both cohorts were unrelated, and that parents in the trios were also unrelated with other parents, resulting in 1,434 and 2,498 trios from ALSPAC and MCS, respectively.

#### Defining trio sample sets for analysis in DDD and GEL

The procedure used for filtering trios used in DDD and GEL is shown in **Supplementary Figure 16** and described below.

##### DDD

We combined trios with GBR ancestry across GSA and OmniChip arrays, then kept unrelated trios (up to three degrees of relatedness, as determined using KING[11]). We removed trios recruited from Scottish centres for the reason described above in the section on "Identifying relatives". We then subset to trios where the proband had a neurodevelopmental condition. We then split trios into those with both parents unaffected and those with one or both parents affected. These were then categorised as genetically diagnosed or undiagnosed. Among the undiagnosed trios, we looked at trios in which the proband had a rare inherited damaging variant in either a constrained gene or a dominant DD-associated gene with a loss of function mechanism. We also looked at trios with a *de novo* diagnosis.

##### GEL

We used participant data and aggregate gVCF sample statistics from the GEL main programme version 13. We identified trios, and subset to those in which each member was whole genome sequenced and the genetically-inferred sex was consistent with the phenotypic sex. We then removed trios in which any member was already in DDD or related to DDD participants up to three degrees of relatedness. These were further subset to trios in which all individuals had GBR ancestry and probands had a neurodevelopmental condition. We then kept the maximal number of trios for which the probands were not related with each other (up to third degree) and none of the parents were related to another parent. We then identified trios with unaffected parents and trios with one or both parents affected. These were then categorised into genetically undiagnosed and diagnosed. Similar to DDD, we looked at undiagnosed trios in which the proband had a rare damaging variant, as well as trios with a *de novo* diagnosis.

#### Calculating polygenic scores

Weights for polygenic scores were estimated using LDPred[14]. We used an LD reference panel for 1,054,330 HapMap3[15] variants based on 5,000 unrelated individuals of white British genetically-inferred ancestry from the UK Biobank[16].

For all scores except that for schizophrenia, we used an LD radius (--ldr) of M/3000, where M is the number of matched SNPs. In the case of schizophrenia, the singular value decomposition (SVD) did not converge, so we used an LD radius of 300. For all scores, as the traits are highly polygenic, we assumed a prior fraction of causal variants to be 1. Once the weights were generated, we calculated polygenic scores for individuals in DDD, GEL, and control cohorts, using the --score function in PLINK v1.9, which calculates the weighted sum of genotypes across a set of SNPs for each individual.

To make PGS comparable across different cohorts, we started from the same 4,570,898 SNPs that were well-imputed in all array cohorts (Minimac4 $R^2 > 0.8$), passed QC in GEL aggV2 samples, and had MAF >1% in all cohorts. Among these, 831,226 SNPs were in the aforementioned LD reference panel. GWAS summary statistics for years of schooling[17], non-cognitive performance of educational attainment[18], cognitive performance[17], schizophrenia[19], and neurodevelopmental conditions[1] were matched with the list of overlapping SNPs (**Supplementary Table 14**). The PGS for neurodevelopmental conditions ($PGS_{NDC,DDD}$) was evaluated in the DDD Omnichip samples and the GEL samples which were not in the GWAS.

To make PGS comparable across cohorts (DDD, GEL, UKHLS, MCS and ALSPAC), we performed a joint PCA across all included samples with genetically-inferred European ancestries and adjusted the raw scores for 20 PCs. We firstly performed a PCA in array samples (i.e. all cohorts except GEL). We focused on the 45,295 SNPs that passed QC in all datasets and applied LD pruning (pairwise $r^2$ <0.5 in batches of 50 SNPs with sliding windows of 5) after removing SNPs in long LD regions, which left 39,784 SNPs in PCA. We calculated PCs in a subset of unrelated individuals (removed up to second-degree relatives) and projected the remaining array samples to the PC space. We then moved the PC loadings to the GEL Research Environment and projected GEL samples with GBR ancestry onto the same PC space. (The rationale for this was that data governance constraints prevented us from moving individual-level ALSPAC and MCS data to the GEL Research Environment.) We regressed out 20 PCs from the raw PGS in array samples and used the same linear regression coefficients to adjust the raw PGS in GEL samples. For all analyses, residuals were scaled so that the combined set of unrelated control samples from GEL and UKHLS (or GEL only for $PGS_{NDC,DDD}$) had mean = 0 and standard deviation = 1.

#### Analyses of polygenic scores

##### PGS in DDD patients with different configurations of affected relatives

Wright *et al.* showed that being the only affected individual in one's family was associated with a higher chance of getting a genetic diagnosis[20]. Moreover, the more affected relatives the patient had, the lower the chance of receiving a diagnosis. We repeated this analysis in DDD patients affected by neurodevelopmental conditions with PGS available and compared the mean PGS of educational attainment ($PGS_{EA}$) in subgroups of patients with different configurations of affected relatives based on the number of affected parents, siblings, and more distant relatives, as shown in **Extended Data Figure 6**. We estimated the odds ratio of getting a diagnosis in these subgroups using a multiple logistic regression model following Wright *et al.*:

$$\begin{gathered} Diagnostic\ status \sim configuration\ of\ affected\ relatives\ +\ age\ +\ sex \\ +time\ since\ consent + trio\ status + FROH + number\ of\ DECIPHER\ variants \\ +number\ of\ organ\ systems\ affected + death + neonatal\ ICU\ stay \\ +gestation\ weeks\ + severity\ of\ intellectual\ disability\ or\ developmental\ delay\ (ID/DD) \\ +presence\ of\ seizures + reminiscent\ syndrome + recruiting\ centre + maternal\ diabetes \\ +maternal\ use\ of\ antiepileptic\ drugs + maternal\ history\ of\ pregnancy\ loss \end{gathered}$$

We no longer controlled for ancestry because we calculated PGS in only patients with genetically-inferred GBR ancestry. The reference group for the odds ratios in **Extended Data Figure 6** is the subset of patients without any affected relatives.

##### Mediation analysis to explore factors mediating the association between $PGS_{EA}$ and diagnostic status

We hypothesised that the effect of the $PGS_{EA}$ on getting a diagnosis is mediated by factors such as being in a parent-offspring trio (which likely affects diagnostic status for technical reasons) and being born prematurely (which likely has a biological effect, in that prematurity itself may be the cause of the neurodevelopmental condition rather than a monogenic cause). To assess this, we first fitted the following logistic regressions:

$$Diagnostic\ status \sim PGS\ +\ trio\ status$$
$$Diagnostic\ status \sim PGS\ +\ prematurity$$

Based on the Bayesian Information Criterion, we found that both of these models were a better fit than the baseline model $Diagnostic\ status \sim PGS$. We then performed mediation analyses using the `mediation` R package (v 4.5.0)[21] to explore whether the association between $PGS_{EA}$ and diagnostic status was medicated by trio status or prematurity. The effect (a) of the exposure (proband's $PGS_{EA}$) on the mediator (trio status or prematurity) was estimated in a logistic regression. The effect (b) of the mediator on the outcome (diagnostic status) accounting for the exposure was estimated in a logistic regression model where the outcome was regressed on both the exposure and the mediator. From the same model, we estimated the direct effect (c) of the exposure on the outcome. The indirect effect of the exposure on the outcome through the mediator was quantified as axb, which is 0 under the null hypothesis. The proportion of total effect that was mediated by a mediator was estimated axb/(axb+c). We used a bootstrap resampling method (n=1,000) in the `mediation` package to estimate the 95% confidence interval and p-value for the mediation effect.

##### Assessing collinearity in the trio model

We were concerned about the possibility of collinearity between the PGSs in the trio model (a logistic regression; results in **Figure 4**):

$$1_{NDC\ status} \sim child\ PGS\ +\ mother\ PGS\ +\ father\ PGS$$

where $1_{NDC\ status}$ is an indicator variable for whether the individual is an NDC case (1) or control (0).

The child's PGS is highly correlated with the parental PGS, and we observed correlation coefficients up to 0.59 (between children and mothers for $PGS_{EA}$). To assess the severity of collinearity in the trio model, we calculated the variance inflation factor (VIF) for the three scores for the various PGSs examined. As expected, the VIF of the child's PGS (ranging from 2.02 to 2.35; highest VIF observed for $PGS_{EA}$) is higher than that of the parental PGS (ranging from 1.49 to 1.62), meaning that the variance explained by parental PGS in the child's PGS is higher than that explained in a parent's PGS by other two PGS. However, the highest VIF amongst all five PGSs examined did not exceed the commonly-used cutoff of 5, suggesting that collinearity is not too concerning.

##### Testing whether prematurity mediates the effects of non-transmitted alleles

To test the hypothesis that the effects of non-transmitted alleles associated with educational attainment and cognition might be mediated by prematurity, we reran the trio model (**Figure 4**) in several ways using a subset of trios for which data on gestational age were available, namely undiagnosed cases from DDD (N=1,521 trios) and control trios from MCS (N=2,451; **Supplementary Figure 5**, **Supplementary Note 5**). We ran the trio model adjusting for prematurity as a binary covariate (<37 full weeks or ≥37 weeks), in trios excluding probands who were born prematurely (16% of cases and 6% of MCS controls), and in only premature probands. Among the probands who were born prematurely, the severity of prematurity in DDD was higher than MCS, with 5% born extremely prematurely (<28 weeks) in DDD versus 3% in MCS and 15% with a gestational age between 28 to 32 weeks in DDD vs 12% in MCS. To account for this, we also ran the trio model in premature probands controlling for the severity of prematurity as a categorical covariate indicating whether the proband belonged to any of the following three categories: with a gestational age <28 full weeks, 28 weeks to <32 weeks, or 32 weeks to <37 weeks.

#### Mendelian randomization

To explore causal relationships between educational attainment, prematurity and neurodevelopmental conditions, we ran standard two-sample Mendelian randomization (MR) and the Latent Heritable Confounder MR (LHC-MR) method using the lhcMR v0.0.0.9000 package in R [22]. We used the GWAS summary statistics listed in **Supplementary Table 14**. For standard MR analysis, SNPs were chosen as genetic instruments for a given exposure using LD clumping with the 1000 Genomes EUR superpopulation as the reference (LD $R^2 < 0.001$ by 10 MB windows) and a p-value cutoff of $5 \times 10^{-8}$, resulting in up to 297 SNPs for educational attainment, three SNPs for preterm delivery, and two SNPs for neurodevelopmental conditions. We performed univariate MR using four methods when there were three or more genetic instruments: inverse-variance weighted, weighted mode, weighted median, and MR Egger. Only the inverse-variance weighted method was run when there were two genetic instruments. In addition, we also assessed bi-directional causal effects between two given traits using LHC-MR, which allows for a reverse causal effect and a latent heritable confounder of the exposure-outcome relationship. LHC-MR used about 4 million genome-wide SNPs with LD score calculated. As recommended by the authors, to avoid potential bias, LHC-MR was performed when the total SNP heritability estimate (by LD score regression) of the exposure was >2.5%, which was the case for educational attainment (11%) but not for prematurity (1.2%). Thus, educational attainment was tested as the exposure for both prematurity and neurodevelopmental conditions, but allowing for a reverse causal effect in each case.

#### Construction and incorporation of weights for the Millennium Cohort Study

The Millennium Cohort Study (MCS) deliberately oversampled minority ethnic and disadvantaged individuals [23], which can lead to biased estimates of mean trait values, including PGSs, within the sample (sampling bias). In addition, nonrandom missingness in each wave of data collection (including the collection of DNA for genotyping) due to biased attrition can also lead to biased estimates of mean trait values (non-response bias). MCS developed sampling weights to adjust for the initial non-random sampling. To correct for non-response bias, we produced non-response weights using inverse probability weighting as previously described[23,24]. First, we conducted a logistic regression on whether a MCS child had genotype data using the following covariates collected at the first study sweep, which had minimal missingness:

*has_genotype_data ~ housing_tenure + parental_years_in_education + language_spoken_at_home + study_strata + single_parent_status + breastfeeding_status*

The variable study_strata consists of nine categories of individuals from each country in the UK (England, Wales, Scotland, Northern Ireland), each of which was stratified by classification as advantaged or disadvantaged, plus an additional classification of “ethnic” sampling only in

England. The variables from which these covariates were taken were as follows: ADROOW00, APLFTE00, ADHLAN00, PTTYPE2, ADHTYS00, ACBFEV00. Individuals with any missingness for these variables were excluded from the weighting procedure. As these variables were collected at the first sweep of the study, missingness was low, with >96% of individuals having complete data for the selected variables.

We fitted this model to predict who was within the sample of unrelated GBR-ancestry individuals with genotype data (N=5,884 of 6,036 children who had no missingness on the variables above), and separately to predict who was within the subset of these that additionally had genotype data on both parents (N=2,445 of 2,498 trio children who had no missingness). In the latter model, we included an additional covariate for whether the child had any genotype data. These models had a Nagelkereke $R^2$ of 0.51 and 0.42, respectively.

We then extracted the predicted value for each genotyped individual as the probability of being genotyped (or the probability of being in a trio from which all three members were genotyped) and used the inverse of that probability as the non-response weight per individual. Intuitively, those with lower predicted probabilities for having genotype data but nonetheless were genotyped were assigned higher weights. We removed individuals with weights more than three standard deviations above the mean, as these likely represent phenotyping errors driving erroneously low non-response probabilities. This removed fourteen individuals for the weights in all genotyped individuals and three when restricting to those in trios. To calculate an overall weight taking into account both sampling and non-response bias, we multiplied the sampling weights provided by MCS with the non-response weights we had calculated.

To calculate the mean PGS for the groups shown in **Extended Data Figure 5C**, we fitted a linear regression for the PGS in R (formula: 1 ~ PGS) with no covariates and included the weights using the “weights” argument, such that the intercept for the model returns the weights-adjusted mean for the sample. To calculate the weighted correlation between PGS and rare variant burden score (RVBS) shown in **Supplementary Figure 7**, we fitted a linear regression in R regressing scaled PGS on scaled RVBS (formula: scale(RVBS) ~ scale(PGS)) incorporating the weights similarly, such that the regression coefficient returns the weights-adjusted correlation.

### Supplementary Figures

*Figure S1*

**Supplementary Figure 1.** Pearson correlations between the five polygenic scores (PGS) used throughout this paper. Correlations were estimated in the following three subgroups: probands with neurodevelopmental conditions (NDCs) regardless of trio status (N=3,618 from GEL and N=6,883 from DDD; N=597 in DDD excluding GWAS samples), parents of probands from 2,174 DDD trios and 2,390 GEL trios, and controls individuals from GEL (N=13,667) and UKHLS (N=9,270). EA: educational attainment; CP: cognitive performance; NonCogEA: the non-cognitive component of EA; SCZ: schizophrenia; NDC,DDD: neurodevelopmental conditions, with the GWAS conducted in DDD versus the UK Household Longitudinal Study, and the polygenic score tested only in samples excluded from the GWAS (GEL and DDD OmniChip).

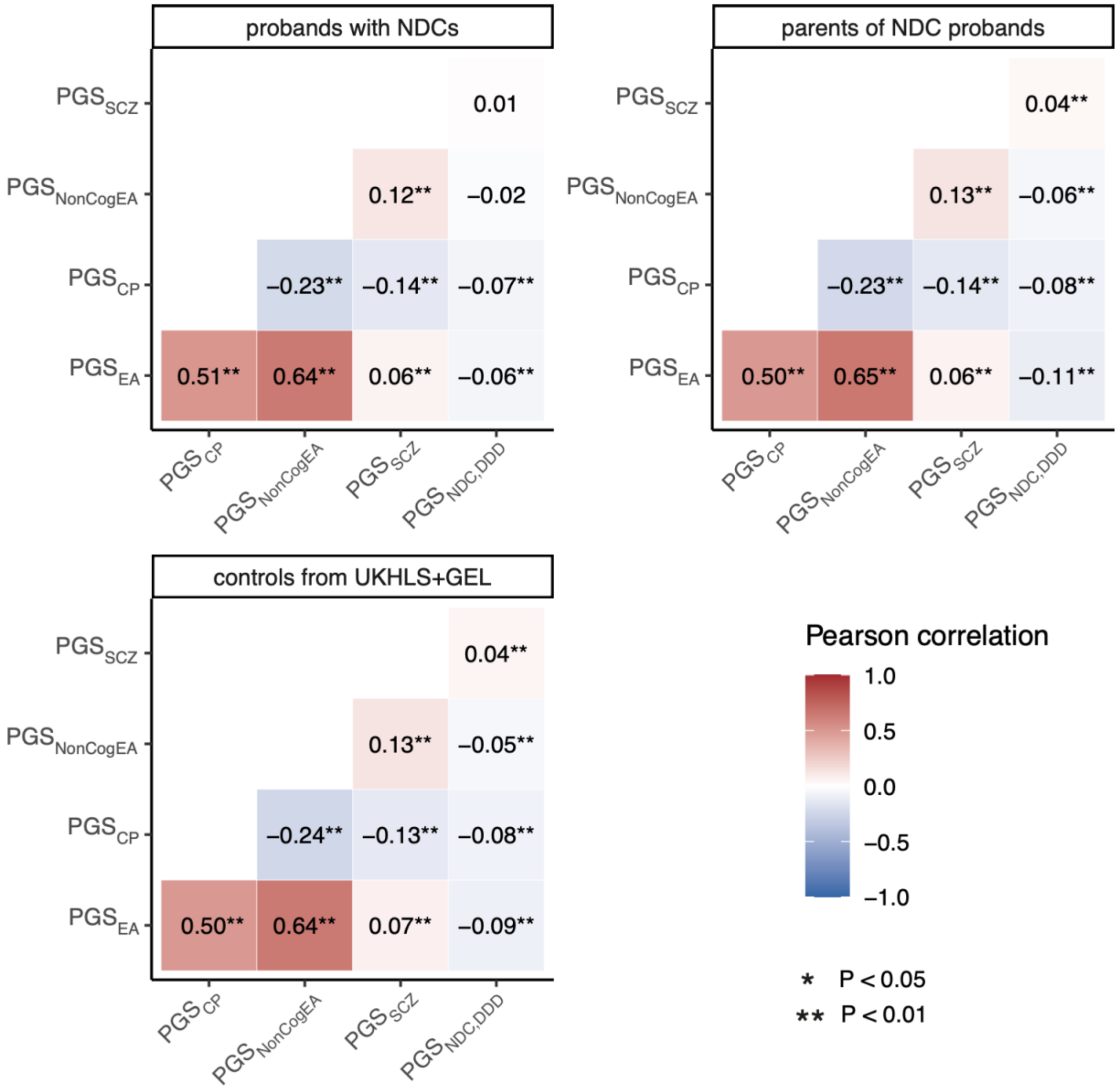

*Figure S2*

**Supplementary Figure 2**. Polygenic transmission disequilibrium test (pTDT) results having excluded autistic probands, in either undiagnosed or diagnosed probands (N=1,298 undiagnosed in DDD, N=192 excluding NDC GWAS samples, N=907 in GEL; N=395 diagnosed in DDD, N=268 excluding NDC GWAS samples, N=389 in GEL).

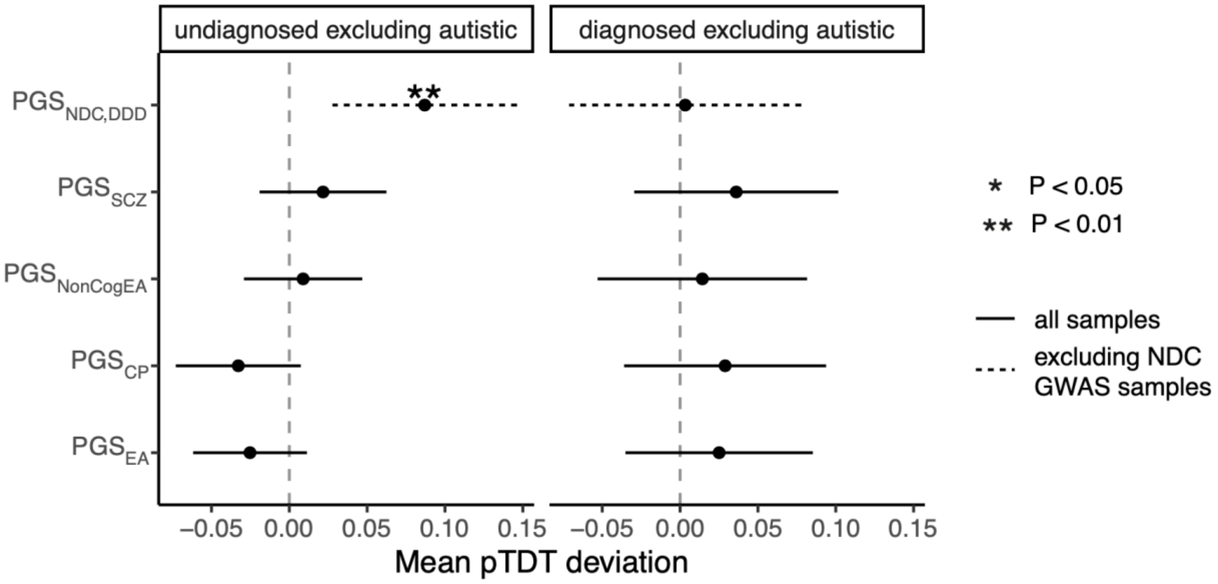

*Figure S3*

**Supplementary Figure 3**. Polygenic transmission disequilibrium test (pTDT) results in probands with a monogenic diagnosis (N=443 in DDD, N=296 excluding GWAS samples; N=507 in GEL)

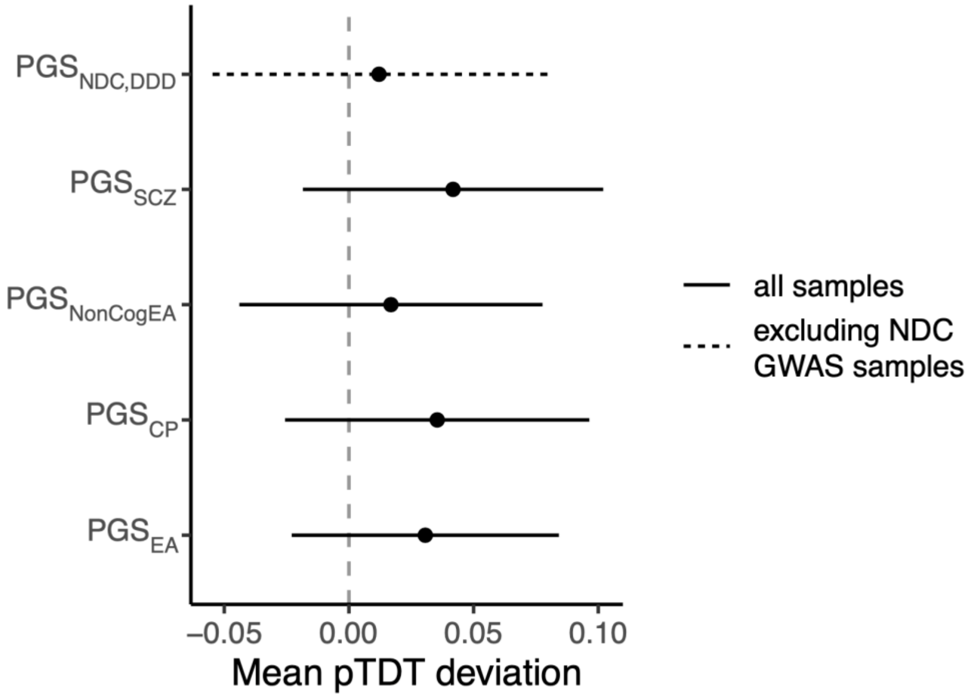

*Figure S4*

**Supplementary Figure 4.** Sensitivity analysis to assess non-transmitted coefficients and direct genetic effects of PGS in **A**) only GEL trios, **B**) in all cases with neurodevelopmental conditions versus GEL control trios, **C**) in all cases versus MCS control trios, and **D**) in all cases versus ALSPAC control trios. All cases with neurodevelopmental conditions are undiagnosed and both parents are unaffected. Y axes show effect sizes of PGSs on case/control status, testing either the child's PGS alone ("proband only"), or while additionally controlling for the parents' PGSs ("trio model"). Two asterisks indicate significance at p-value < 0.01 (Bonferroni correction for five PGSs) and one asterisk indicates nominally significant at p-value <0.05. Error bars indicate 95% confidence intervals.

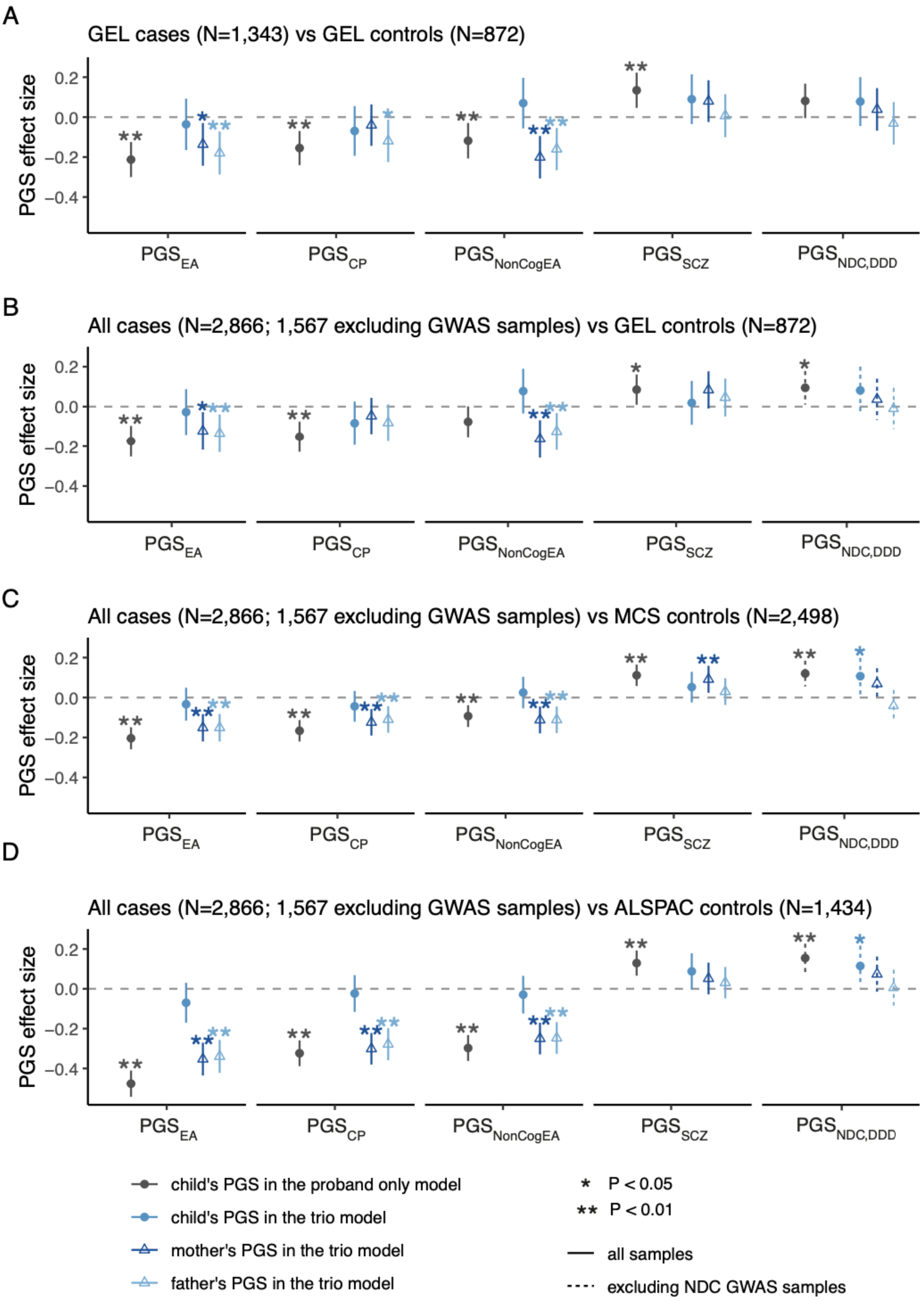

A
GEL cases (N=1,343) vs GEL controls (N=872)
B
All cases (N=2,866; 1,567 excluding GWAS samples) vs GEL controls (N=872)
C
All cases (N=2,866; 1,567 excluding GWAS samples) vs MCS controls (N=2,498)
D
All cases (N=2,866; 1,567 excluding GWAS samples) vs ALSPAC controls (N=1,434)
PGS effect size
PGS_EA
PGS_CP
PGS_NonCogEA
PGS_SCZ
PGS_NDC,DDD
child's PGS in the proband only model
child's PGS in the trio model
mother's PGS in the trio model
father's PGS in the trio model
* P < 0.05
** P < 0.01
all samples
excluding NDC GWAS samples

*Figure S5*

**Supplementary Figure 5**. Exploring whether prematurity may mediate the association between non-transmitted alleles of $PGS_{EA}$ and risk of neurodevelopmental conditions. Estimates of the effect of $PGS_{EA}$ from the "proband-only" model (grey) or the "trio model" (different shades of blue), as in **Figure 4**, run on various different subsets of probands. Three different models were run, plotted in this order: using all DDD and MCS probands either before ("DDD versus MCS") or after controlling for whether or not the proband was born prematurely ("adj prematurity"), or using DDD and MCS probands excluding those who were born prematurely ("excluding premature probands"). Error bars indicate 95% confidence intervals.

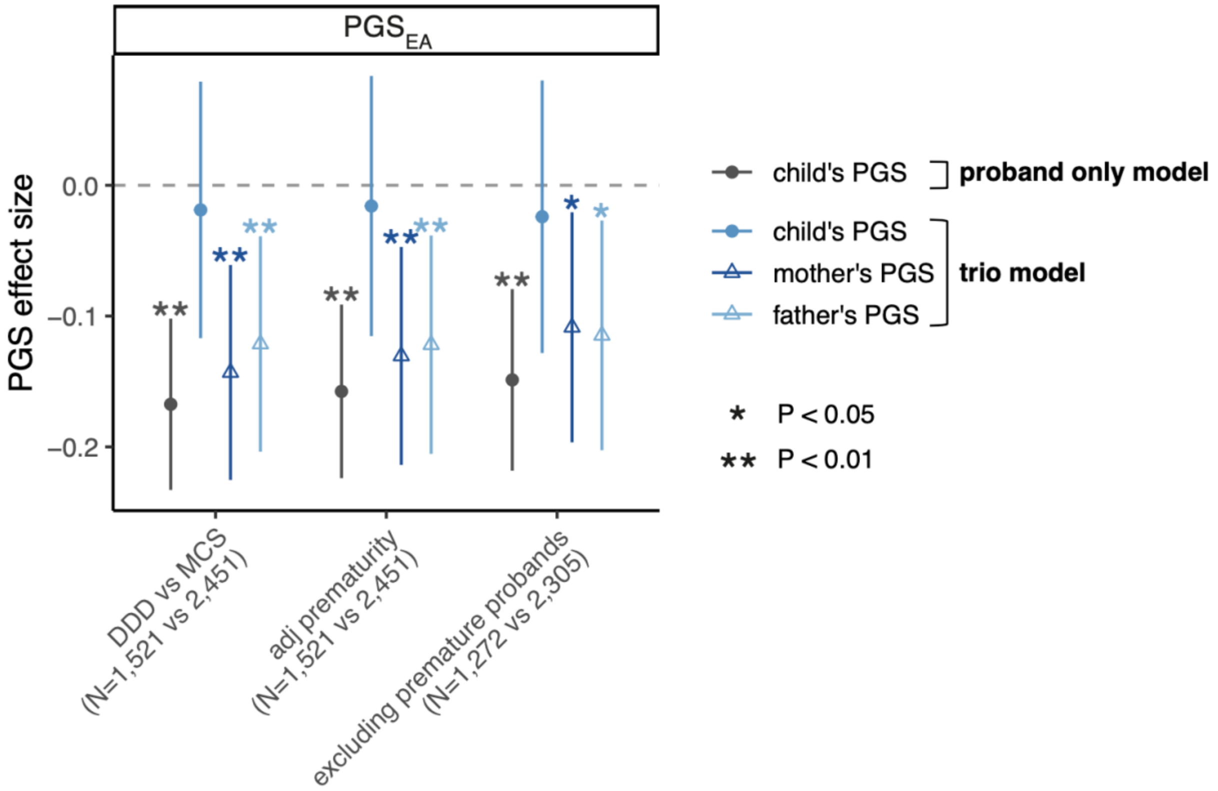

*Figure S6*

**Supplementary Figure 6.** Correlation coefficients between PGSs and the number of inherited rare coding variants defined in various ways within/between different sets of individuals, in different subsets of trios with neurodevelopmental conditions. The correlations within probands with neurodevelopmental conditions whose parents are unaffected are shown in blue (i.e. the child's rare variant burden score, RVBS, with their own PGS), and those within their parents are shown in purple. The cross-parental correlation (i.e. one parent's RVBS with the other parent's PGS) is shown in orange. We conducted the analysis either in all trios with neurodevelopmental conditions in which both parents were unaffected (**A**), in undiagnosed trios with unaffected parents (**B**) or in trios with *de novo* diagnoses and unaffected parents (**C**). RVBSs were calculated using PTVs, PTV and missense variants combined, or synonymous variants in dominant DD genes with a loss-of-function mechanism or in constrained genes. Significant correlations that pass Bonferroni correction for 30 tests (P-value < 0.0017; five PGSs, three variant types, and two gene sets) are indicated by two asterisks, and nominally significant correlations (P-value < 0.05) are indicated by one asterisk. Note that both the rare variant burden scores and PGS have been corrected for 20 genetic principal components. Error bars represent 95% confidence intervals.

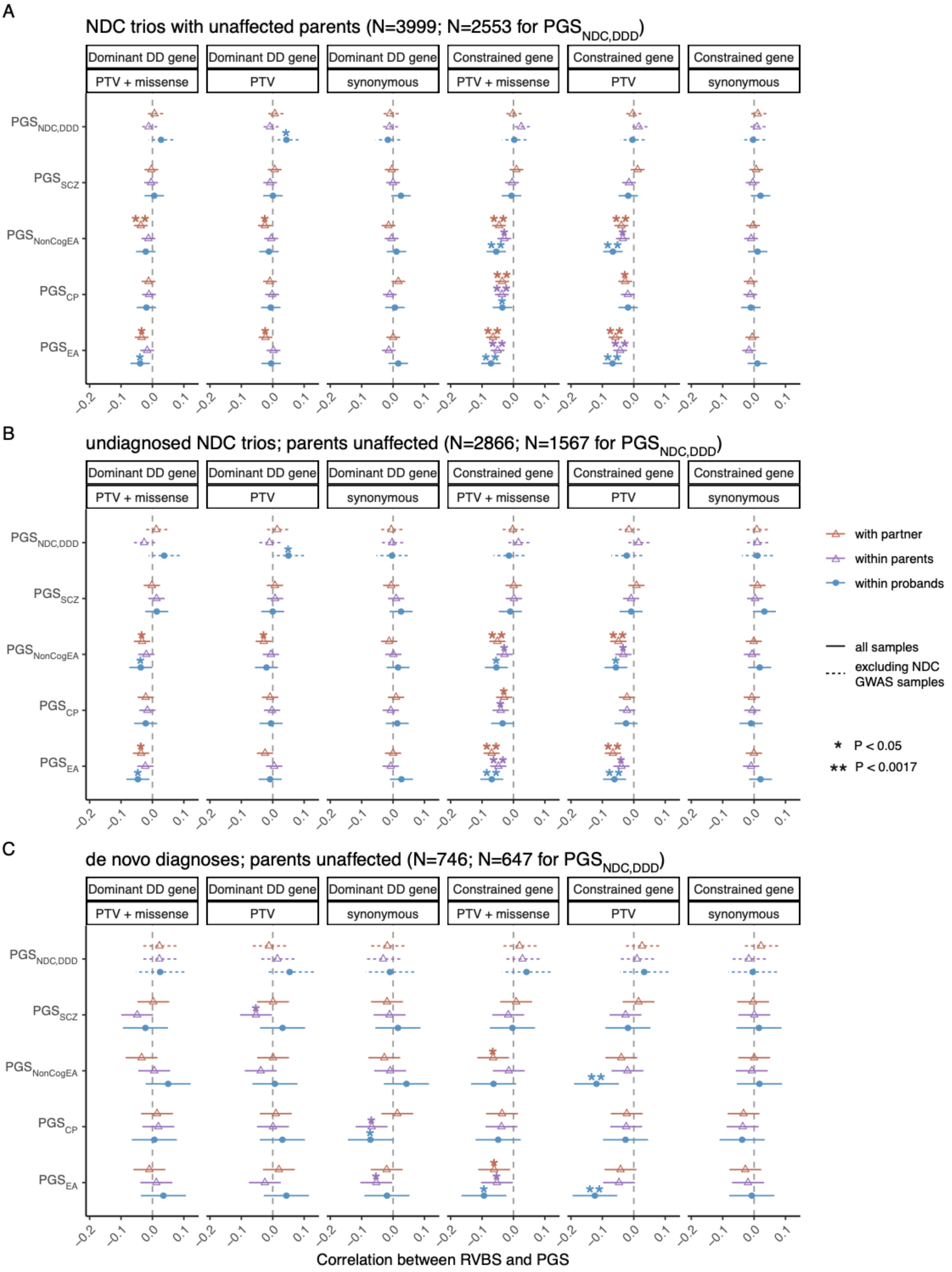

A
NDC trios with unaffected parents (N=3999; N=2553 for PGS_NDC,DDD)
B
undiagnosed NDC trios; parents unaffected (N=2866; N=1567 for PGS_NDC,DDD)
C
de novo diagnoses; parents unaffected (N=746; N=647 for PGS_NDC,DDD)
Dominant DD gene
PTV + missense
PTV
synonymous
Constrained gene
PGS_NDC,DDD
PGS_SCZ
PGS_NonCogEA
PGS_CP
PGS_EA
with partner
within parents
within probands
all samples
excluding NDC GWAS samples
* P < 0.05
** P < 0.0017
Correlation between RVBS and PGS

*Figure S7*

**Supplementary Figure 7.** Correlation coefficients between PGSs and the number of rare coding variants filtered in various ways within/between different sets of individuals, in control trios (N=2,246) from the Millennium Cohort Study (MCS). The correlations within children are shown in blue (i.e. the child's inherited rare variant burden score, RVBS, with their own PGS), and those within their parents are shown in purple. The cross-parental correlation (i.e. one parent's RVBS with the other parent's PGS) is shown in orange. RVBSs were calculated using PTVs, PTV and missense variants combined, or synonymous variants in dominant DD genes with a loss-of-function mechanism or in constrained genes. The weighted correlations after adjusting for sampling bias and non-response bias (attrition) (Supplementary Methods) are shown in darker colours. Significant correlations that pass Bonferroni correction for 30 tests (P-value < 0.0017; five PGSs, three variant types, and two gene sets) are indicated by two asterisks, and nominally significant correlations (p-value<0.05) are indicated by one asterisk. Note that both the rare variant burden scores and PGS have been corrected for 20 genetic principal components. Error bars represent 95% confidence intervals.

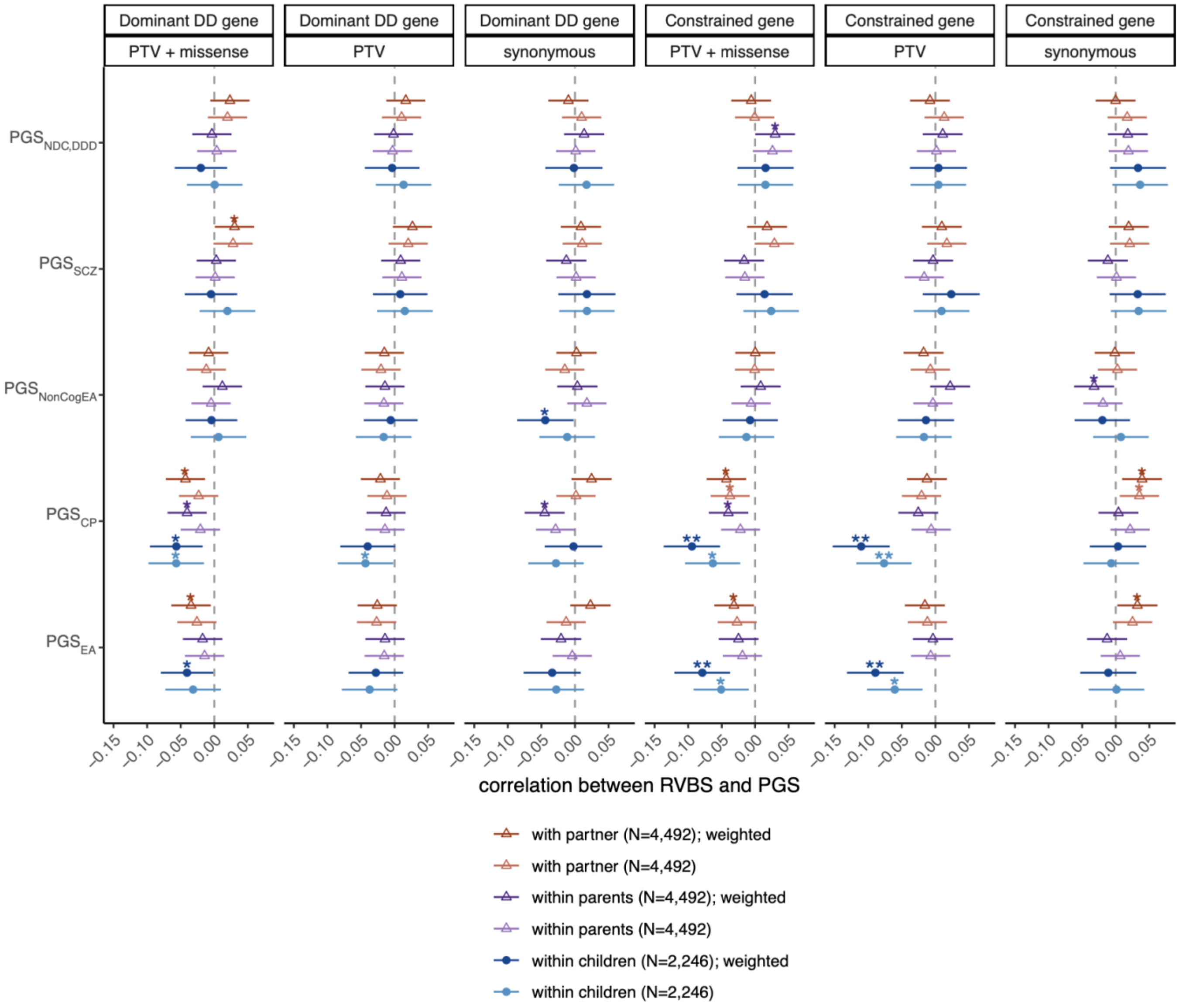

*Figure S8*

**Supplementary Figure 8.** Investigation of PGSs modifying penetrance of rare damaging coding variants within families. The plot shows results from one-sided, paired *t*-tests comparing PGSs between unaffected parents transmitting damaging rare variants and their undiagnosed children. A positive difference indicates that the unaffected parents have higher PGS than the children. One asterisk indicates a nominally significant difference; none of the differences passed Bonferroni correction for 20 tests (five PGSs, two variant types, and two gene sets). Error bars indicate 95% confidence intervals (CIs). The upper bound of CI is shown for $PGS_{SCZ}$ and $PGS_{NDC,DDD}$, for which an upper-tailed test was used and the lower bound is -infinity. The lower bound of CI is shown for $PGS_{EA}$, $PGS_{CP}$, $PGS_{NonCogEA}$, for which a lower-tailed test was used, and the upper bound is +infinity.

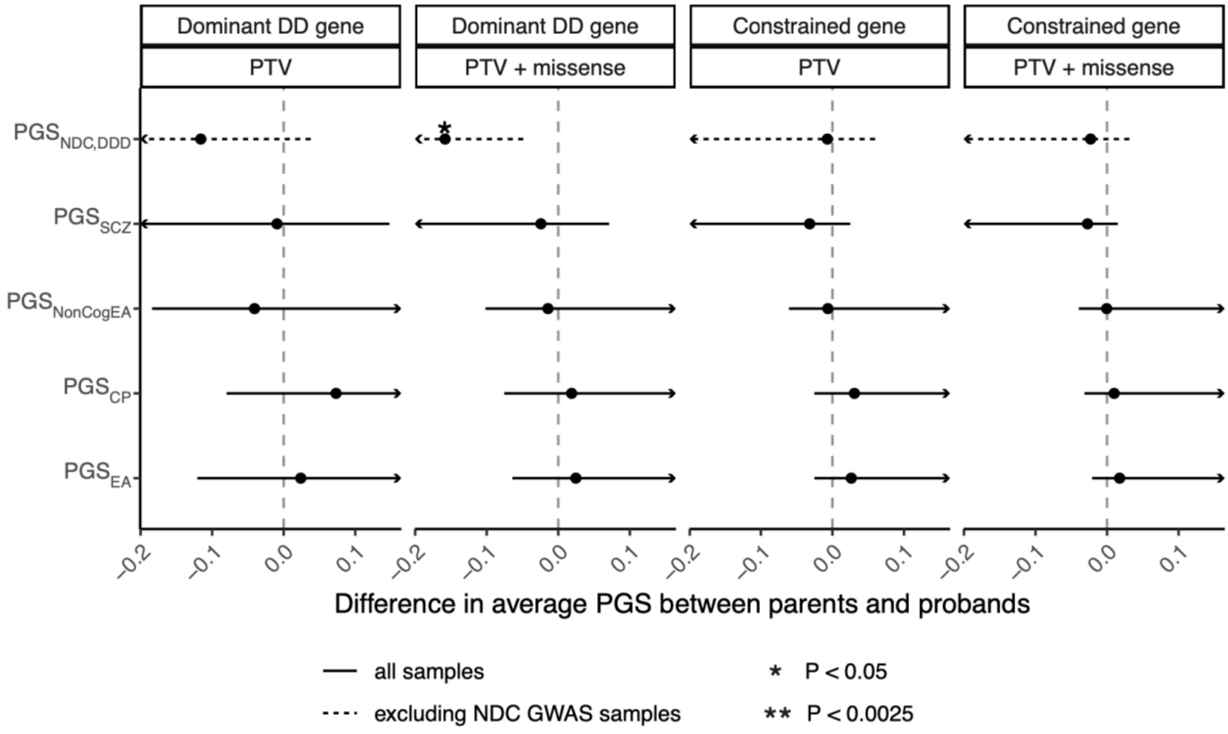

*Figure S9*

**Supplementary Figure 9**. Venn Diagrams of DDD samples by genotyping array. Overlapping proband samples (left) and overlapping parent samples (right) across three genotype array chips used in the DDD cohort.

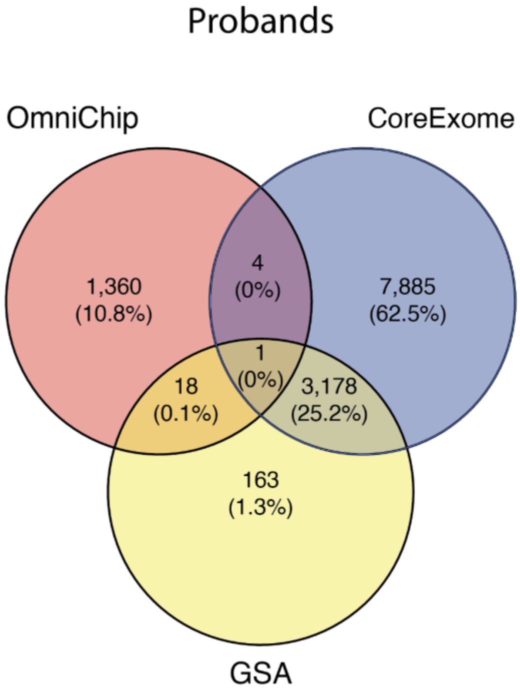

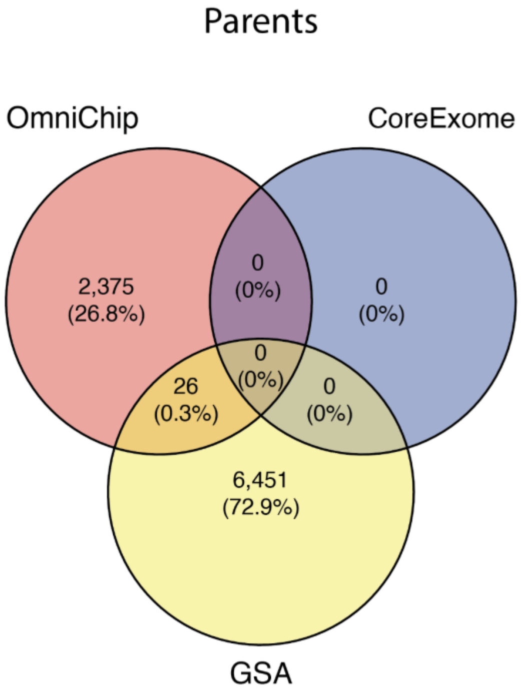

*Figure S10*

**Supplementary Figure 10.** Principal components (PCs) of DDD Global Screening Array samples (N = 9,572) and 1,000 Genomes phase 3 samples (N=2,548). DDD individuals are in black, coloured by superpopulation, with the exception of GBR-ancestry individuals. GBR: Great British, EAS: East Asian, EUR: European, AFR: African, AMR: Ad Mixed American, SAS: South Asian.

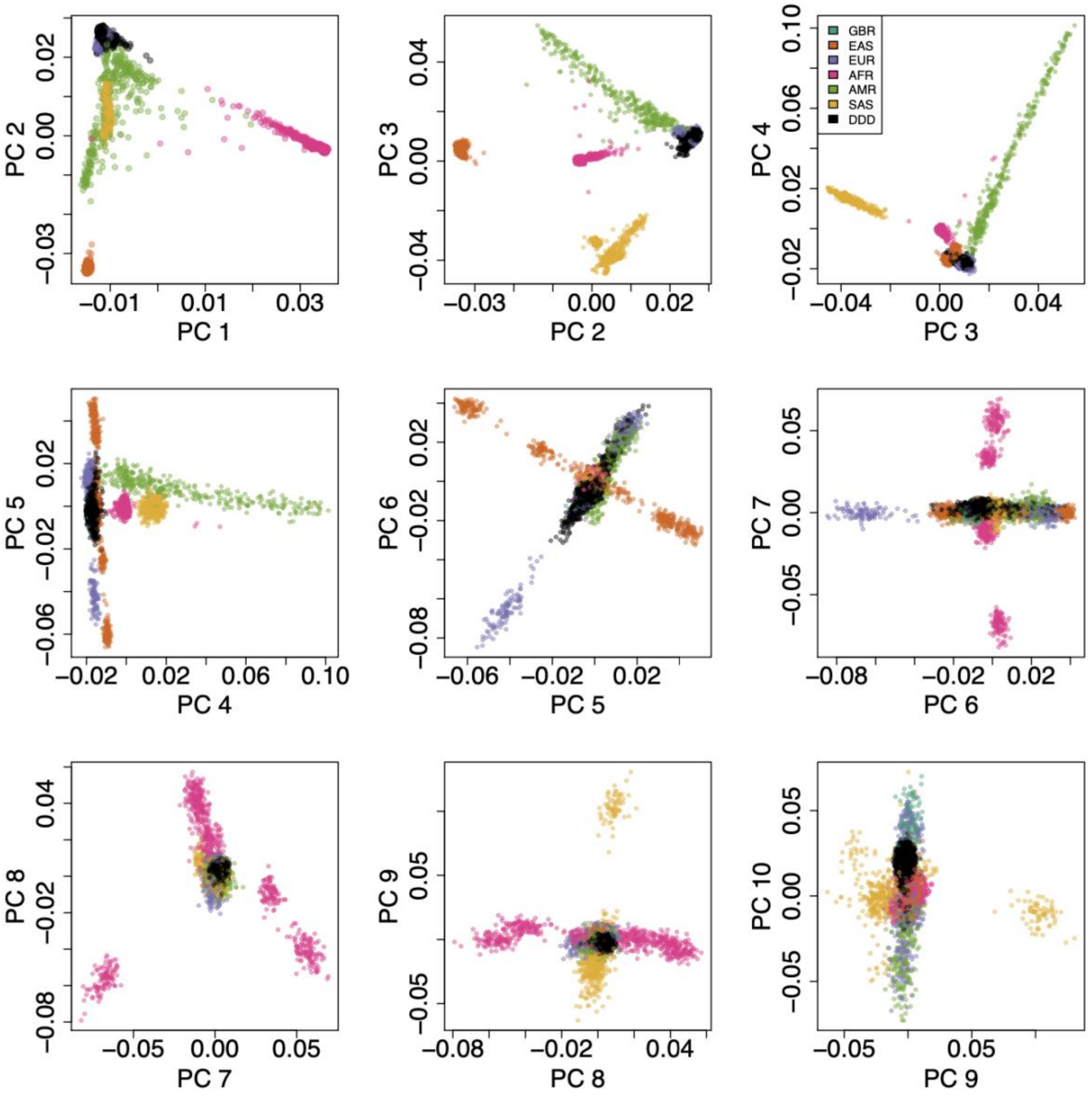

*Figure S11*

**Supplementary Figure 11.** UMAP using the first ten principal components from a principal component analysis (PCA) within loosely European-ancestry DDD samples on the Global Screening Array (N=9,534). Black lines indicate the cut-offs chosen to delineate the homogeneous European-ancestry group shown in the bottom left area of the plot, which was taken forward for analysis (N=8,489).

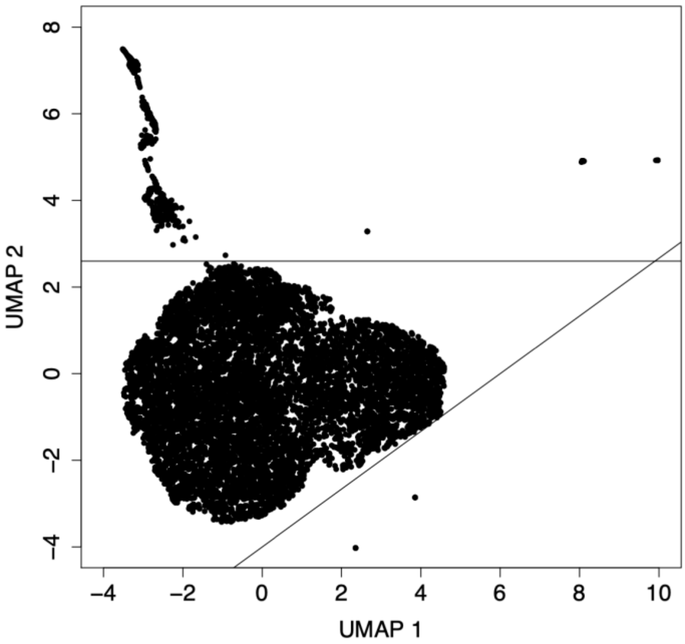

*Figure S12*

**Supplementary Figure 12.** Principal components (PCs) of European-ancestry participants in DDD genotyped on the Global Screening Array and GBR-ancestry individuals genotyped on OmniExpress chip from Niemi *et al*.[1].

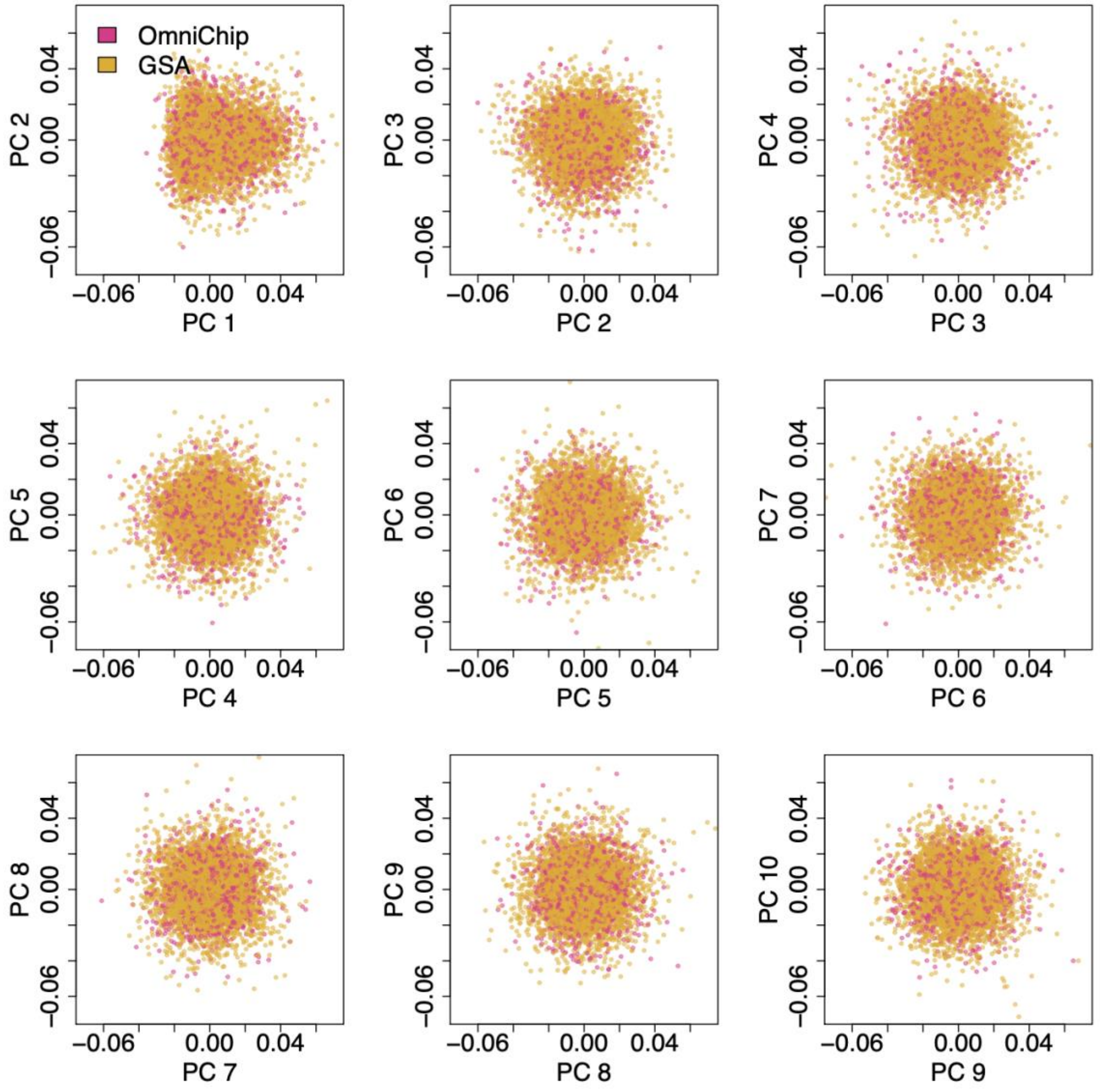

*Figure S13*

**Supplementary Figure 13**. Principal components (PCs) of GEL individuals with predicted European ancestry (N= 62,366). All individuals plotted here had a probability of being in the 1,000 Genomes EUR super population > 0.8 (according to a random forest model), and a subset of those with probability of being in the 1,000 Genomes GBR subpopulation > 0.1 (according to a random forest model) are in orange. The black line on PC 2 indicates the PC2 cut-off for individuals selected as GBR ancestry.

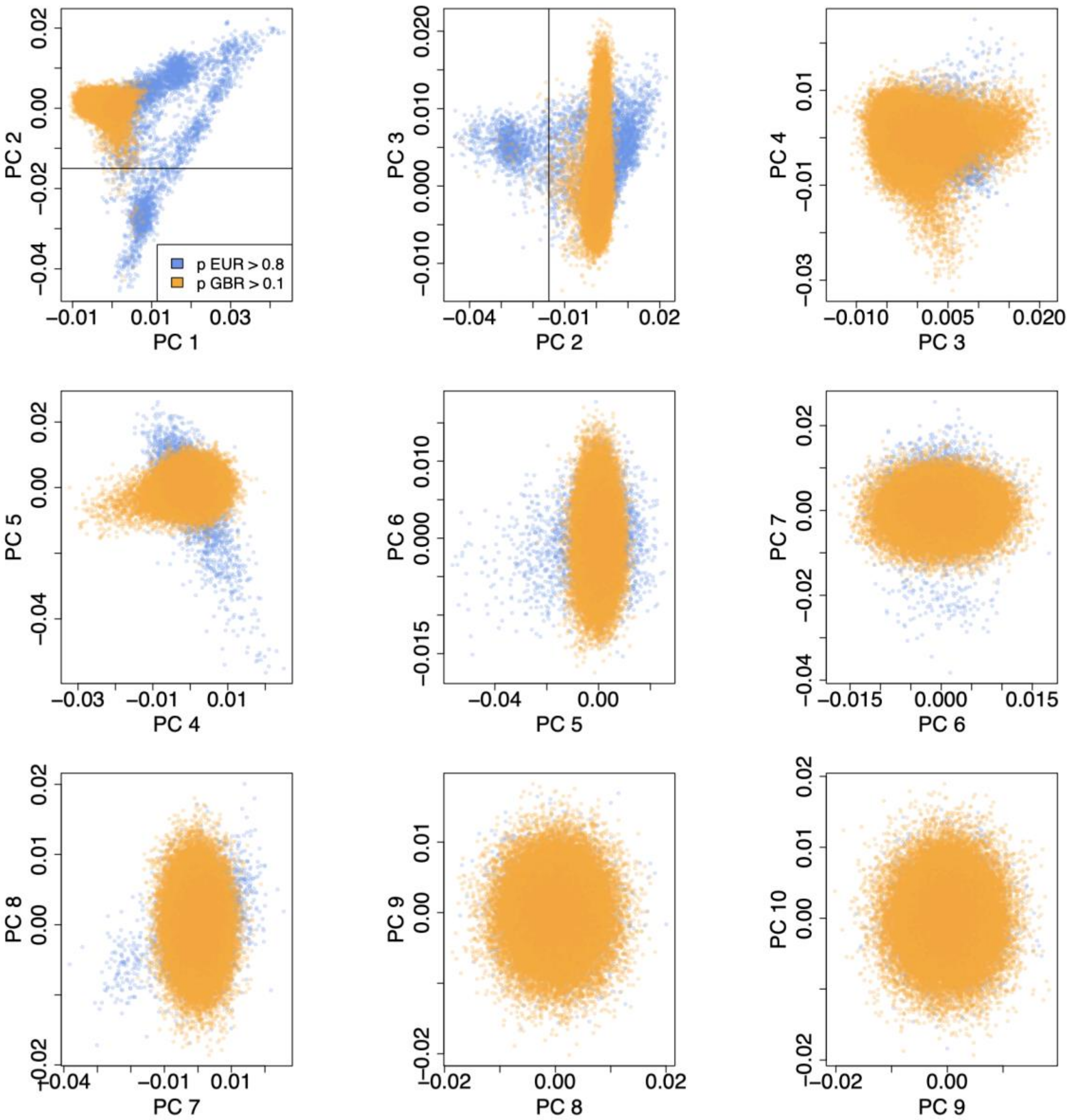

*Figure S14*

**Supplementary Figure 14. A**) Principal components (PCs) calculated in 1,000 Genomes phase 3 individuals, with MCS individuals shown in black projected to the same PC space. Colours indicate continental-level populations from the 1,000 Genomes project: European (EUR), African (AFR), Ad Mixed American (AMR), East Asian (EAS) and South Asian (SAS). **B**) UMAP using the first four PCs.

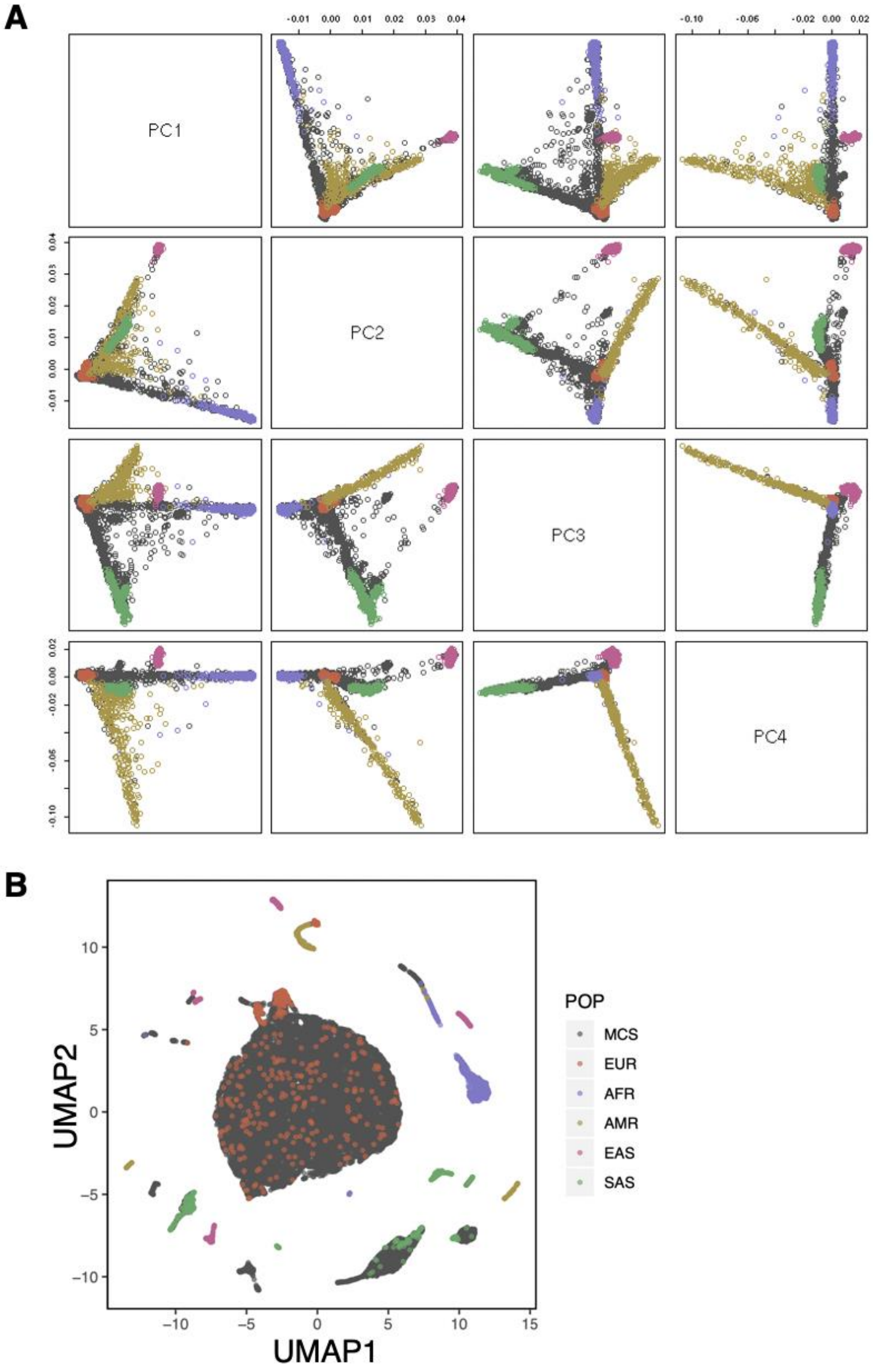

*Figure S15*

**Supplementary Figure 15. A**) Principal components (PCs) of 17,599 MCS samples who were reported to have White ethnicity clustered together with non-Finnish European samples from the 1,000 Genomes project. Red indicates outlier samples that were removed based on being below the line in panel (B). **B**) UMAP using the first four PCs.

A

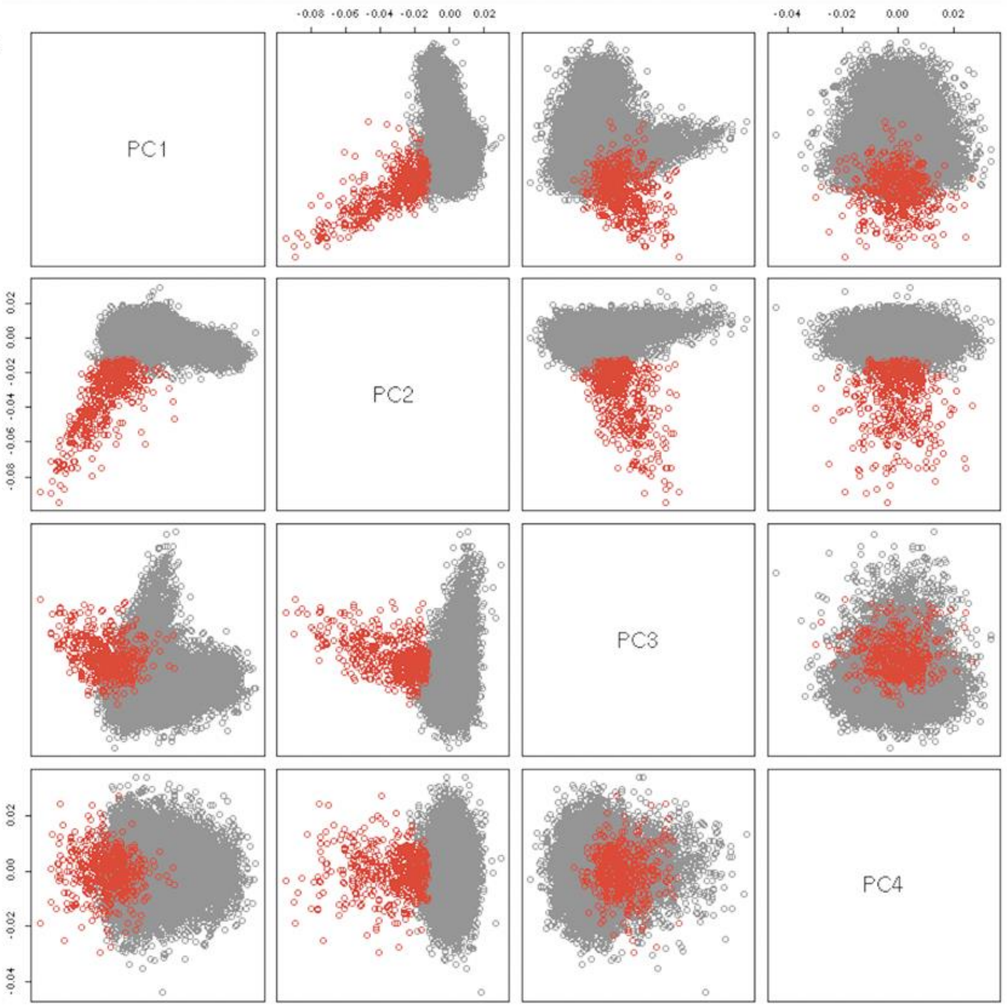

B

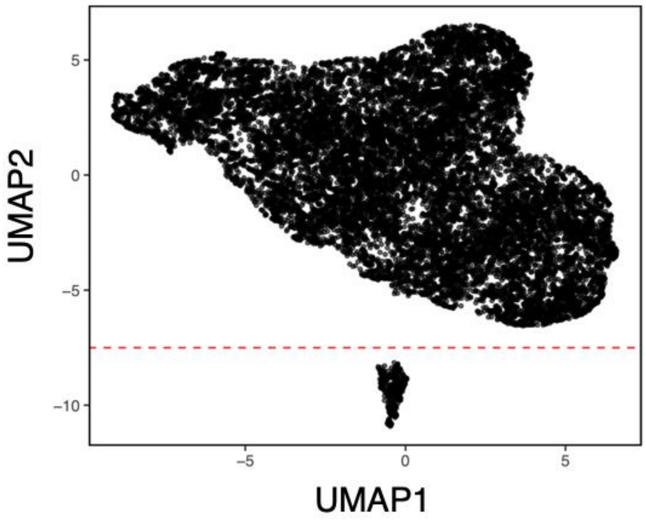

*Figure S16*

**Supplementary Figure 16.** Flow diagram of trio filtering in **A**) DDD and **B**) GEL.

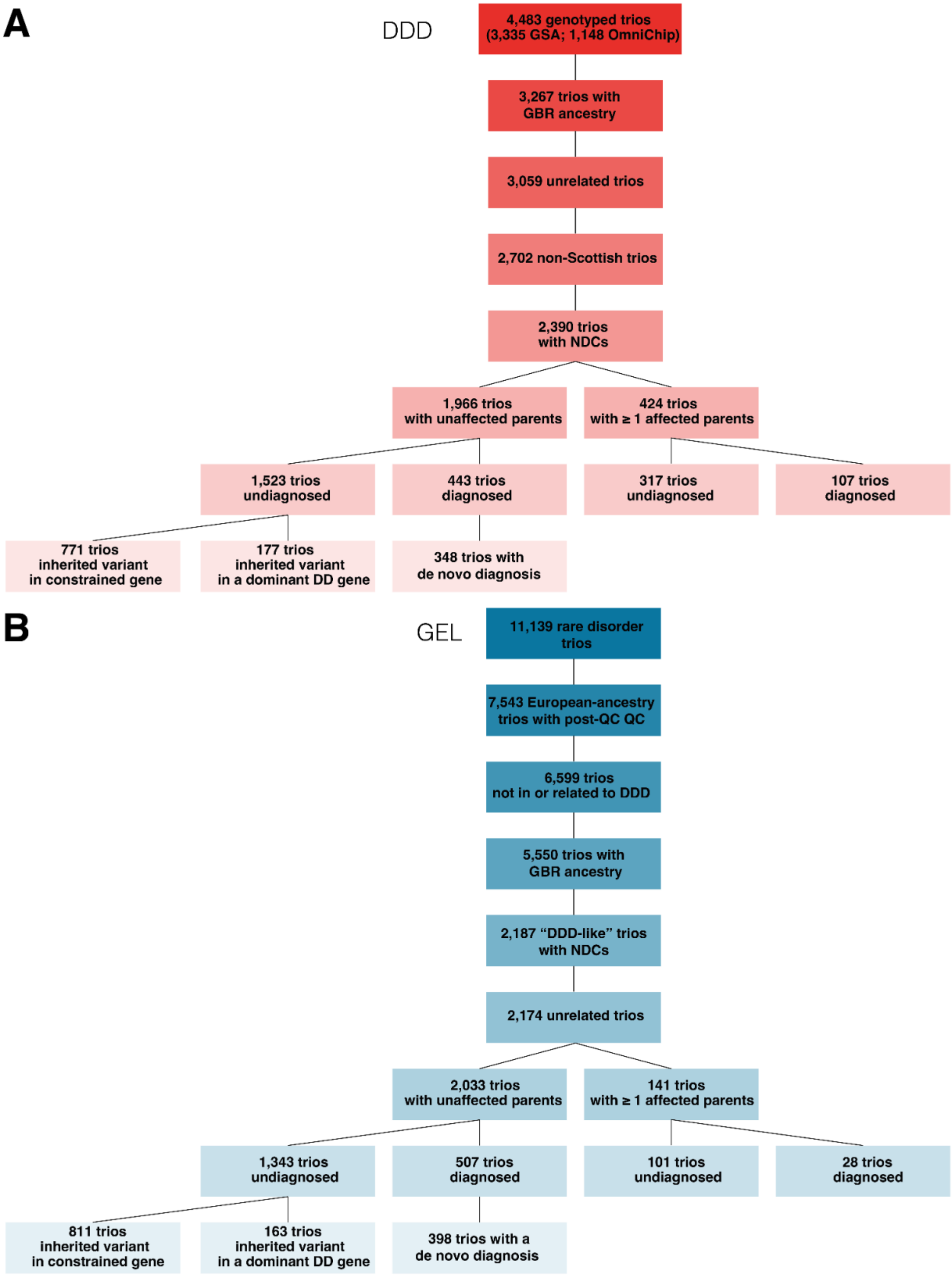

### Supplementary Tables

**Supplementary Table 1.** Number of samples used in each analysis. All individuals have genetically inferred European ancestry. Note that we did not calculate the PGS for neurodevelopmental conditions in UKHLS samples and DDD probands genotyped using CoreExome chip or Global Screening Array, since they were in the original GWAS (Niemi et al., Nature, 2018), but rather only tested it in GEL probands and in DDD probands who were genotyped using the OmniChip as well as in parents and UK birth cohorts. Scottish participants and those who are related to GEL participants were excluded from DDD when they are analysed together with GEL samples in a combined analysis.

**Supplementary Table 2.** Variance explained by polygenic scores (PGSs) for educational attainment (EA), cognitive performance (CP), the non-cognitive component of educational attainment (NonCogEA), schizophrenia (SCZ), and rare neurodevelopmental conditions (NDC,DDD) on the liability scale, either comparing DDD with UKHLS or GEL cases with GEL controls.

**Supplementary Table 3**. SNP heritability estimates using different methods. LD score regression (LDSC) was run on summary statistics from the GEL-derived GWAS, DDD-derived GWAS, and the meta-analysed GWAS. LD- and MAF-stratified GREML (GREML-LDMS)[25] and phenotype-correlation genotype-correlation (PCGC)[26] regression were run in DDD and GEL GWAS samples separately, then the SNP heritability estimates were meta-analyzed. We observed higher SNP heritability estimates using GREML-LDMS and PCGC, possibly due to the fact that LDSC uses only the subset of SNPs in HapMap (Methods) and because the heritability estimate tends to be downward-biased at sample sizes of this order[27].

**Supplementary Table 4.** Genetic correlations ($r_g$) between neurodevelopmental conditions and other brain-related traits and disorders. We calculated $r_g$ between brain-related traits and either GWAS of neurodevelopmental conditions derived from DDD or the GWAS meta-analysis of DDD and GEL using Linkage Disequilibrium Score Regression (LDSC). We also calculated $r_g$ between the GWAS meta-analysis of neurodevelopmental conditions and those brain-related traits after conditioning on educational attainment and cognitive performance using GenomicSEM. We performed two-sided z-score tests ("z_pvalue") to compare $r_g$ estimates before and after conditioning on the two traits. Confidence intervals (CI) were calculated using the standard error (se) of the $r_g$ estimate. We corrected for 13 traits using the Bonferroni approach. For each external trait, we also report SNP heritability on the observed scale by LDSC. See Supplementary Table 14 for information on external GWASs.

**Supplementary Table 5**. Two-sided *t*-tests comparing average polygenic scores (PGSs) between subsets of probands with neurodevelopmental conditions and control individuals shown in Figure 2A. The table contains comparisons (1) between subsets of probands with neurodevelopmental conditions (from DDD+GEL combined) and (2) between probands with neurodevelopmental conditions and control individuals from a combined set of unrelated individuals from GEL and UKHLS. The PGS for neurodevelopmental conditions derived from the GWAS of DDD and UKHLS samples was tested in only GEL samples and a held-out set in DDD. Nominally significant *t*-test results are indicated by one asterisk and tests that pass the Bonferroni correction for five PGSs are indicated by two asterisks. The last column highlights comparisons that were mentioned in the main text.

**Supplementary Table 6**. Two-sided *t*-tests comparing polygenic scores (PGSs) between different control cohorts and subsets thereof, subsets of probands with neurodevelopmental conditions, and their parents in Extended Data Figure 5. The table contains comparisons (1) between MCS children before and after reweighting to adjust for sampling bias and attrition, (2) between probands with neurodevelopmental conditions or parents (from DDD+GEL combined) with reweighted MCS children, (3) between control subsets, (4) between probands with neurodevelopmental conditions or parents (from DDD+GEL combined) with each control subset, and (5) between trio and non-trio probands with neurodevelopmental conditions and those from birth cohorts. The PGS for neurodevelopmental conditions was derived from the GWAS of DDD and UKHLS samples so it was not tested in the GWAS sample. Nominally significant *t*-test results are indicated by one asterisk and tests that pass the Bonferroni correction for five PGSs are indicated by two asterisks.

**Supplementary Table 8**. Association between the polygenic score (PGS) for educational attainment ($PGS_{EA}$) and factors affecting the chance of getting a monogenic diagnosis in DDD probands with white British ancestry affected by neurodevelopmental conditions (as shown in Figure 2BC). $PGS_{EA}$ was regressed on the indicated variable in a linear regression. The probands' $PGS_{EA}$ was tested in a maximum of 7,549 probands with neurodevelopmental conditions (without excluding Scottish samples). The fathers' or mothers' $PGS_{EA}$ was tested in a maximum of 2497 trios. Nominally significant differences are indicated by one asterisk and tests that pass the Bonferroni correction of seven factors are indicated by two asterisks. We also estimated the effect size (in odds ratio) of each factor on the chance of getting a genetic diagnosis in this subset of 7,549 probands with neurodevelopmental conditions, with and without controlling for proband's PGS for educational attainment.

**Supplementary Table 7.** Two-sided *t*-tests comparing average PGS between parents and their affected offspring from different subsets of trios, and between probands with neurodevelopmental conditions or parents and control individuals, shown in Figure 3B and Extended Data Figure 4. More specifically, the table contains comparisons (1) between undiagnosed probands with unaffected parents and their parents (in DDD+GEL combined; Figure 3B), (2) between these two groups and control individuals, (3) between parents and probands in other subsets of trios (Extended Data Figure 4), and (4) between other subsets of probands with neurodevelopmental conditions or their parents and control individuals (Extended Data Figure 4). The PGS for neurodevelopmental conditions derived from the GWAS of DDD and UKHLS samples was not tested in the GWAS samples. Nominally significant differences are indicated by one asterisk and tests that pass the Bonferroni correction for five PGSs are indicated by two asterisks.

**Supplementary Table 9.** Genetic correlations ($r_g$) between neurodevelopmental conditions and prenatal risk factors shown in Extended Data Figure 8A. We calculated $r_g$ for the GWAS meta-analysis of DDD and GEL using Linkage Disequilibrium Score Regression (LDSC). We also calculated $r_g$ between the meta-GWAS and those risk factors after conditioning on educational attainment and cognitive performance using GenomicSEM. Confidence intervals (CI) were calculated using the standard error (se) of the $r_g$ estimate. We corrected for five traits using the Bonferroni approach. For each external trait, we also report SNP heritability on the observed scale estimated by LDSC.

**Supplementary Table 10:** Number of samples and variants remaining after each step of quality control for two batches of Global Screening Array data from DDD samples, prior to merging.

**Supplementary Table 11.** Number of samples and variants remaining after each step of quality control for Global Screening Array data from DDD samples, after merging the two initial batches.

**Supplementary Table 12.** Variant quality control filters applied to whole genome sequence data from the Genomics England 100,000 Genomes variant callset known as "aggV2". We used variants that passed all these filters ("PASS" variants).

**Supplementary Table 13**. Variant quality control filters applied to whole-exome sequence data from DDD, for the analyses of polygenic scores modifying penetrance of rare coding variants and correlations between polygenic scores and rare variant burden.

**Supplementary Table 14.** Previously-published GWASs used to calculate genetic correlations and/or polygenic scores, including the number of SNPs in the polygenic score.

### Supplementary Data

**Supplementary Data 1.** Summary statistics from the GWAS of neurodevelopmental conditions comparing cases to controls within the Genomics England (GEL) 100,000 Genomes Project.

**Supplementary Data 2.** Summary statistics from the GWAS of neurodevelopmental conditions comparing DDD cases to UKHLS controls, excluding the Scottish samples from DDD.

**Supplementary Data 3.** Summary statistics from the GWAS meta-analysis of neurodevelopmental conditions combining the DDD and GEL GWASs.

### Supplementary Notes

#### Supplementary Note 1: Phenotypic comparisons of the cohorts

We compared the sex, age and Human Phenotype Ontology (HPO) terms between DDD and GEL patients with neurodevelopmental conditions included in the study. There was no significant difference in the sex of probands in DDD compared to GEL (DDD 41.0% female versus GEL 39.3% female, Fisher's exact test p-value = 0.08). DDD probands, however, were significantly younger at assessment than probands in GEL (DDD mean 7.74, standard deviation 6.29; GEL mean 10.78, standard deviation 9.10; Welch's t-test two-sided p-value = $9.51 \times 10^{-71}$) (**Extended Data Figure 2A**).

DDD probands had fewer HPO terms on average than GEL probands (DDD mean 7.28, standard deviation 4.04; GEL mean 9.34, standard deviation 5.34) (**Extended Data Figure 2B**). This difference was significant after controlling for age at assessment and sex in a multiple Poisson regression (p-value = $6.31 \times 10^{-251}$). We then compared the prevalence of HPO terms in overarching HPO chapters and some selected phenotypes in DDD and GEL, controlling for age and sex. There were significant differences between the two cohorts for numerous HPO chapters and chosen terms (**Extended Data Figure 2C**). Three notable differences between the cohorts were the greater proportion of GEL probands recorded as having autistic behaviours (31% of GEL vs 16% DDD; p-value < 0.0001), speech and language impairment (66% GEL vs 25% DDD; p-value < 0.0001), and ID/DD cases of unspecified/unknown severity (80% GEL vs 33% DDD; p-value < 0.0001).

Differences in HPO term prevalence may reflect differing practices in how terms were recorded. The DDD recruitment form asked the recruiting clinicians (who were all clinical geneticists) to record HPO terms they thought were relevant to the child's condition. In contrast, patients were recruited into GEL by a range of clinical practitioners (clinical geneticists, other speciality doctors, clinical/research nurses, genetic counsellors), who were asked to select 'yes/no/unsure' against a set of HPO terms considered to be common for patients in the phenotypic category into which the patient was recruited (e.g. 'intellectual disability'/'epilepsy plus other features'/'malformations of cortical development'). Clinical geneticists may have been more likely to select terms focused holistically on signs and symptoms of the monogenic presentation rather than non-specific HPO terms or terms relating to only one organ system (as a single organ/system specialist doctor may select). Additionally, the clinical geneticists recruiting to DDD were likely more focused on the degree of severity of ID/DD than the healthcare professionals recruiting to GEL, since they were asked a specific question about it as it was relevant to the inclusion criteria for DDD. (Patients could be included if they had moderate/severe ID/DD or if they had mild ID/DD plus other abnormalities.) These differing coding practices are likely to have artificially created HPO term differences between the cohorts rather than reflecting true clinical differences. To test this, we compared the prevalence of HPO terms in GEL versus DDD for neurodevelopmental probands who were recruited to both cohorts (N=789), controlling for age at recruitment to that study and sex. We saw similar significant differences in the HPO terms recorded for this identical set of probands between the two programs, re-enforcing these differences were likely created by differences in recording practises rather than actual differences between the GEL and DDD cohorts (**Extended Data Figure 2D**). However, it is possible that part of the reason for the increased prevalence of autistic behaviour in GEL is that the GEL recruitment of probands began a few years later than DDD (recruitment 2015-18 versus 2011-15), and the rates of diagnosed autism have been increasing over time [28].

#### Supplementary Note 2: Genome-wide significant hits from the GWAS meta-analysis of neurodevelopmental conditions

We found two genome-wide significant loci in the GWAS meta-analysis of neurodevelopmental conditions (**Extended Data Figure 3**). The locus on chromosome 22 is in an intron of *SREBF2*. Variants in this gene have been reported to be significantly associated with brain-related traits including intelligence[29–31], mathematical ability[17], and schizophrenia[19] (GWAS Catalog https://www.ebi.ac.uk/gwas/, queried on 24 Oct 2023). The lead SNP associated with neurodevelopmental conditions (rs2284084, odds ratio of risk allele T = 1.16, $p = 1.71 \times 10^{-8}$) is in weak LD with rs2267442 ($r^2 = 0.22$ estimated in 1000 Genomes GBR-ancestry subpopulation), which is associated with decreased intelligence[29,30].

The other genome-wide significant locus is located upstream of gene *PWRN4*. Lead SNP rs113446150 (odds ratio of risk allele A = 1.12, $p = 4.04 \times 10^{-8}$) is in high LD with a SNP associated with height (rs4396492; $r^2 = 0.89$)[32] reported in the GWAS Catalog. The lead SNP is a splice quantitative trait locus (sQTL) for *PWRN4* specific to the pituitary gland in GTEx. Other variants in *PWRN4* are reported to be associated with age at menarche[33].

Formal colocalization would be required to determine whether the GWAS hits from our GWAS of neurodevelopmental conditions are the same as those reported for these other traits, but the power of these is likely to be limited by the small size of our GWAS. None of the other variants within 500 kb of and in moderate or high LD with our lead SNPs (LD $r^2 > 0.5$ in 1000 Genomes GBR-ancestry subpopulation) were reported to have a significant association in the GWAS Catalog.

#### Supplementary Note 3: Potential ascertainment biases in control cohorts and their effects

In this section, we first describe how the various control cohorts were recruited and discuss whether they are likely to be biased according to educational attainment. We then consider the extent to which this affects our comparisons with these groups, and conduct a sensitivity analysis using MCS.

##### Likely ascertainment biases in control cohorts

The four control cohorts used in this study were as follows:

- The UK Household Longitudinal Study (UKHLS), which was a continuation of the British Household Panel Survey[34]. The study aimed to capture a representative sample of people living in the UK and to collect longitudinal socioeconomic and other data on them. Individuals were selected to include in the study based on their postcodes, and incentivised with monetary reward. In waves two and three of the study (2010-2012), participants aged 16 and over were invited to take part in a nurse visit, at which blood samples were taken if participants consented [35], and used to extract DNA. Those with genotype data were slightly enriched for having a university degree compared to the 2011 UK census (33.8% versus 27.2%).
- Adult cancer patients and relatives of other rare disease patients not affected by neurodevelopmental conditions or DDD-like developmental disorders from the Genomics England (GEL) 100,000 Genomes Project. GEL participants were recruited from the National Health Service, which is free at the point of care to all residents of the UK. Thus, in theory, we would not expect the control samples chosen from this cohort to be biased according to educational attainment, unless there are education/cognition-related risk factors for cancer or for rare conditions other than neurodevelopmental conditions, or unless patients who agreed to participate tended to come from a particular socioeconomic background.
- The Avon Longitudinal Study of Parents and Chidlren (ALSPAC), a birth cohort which recruited 14,775 babies born in the Avon region of southwest England in 1991-1992. About three-quarters of those eligible agreed to participate[10]. ALSPAC mothers are known to have slightly higher average socioeconomic status (SES) than mothers in the

whole of Avon and of Britain [36]. Most of the mothers' and children's DNA samples were obtained from blood samples taken at birth. Most of the fathers' DNA samples were obtained at clinics when their children were teenagers or in their early 20s.

- The Millennium Cohort Study (MCS), a birth cohort which recruited 18,827 children born 'between 1 September 2000 and 31 August 2001 (for England and Wales), and between 24 November 2000 and 11 January 2002 (for Scotland and Northern Ireland), alive and living in the UK at age 9 months, and eligible to receive child benefit at that age' [23]. Certain subgroups were intentionally over-sampled, namely children living in disadvantaged areas, children of ethnic minority backgrounds, and children growing up in the smaller nations of the UK[23]. DNA samples were taken when the probands were aged 14, at sweep 6. Only 11,872 (63%) of the original sample participated in this sweep, and they were biased towards families with higher SES compared to eligible families who did not participate [37]. Thus, this biased attrition might be expected to reduce the bias introduced by the initial over-sampling of low-SES families, but it is unclear to what extent.

Comparing average $PGS_{EA}$ between these different control cohorts can give us a sense of the relative degrees of education-related ascertainment bias (**Extended Data Figure 5A**; **Supplementary Table 6**). On average, we see that differences in PGSs between the GEL controls and UKHLS controls are small, although significant (difference in mean PGS = -0.042 SD, two-sided *t*-test p = 0.002). Within the GEL controls, we found that the cancer patients had a slightly higher average $PGS_{EA}$ than the relatives of other rare disease probands without neurodevelopmental conditions (difference in mean PGS = 0.05 SD, two-sided *t*-test p=0.013). ALSPAC children were very similar to UKHLS and GEL controls in their average $PGS_{EA}$ (on average 0.027 SD higher than GEL and 0.015 SD lower than UKHLS; two-sided *t*-test p=0.06 and 0.34), but MCS children had lower average $PGS_{EA}$ than all of the control groups (difference ranging from 0.090 to 0.132 SD, two-sided *t*-test $p < 7x10^{-9}$), likely reflecting the cohort's deliberate over-sampling of low-SES households.

In ALSPAC and MCS, as well as amongst the cases with neurodevelopmental conditions, trio probands had significantly higher $PGS_{EA}$ than probands who did not have genetic data on both parents (**Extended Data Figure 5B**; **Supplementary Table 6**) (average difference in $PGS_{EA}$ between trio and non-trio probands > 0.20 SD, two-sided *t*-test $p<3x10^{-14}$). This likely reflects the fact that families with low SES backgrounds are more likely to be single-parent households[38]. Additionally, in ALSPAC, it may reflect the fact that most of the paternal DNA samples were taken when the children were teenagers or older, so the fathers who were still engaged in the study at this point might have been biased towards higher educational attainment.

##### Sensitivity analyses to assess the effect of ascertainment bias in controls

Unsurprisingly given the results above, the estimates of differences between probands with neurodevelopmental conditions and controls are sensitive to the selection of control samples. For example, the set of all probands with neurodevelopmental conditions had significantly lower $PGS_{EA}$ than all control groups considered (two-sided *t*-test p-value $< 2x10^{-7}$), but the difference in mean ranged from -0.09 SD with the MCS children (two-sided *t*-test $p=5.2x10^{-9}$) to -0.5 SD with the ALSPAC trio children ($p=6.3x10^{-60}$; **Extended Data Figure 5A**, **Supplementary Table 6**). Since it is impossible to know which (if any) of these control cohorts are really unbiased samples of the general population, we turned to a different approach using MCS.

MCS has calculated weights based on various sociodemographic variables (e.g. SES) which can be used to reweight the individuals in the study to make them representative of the general population, in order to calculate adjusted prevalences/mean estimates and robust standard errors[23]. These include the initial sampling weights, which are intended to reweight the initial sample gathered in the first sweep, and non-response weights for each sweep. We constructed new weights for the set of MCS children who had genetic data (or specifically, the unrelated GBR-ancestry sample shown in **Extended Data Figure 5A**) and for the set who had genetic data on themselves and both parents (i.e. trio children used in **Figure 4** and **Extended Data Figure 5A**) (see **Supplementary Methods**). Applying these to recalculate the $PGS_{EA}$ for all MCS children adjusting for sampling and non-response bias, the mean $PGS_{EA}$ did not significantly change, with a mean $PGS_{EA}$ of -0.1069 (standard error, se=0.0129) prior to weighting versus -0.073 (0.0132) after (p=0.065, Wald test). When conducting the same analysis for MCS trio children, the mean $PGS_{EA}$ significantly decreased, with a mean of 0.012 (0.020) prior to weighting versus -0.055 (0.020) after (p=0.0178; **Extended Data Figure 5C**). Prior to adding weights, the difference in mean $PGS_{EA}$ between all children versus only those in trios was highly significant ($p<10^{-5}$), but was fully attenuated after weighting (p=0.45). (**Extended Data Figure 5C**). Many of the subsets of probands with neurodevelopmental conditions considered showed significantly lower $PGS_{EA}$ than the weighted MCS mean, including the undiagnosed probands ($\Delta$ = -0.17, $p=3.3\times10^{-21}$) and diagnosed probands with affected parents ($\Delta$ = -0.258, p=0.0022); however, the diagnosed probands with unaffected parents or with *de novo* diagnoses did not significantly differ from the weighted MCS sample (**Extended Data Figure 4A**; **Supplementary Table 7**).

#### Supplementary Note 4: Examining sex differences in polygenic risk

There are 1.5-times more male (N=6,879) than female (N=4,694) probands with neurodevelopmental conditions in DDD and GEL combined, which is consistent with the “female protective effect” whereby females either have lower mean liability for neurodevelopmental conditions, or require higher liability than males to get a diagnosis [39–48]. Indeed, female patients in DDD are more likely to get a monogenic diagnosis [20] and have a higher burden of damaging *de novo* mutations than males [49]. We did not detect any significant differences in PGS between male and female undiagnosed probands with neurodevelopmental conditions for any PGS (**Extended Data Figure 7A**). This is not inconsistent with recent work in autism: Wigdor et al. found that autistic females have higher autism PGS than males after accounting for co-occurring ID[45], but saw no significant difference otherwise. Similarly, Warrier et al. showed that amongst autistic individuals without ID, females over-inherited more polygenic risk for autism than males[50]., but did not detect a difference when including individuals with ID. In our data, although $PGS_{NDC,DDD}$ was significantly over-transmitted in females (pTDT deviation = 0.10, p-value = 0.0078, N=589 trios in DDD and GEL combined) but not males (pTDT deviation = 0.036, p-value = 0.27, N=978 trios), there was no significant difference in the pTDT deviation between them (two-sided z-test p=0.19) (**Extended Data Figure 7C**). Notably, in contrast to our findings, Antaki et al. showed that autistic females have higher values for a combined PGS for autism + educational attainment + schizophrenia than autistic males [51]. Our findings from these sex comparisons emphasise that ID, which is present in the majority of DDD and GEL NDC probands, has a distinct genetic architecture from autism more broadly.

In families of autistic children in whom neither parent has a known autism diagnosis, mothers were found to have a higher PGS for autism than fathers, possibly reflecting the fact that women can 'tolerate' a higher burden of risk alleles before manifesting the phenotype due to the so-called "female protective effect" [45]. Thus, we also compared the five NDC-related PGSs between mothers and fathers, focusing on unaffected parents of undiagnosed probands (**Extended Data Figure 7B**). We found no significant differences.

#### Supplementary Note 5: Exploring the role of prenatal risk factors in mediating common variant risk

##### Genetic correlations

We hypothesized that the effects of non-transmitted parental alleles on risk of neurodevelopmental conditions (**Figure 4**) may be partly mediated by the prenatal environment, aspects of which are associated with risk of these conditions. For example, being born prematurely is a risk factor for neurodevelopmental conditions[52–54], and we see a negative genetic correlation between preterm delivery [55] and educational attainment ($r_g$=-0.30; p=2x$10^{-10}$). This correlation may be partly explained by the fact that women with lower educational levels are more likely to have risk factors for premature birth, such as exposure to tobacco smoke during pregnancy[56,57]. Within DDD, lower $PGS_{EA}$ and $PGS_{NonCogEA}$ in mothers was significantly associated with the proband having been born prematurely (**Extended Data Figure 8B**).

To explore the potential contribution of prenatal risk factors to polygenic risk for neurodevelopmental conditions, we calculated genetic correlations between our GWAS meta-analysis and risk factors for which GWASs were available: preterm delivery [52–55], smoking[54,58], alcohol use[54,58], gestational hypertension[54,59] and sleep apnoea[60–62] (**Extended Data Figure 8A**). We observed significant genetic correlations between neurodevelopmental conditions and preterm delivery ($r_g$=0.58; p=0.004) and smoking initiation ($r_g$=0.27; p=2x$10^{-5}$). After conditioning on the educational attainment GWAS with GenomicSEM, the genetic correlation with smoking initiation was greatly attenuated and no longer significant ($r_g$=0.04; p=0.62), while that with preterm delivery was slightly attenuated but still nominally significant ($r_g$=0.48; p=0.04) (**Supplementary Table 9**). Conversely, after conditioning on the preterm delivery GWAS, the genetic correlation between educational attainment and neurodevelopmental conditions was still highly significant ($r_g$=-0.60, p=8x$10^{-7}$), although slightly attenuated compared to the unconditional result ($r_g$=-0.65, p=5x$10^{-12}$).

##### Mendelian randomization

Given these genetic correlation results, we explored potential causal relationships between educational attainment, preterm delivery and neurodevelopmental conditions using different Mendelian Randomization (MR) methods (**Extended Data Figure 9**; **Supplementary Methods**). We hypothesized that the following causal effects exist:

- a causal effect of lower educational attainment on neurodevelopmental conditions that reflects indirect effects via the parents
- a causal effect of lower educational attainment on preterm delivery, reflecting the association between lower education levels and risk factors for preterm delivery (e.g. a short inter-pregnancy interval[63], exposure to tobacco smoke during pregnancy[56,57], and pre-eclampsia[64]
- a causal effect of preterm delivery on lower educational attainment and on neurodevelopmental conditions, reflecting the strong associations between premature

birth and adverse neurodevelopmental outcomes that hold after controlling for socioeconomic confounders [52,65–71]

We employed several standard two-sample MR methods using genome-wide significant SNPs from the GWAS for the exposure, as well as the Latent Heritable Confounder-Mendelian randomisation (LHC-MR)[22] method which exploits genome-wide SNPs. Consistent with our hypothesis, two out of the four standard methods tested showed statistically significant evidence that lower educational attainment causally increases risk for neurodevelopmental conditions (weighted median p-value = $9.2 \times 10^{-8}$ and inverse variance weighted p-value = $5.2 \times 10^{-18}$), as did LHC-MR ($p = 6.3 \times 10^{-44}$) (**Extended Data Figure 9A**). Also consistent with the hypotheses above, LHC-MR showed significant negative bi-directional causal effects between educational attainment and preterm delivery ($p = 3.9 \times 10^{-12}$ and p = 0.016 for the forward and reverse causal effects, respectively). However, a caveat of this is that the inferred causal effect of preterm delivery on educational attainment could simply reflect the effect of transmitted variants from the mother with predispose to lower educational attainment; we acknowledge that LHC-MR is not intended to test for the kind of intergenerational causality we hypothesize may be at play here, and it also assumes a single heritable confounding variable and can give biased estimates if this is violated[22]. Using standard Mendelian randomization, we found only weak evidence for lower educational attainment being causally associated with preterm delivery with only the inverse variance weighted method being significant ($p = 1.5 \times 10^{-5}$). We did not find significant evidence that preterm delivery causally influences neurodevelopmental conditions or educational attainment using standard MR methods (**Extended Data Figure 9BC**). However, this analysis is limited by the relatively small GWASs of neurodevelopmental conditions and preterm delivery. Specifically, these underpowered GWASs lead to noisy estimates when they are considered as outcomes in both standard MR and LHC-MR analyses, and additionally, Winner's Curse (i.e. overestimation of effect sizes) in the original GWAS of the exposure will reduce power in standard MR, which uses genome-wide significant SNPs as genetic instruments. Furthermore, when considering them as exposures, power was limited by weak instrument bias in the standard MR analyses, which biases the estimate towards the null, and by the fact that the preterm delivery GWAS was conducted in mothers whereas our GWAS was done in offspring. Finally, we note that MR based on population-based GWAS is prone to confounding, and these analyses should be revisited once sufficiently large within-family GWASs are available[72].

##### Testing whether prematurity mediates the effect of non-transmitted common alleles

Based on these results, we hypothesized that prematurity may partly mediate the effect of non-transmitted common variants associated with educational attainment on risk of neurodevelopmental conditions. To test this, we reran the "trio model" from **Figure 4** using $PGS_{EA}$ in several different ways, restricting to undiagnosed DDD cases and MCS controls for which information on gestational age at birth was available (**Supplementary Figure 5A**). After excluding probands born prematurely from both cases (16%) and controls (6%), the effect of non-transmitted parental alleles was slightly attenuated compared to that seen when using all trio probands, particularly in mothers ($\beta_{mother}$ = -0.14 [-0.23 – -0.06] and $p=6 \times 10^{-4}$ using all samples, versus $\beta_{mother}$ = -0.11 [-0.20 – -0.02] and p=0.02 after excluding probands born prematurely). A similar result was obtained when using all probands but controlling for whether they were born prematurely. We would expect such attenuation if there were an indirect genetic effect of $PGS_{EA}$ mediated partly via premature delivery. However, the effect sizes were not significantly altered for any PGS in either analysis compared to the original analysis of all trio probands (z-test p>0.05). Thus, there is no significant evidence from these data that indirect genetic effects mediated through prematurity contribute to the association between non-transmitted common variants of $PGS_{EA}$ and risk of neurodevelopmental conditions.

#### Supplementary Note 6: Role of PGS in modifying the penetrance of rare variants

Various lines of evidence from both neurodevelopmental conditions [73] and autism [39,74] cohorts and from population-based studies [75] suggest that incompletely penetrant inherited rare coding variants contribute to risk of rare neurodevelopmental conditions. A recent study found that undiagnosed rare disease patients with variants of unknown significance (VUS) had on average more polygenic risk than their unaffected carrier parent, suggesting PGS might modify penetrance of VUS [76]. Rare protein-truncating variants (PTVs) and predicted damaging missense variants in constrained (loss-of-function-intolerant) genes[77] have been shown to act additively with PGSs on fluid intelligence and educational attainment within UK Biobank [78,79]. This implies that PGSs are likely to modify penetrance of these rare variants within families with neurodevelopmental conditions.

To test this, we used the whole-genome sequence data from GEL and exome sequence data from DDD to extract rare damaging coding variants from undiagnosed probands and their parents; specifically, we extracted heterozygous PTVs and predicted damaging missense variants with minor allele frequency < $1x10^{-4}$ within each cohort and < $1x10^{-5}$ in each gnomAD super-population in either dominant DD-associated genes (DDG2P)[80] with a loss-of-function mechanism, or in constrained genes (see **Methods**). In unpublished work, such variants are seen to be enriched in undiagnosed DDD cases compared to controls, and over-transmitted from unaffected parents to affected offspring (K. Samocha, personal communication). We tested whether unaffected parents transmitting a damaging rare variant have significantly more protective PGSs than their affected children without a monogenic diagnosis. We observed nominally significant differences in $PGS_{NDC,DDD}$ between parents who transmitted a damaging rare variant in a dominant DD-associated gene and their children in a combined analysis of DDD and GEL (one-sided p=0.009, mean difference = -0.16 SD, N=186 pairs; **Supplementary Figure 8**). However, none of the differences passed multiple testing correction (p>0.05/[5 PGSs * 2 gene sets * 2 consequence classes]) (**Supplementary Figure 8**).

The interpretation of these results is potentially complicated by several factors, including the correlation between rare and common variants that is likely generated by parental assortment (**Figure 5**). Power may be reduced by our limited sample size and aggregation of rare variants with heterogeneous effects; it may be that many of the rare variants considered are not damaging, but due to their rarity, we aggregated across variants and genes to try to boost power. Additionally, if some parents actually show sub-clinical phenotypes as a result of these rare variants or polygenic burden, the inclusion of these parents in the analyses could be confounding these results.

### References

1. Niemi, M. E. K. *et al.* Common genetic variants contribute to risk of rare severe neurodevelopmental disorders. *Nature* **562**, 268–271 (2018).
2. Deciphering Developmental Disorders Study. Large-scale discovery of novel genetic causes of developmental disorders. *Nature* **519**, 223–228 (2015).
3. 1000 Genomes Project Consortium *et al.* A global reference for human genetic variation. *Nature* **526**, 68–74 (2015).
4. Patterson, N., Price, A. L. & Reich, D. Population structure and eigenanalysis. *PLoS Genet.* **2**, e190 (2006).
5. Price, A. L. *et al.* Long-range LD can confound genome scans in admixed populations. *American journal of human genetics* vol. 83 132–5; author reply 135–9 (2008).
6. McInnes, L., Healy, J. & Melville, J. UMAP: Uniform Manifold Approximation and Projection for Dimension Reduction. *arXiv [stat.ML]* (2018).
7. Kousathanas, A. *et al.* Whole-genome sequencing reveals host factors underlying critical COVID-19. *Nature* **607**, 97–103 (2022).
8. Purcell, S. *et al.* PLINK: a tool set for whole-genome association and population-based linkage analyses. *Am. J. Hum. Genet.* **81**, 559–575 (2007).
9. Yang, J., Lee, S. H., Goddard, M. E. & Visscher, P. M. GCTA: a tool for genome-wide complex trait analysis. *Am. J. Hum. Genet.* **88**, 76–82 (2011).
10. Boyd, A. *et al.* Cohort Profile: the ‘children of the 90s’--the index offspring of the Avon Longitudinal Study of Parents and Children. *Int. J. Epidemiol.* **42**, 111–127 (2013).
11. Manichaikul, A. *et al.* Robust relationship inference in genome-wide association studies. *Bioinformatics* **26**, 2867–2873 (2010).
12. Fitzsimons, E. *et al.* Collection of genetic data at scale for a nationally representative population: the UK Millennium Cohort Study. *Longit. Life Course Stud.* **13**, 169–187 (2021).
13. Mills, R. E. *et al.* An initial map of insertion and deletion (INDEL) variation in the human

genome. *Genome Res.* **16**, 1182–1190 (2006).

14. Vilhjálmsson, B. J. *et al.* Modeling Linkage Disequilibrium Increases Accuracy of Polygenic Risk Scores. *Am. J. Hum. Genet.* **97**, 576–592 (2015).
15. International HapMap 3 Consortium *et al.* Integrating common and rare genetic variation in diverse human populations. *Nature* **467**, 52–58 (2010).
16. Bycroft, C. *et al.* The UK Biobank resource with deep phenotyping and genomic data. *Nature* **562**, 203–209 (2018).
17. Lee, J. J. *et al.* Gene discovery and polygenic prediction from a genome-wide association study of educational attainment in 1.1 million individuals. *Nat. Genet.* **50**, 1112–1121 (2018).
18. Demange, P. A. *et al.* Investigating the genetic architecture of noncognitive skills using GWAS-by-subtraction. *Nat. Genet.* **53**, 35–44 (2021).
19. Trubetskoy, V. *et al.* Mapping genomic loci implicates genes and synaptic biology in schizophrenia. *Nature* **604**, 502–508 (2022).
20. Wright, C. F. *et al.* Genomic Diagnosis of Rare Pediatric Disease in the United Kingdom and Ireland. *N. Engl. J. Med.* **388**, 1559–1571 (2023).
21. Tingley, D., Yamamoto, T., Hirose, K., Keele, L. & Imai, K. mediation: R package for causal mediation analysis. (2014).
22. Darrous, L., Mounier, N. & Kutalik, Z. Simultaneous estimation of bi-directional causal effects and heritable confounding from GWAS summary statistics. *Nat. Commun.* **12**, 7274 (2021).
23. Plewis, I. The Millennium Cohort Study: Technical Report on Sampling (4th Edition). http://doc.ukdataservice.ac.uk/doc/4683/mrdoc/pdf/mcs_technical_report_on_sampling_4th_edition.pdf (2007).
24. Plewis, I. Non-Response in a Birth Cohort Study: The Case of the Millennium Cohort Study. *Int. J. Soc. Res. Methodol.* **10**, 325–334 (2007).
25. Yang, J. *et al.* Genetic variance estimation with imputed variants finds negligible missing heritability for human height and body mass index. *Nat. Genet.* **47**, 1114–1120 (2015).

26. Golan, D., Lander, E. S. & Rosset, S. Measuring missing heritability: inferring the contribution of common variants. *Proc. Natl. Acad. Sci. U. S. A.* **111**, E5272–81 (2014).

27. Churchhouse, C. Insights from estimates of SNP-heritability for >2,000 traits and disorders in UK Biobank. *Neale lab* http://www.nealelab.is/blog/2017/9/20/insights-from-estimates-of-snp-heritability-for-2000-traits-and-disorders-in-uk-biobank (2017).

28. Russell, G. *et al.* Time trends in autism diagnosis over 20 years: a UK population-based cohort study. *J. Child Psychol. Psychiatry* **63**, 674–682 (2022).

29. Davies, G. *et al.* Study of 300,486 individuals identifies 148 independent genetic loci influencing general cognitive function. *Nat. Commun.* **9**, 2098 (2018).

30. Savage, J. E. *et al.* Genome-wide association meta-analysis in 269,867 individuals identifies new genetic and functional links to intelligence. *Nat. Genet.* **50**, 912–919 (2018).

31. Williams, C. M., Labouret, G., Wolfram, T., Peyre, H. & Ramus, F. A General Cognitive Ability Factor for the UK Biobank. *Behav. Genet.* **53**, 85–100 (2023).

32. Yengo, L. *et al.* A saturated map of common genetic variants associated with human height. *Nature* **610**, 704–712 (2022).

33. Perry, J. R. *et al.* Parent-of-origin-specific allelic associations among 106 genomic loci for age at menarche. *Nature* **514**, 92–97 (2014).

34. Knies, G. Understanding society: waves 1–7, 2009–2016 and harmonised BHPS: waves 1–18, 1991–2009, user guide. *Colchester: University of Essex*.

35. of Essex, U. Institute for Social and Economic Research and National Centre for Social Research, Understanding Society: Waves 2 and 3 Nurse Health Assessment …. *UK Data Service*.

36. Fraser, A. *et al.* Cohort Profile: the Avon Longitudinal Study of Parents and Children: ALSPAC mothers cohort. *Int. J. Epidemiol.* **42**, 97–110 (2013).

37. Mostafa, T. & Ploubidis, G. Millennium Cohort Study. https://discovery.ucl.ac.uk/id/eprint/10060140/1/mcs6_report_on_response.pdf.

38. Morelli, S., Nolan, B., Palomino, J. C. & Van Kerm, P. The Wealth (Disadvantage) of Single-Parent Households. *Ann. Am. Acad. Pol. Soc. Sci.* **702**, 188–204 (2022).

39. Fu, J. M. *et al.* Rare coding variation provides insight into the genetic architecture and phenotypic context of autism. *Nat. Genet.* **54**, 1320–1331 (2022).

40. Satterstrom, F. K. *et al.* Large-Scale Exome Sequencing Study Implicates Both Developmental and Functional Changes in the Neurobiology of Autism. *Cell* **180**, 568–584.e23 (2020).

41. Sanders, S. J. *et al.* Insights into Autism Spectrum Disorder Genomic Architecture and Biology from 71 Risk Loci. *Neuron* **87**, 1215–1233 (2015).

42. Sanders, S. J. *et al.* De novo mutations revealed by whole-exome sequencing are strongly associated with autism. *Nature* **485**, 237–241 (2012).

43. Ronemus, M., Iossifov, I., Levy, D. & Wigler, M. The role of de novo mutations in the genetics of autism spectrum disorders. *Nat. Rev. Genet.* **15**, 133–141 (2014).

44. Iossifov, I. *et al.* The contribution of de novo coding mutations to autism spectrum disorder. *Nature* **515**, 216–221 (2014).

45. Wigdor, E. M. *et al.* The female protective effect against autism spectrum disorder. *Cell Genomics* **2**, 100134 (2022).

46. Robinson, E. B., Lichtenstein, P., Anckarsäter, H., Happé, F. & Ronald, A. Examining and interpreting the female protective effect against autistic behavior. *Proc. Natl. Acad. Sci. U. S. A.* **110**, 5258–5262 (2013).

47. Baron-Cohen, S. *et al.* Prevalence of autism-spectrum conditions: UK school-based population study. *Br. J. Psychiatry* **194**, 500–509 (2009).

48. Fombonne, E. Epidemiology of pervasive developmental disorders. *Pediatr. Res.* **65**, 591–598 (2009).

49. Kaplanis, J. *et al.* Evidence for 28 genetic disorders discovered by combining healthcare and research data. *Nature* **586**, 757–762 (2020).

50. Warrier, V. *et al.* Genetic correlates of phenotypic heterogeneity in autism. *Nat. Genet.* **54**, 1293–1304 (2022).

51. Antaki, D. *et al.* A phenotypic spectrum of autism is attributable to the combined effects of rare variants, polygenic risk and sex. *Nat. Genet.* 1–9 (2022).

52. Joseph, R. M. *et al.* Neurocognitive and Academic Outcomes at Age 10 Years of Extremely Preterm Newborns. *Pediatrics* **137**, (2016).
53. Aarnoudse-Moens, C. S. H., Weisglas-Kuperus, N., van Goudoever, J. B. & Oosterlaan, J. Meta-analysis of neurobehavioral outcomes in very preterm and/or very low birth weight children. *Pediatrics* **124**, 717–728 (2009).
54. Huang, J., Zhu, T., Qu, Y. & Mu, D. Prenatal, Perinatal and Neonatal Risk Factors for Intellectual Disability: A Systemic Review and Meta-Analysis. *PLoS One* **11**, e0153655 (2016).
55. Solé-Navais, P. *et al.* Genetic effects on the timing of parturition and links to fetal birth weight. *Nat. Genet.* **55**, 559–567 (2023).
56. Kandel, D. B., Griesler, P. C. & Schaffran, C. Educational attainment and smoking among women: risk factors and consequences for offspring. *Drug Alcohol Depend.* **104 Suppl 1**, S24–33 (2009).
57. Goldenberg, R. L., Culhane, J. F., Iams, J. D. & Romero, R. Epidemiology and causes of preterm birth. *Lancet* **371**, 75–84 (2008).
58. Saunders, G. R. B. *et al.* Genetic diversity fuels gene discovery for tobacco and alcohol use. *Nature* **612**, 720–724 (2022).
59. Honigberg, M. C. *et al.* Polygenic prediction of preeclampsia and gestational hypertension. *Nat. Med.* **29**, 1540–1549 (2023).
60. Xu, T., Feng, Y., Peng, H., Guo, D. & Li, T. Obstructive sleep apnea and the risk of perinatal outcomes: a meta-analysis of cohort studies. *Sci. Rep.* **4**, 6982 (2014).
61. Campos, A. I. *et al.* Discovery of genomic loci associated with sleep apnea risk through multi-trait GWAS analysis with snoring. *Sleep* **46**, (2023).
62. Pamidi, S. *et al.* Maternal sleep-disordered breathing and adverse pregnancy outcomes: a systematic review and metaanalysis. *Am. J. Obstet. Gynecol.* **210**, 52.e1–52.e14 (2014).
63. Thoma, M. E., Copen, C. E. & Kirmeyer, S. E. Short Interpregnancy Intervals in 2014: Differences by Maternal Demographic Characteristics. *NCHS Data Brief* 1–8 (2016).

64. Granés, L., Torà-Rocamora, I., Palacio, M., De la Torre, L. & Llupià, A. Maternal educational level and preterm birth: Exploring inequalities in a hospital-based cohort study. *PLoS One* **18**, e0283901 (2023).
65. Potharst, E. S. *et al.* High incidence of multi-domain disabilities in very preterm children at five years of age. *J. Pediatr.* **159**, 79–85 (2011).
66. Wong, H. S. & Edwards, P. Nature or nurture: a systematic review of the effect of socio-economic status on the developmental and cognitive outcomes of children born preterm. *Matern. Child Health J.* **17**, 1689–1700 (2013).
67. Cheong, J. L. Y. *et al.* Changing Neurodevelopment at 8 Years in Children Born Extremely Preterm Since the 1990s. *Pediatrics* **139**, (2017).
68. Beauregard, J. L., Drews-Botsch, C., Sales, J. M., Flanders, W. D. & Kramer, M. R. Does Socioeconomic Status Modify the Association Between Preterm Birth and Children's Early Cognitive Ability and Kindergarten Academic Achievement in the United States? *Am. J. Epidemiol.* **187**, 1704–1713 (2018).
69. Lacalle, L., Martínez-Shaw, M. L., Marín, Y. & Sánchez-Sandoval, Y. Intelligence Quotient (IQ) in school-aged preterm infants: A systematic review. *Front. Psychol.* **14**, 1216825 (2023).
70. Crump, C., Sundquist, J. & Sundquist, K. Preterm or early term birth and risk of attention-deficit/hyperactivity disorder: a national cohort and co-sibling study. *Ann. Epidemiol.* **86**, 119–125.e4 (2023).
71. Husby, A., Wohlfahrt, J. & Melbye, M. Gestational age at birth and cognitive outcomes in adolescence: population based full sibling cohort study. *BMJ* **380**, e072779 (2023).
72. Howe, L. J. *et al.* Within-sibship genome-wide association analyses decrease bias in estimates of direct genetic effects. *Nat. Genet.* **54**, 581–592 (2022).
73. Wright, C. F. *et al.* Evaluating variants classified as pathogenic in ClinVar in the DDD Study. *Genet. Med.* **23**, 571–575 (2021).
74. Wilfert, A. B. *et al.* Recent ultra-rare inherited variants implicate new autism candidate risk genes. *Nat. Genet.* **53**, 1125–1134 (2021).

75. Kingdom, R. *et al.* Rare genetic variants in genes and loci linked to dominant monogenic developmental disorders cause milder related phenotypes in the general population. *The American Journal of Human Genetics* Preprint at https://doi.org/10.1016/j.ajhg.2022.05.011 (2022).

76. Smail, C. *et al.* Complex trait associations in rare diseases and impacts on Mendelian variant interpretation. *medRxiv* 2024.01.10.24301111 (2024) doi:10.1101/2024.01.10.24301111.

77. Lek, M. *et al.* Analysis of protein-coding genetic variation in 60,706 humans. *Nature* **536**, 285–291 (2016).

78. Chen, C.-Y. *et al.* The impact of rare protein coding genetic variation on adult cognitive function. *Nat. Genet.* **55**, 927–938 (2023).

79. Kingdom, R., Beaumont, R. N., Wood, A. R., Weedon, M. N. & Wright, C. F. Genetic modifiers of rare variants in monogenic developmental disorder loci. *medRxiv* (2022) doi:10.1101/2022.12.15.22283523.

80. DECIPHER: Database of Chromosomal Imbalance and Phenotype in Humans Using Ensembl Resources. *Am. J. Hum. Genet.* **84**, 524–533 (2009).
